# Predicting individualized treatment effects of corticosteroids in community-acquired-pneumonia: a data-driven analysis of randomized controlled trials

**DOI:** 10.1101/2023.10.03.23296132

**Authors:** J.M. Smit, P.A. Van Der Zee, S.C.M. Stoof, M.E. Van Genderen, D. Snijders, W. G. Boersma, P. Confalonieri, F. Salton, M. Confalonieri, M-C. Shih, G.U. Meduri, P.-F. Dequin, A. Le Gouge, M. Lloyd, H. Karunajeewa, G. Bartminski, S. Fernández-Serrano, G. Suárez-Cuartín, D. van Klaveren, M. Briel, C.M. Schoenenberger, E.W. Steyerberg, D.A.M.P.J. Gommers, H.I. Bax, W J. W. Bos, E.M.W. Van De Garde, E. Wittermans, J.C. Grutters, C.A. Blum, M. Christ-Crain, A. Torres, A. Motos, M.J.T. Reinders, J. Van Bommel, J.H. Krijthe, H. Endeman

## Abstract

**Background:** Corticosteroids could improve outcomes in patients with community-acquired pneumonia (CAP). However, we hypothesize that corticosteroid effectiveness varies among individual patients, resulting in inconsistent outcomes and unclear clinical indication. Therefore, we developed and validated a predictive, causal model based on baseline characteristics to predict individualized treatment effects (ITEs) of corticosteroids on mortality in patients with CAP.

**Methods:** We obtained individual patient data from six randomized controlled trials comparing corticosteroid therapy to placebo in 1,869 adult CAP patients. The study endpoint was 30-day mortality. We performed effect modelling through logistic regression and evaluated the predicted ITEs in terms of discrimination and calibration for benefit. Our modelling procedure involved variable selection, missing value imputation, data normalization, encoding treatment variables, creating interaction terms, optimizing penalization strength, and training logistic regression models. We evaluated discriminative performance using the newly proposed ‘AUC-benefit’.

**Findings:** The model identified high levels of CRP and glucose, at baseline, as main predictors for benefit of corticosteroid treatment. Using a decision threshold of ITE=0, the model predicted harm in 1,004 patient and benefit in 864 patients. We observed benefit in patients where the model predicted benefit, with an odds ratio of 0.5 (95% CI: 0.3 to 0.9) and a mortality reduction of 3.2% (95% CI: 0.7 to 5.6), and no statistically significant benefit in the patients where the model predicted harm, with an odds ratio of 1.1 (95% CI: 0.7 to 1.8) and a negative mortality reduction (hence, increase) of −0.3% (95% CI: −2.6 to 1.8). The model yielded an AUC-benefit of 184.9 (28.6 to 347.6, 95% CI), underestimated ITEs in the lower ITE region and slightly overestimated ITEs in the higher ITE region.

**Interpretation:** Our model has potential to identify patients with CAP who benefit from corticosteroid treatment, and aid in the design of personalized clinical trials. We will prospectively validate the model in two recent CAP trials.

## 1. Introduction

Community-acquired pneumonia (CAP) is a significant cause of hospitalization and has a high mortality rate.^1,2^ Adjuvant treatment with corticosteroids could reduce the excessive systemic inflammatory response, which is associated with increased mortality rate.^3,4^ Despite multiple, randomized controlled trials (RCTs) on the use of adjuvant treatment with corticosteroids in patients with CAP, the effect on mortality remains controversial.^5^ Therefore, routine use of corticosteroids is not recommended.^6^ However, an individual patient data meta-analysis suggested a larger benefit from corticosteroid treatment in patients with more severe pneumonia, indicating heterogeneity in treatment effect (HTE).^7^ Understanding how these effects vary based on baseline characteristics could assist in the identification of patients with CAP who would likely benefit from adjuvant treatment with corticosteroids, potentially leading to a more personalized treatment.^8–10^ Conventional subgroup analyses, which are based on single variables (such as age or C-reactive protein), are limited by low statistical power, multiple testing, and the inability to integrate multiple patient characteristics simultaneously. A predictive HTE analysis aims to address these limitations by providing predictions of individualized treatment effect (ITEs).^8^ The ITE is the difference between the predicted outcome under corticosteroid treatment versus placebo in a specific patient, by considering multiple relevant patient characteristics simultaneously. The aim of this study is to develop and validate a model that predicts the ITE of adjuvant treatment with corticosteroids in patients with CAP on 30-day mortality. Thereafter, we will prospective validation in the two most recent trials for adjuvant treatment with corticosteroids in patients with CAP.^11,12^

## 2. Methods

### 2.1 Trial Selection and Data Collection

We performed an update of the systematic search by Briel et al.,^7^ which identified RCTs that compared adjunctive therapy with oral or intravenous corticosteroid to placebo in hospitalized patients with CAP aged 18 years or older, up to July 2017. Eligibility screening and risk of bias assessment were performed and are described in full detail in appendix A. We identified 10 eligible trials and obtained data from six trials (supplementary Figure A1).^13–19^ From six of these eligible trials, we obtained individual patient data (IPD), which consisted of age, sex, six clinical parameters, six laboratory values at baseline (ie, hospital admission), comorbidities and the pneumonia severity index (PSI).^20^ Additionally, one observational data set of prospectively enrolled hospitalized patients with CAP was used for data imputation in the trial datasets.^21^

### 2.2 Study endpoint

The endpoint of our study was 30-days mortality rate. For each patient, we defined the ITE as the predicted probability of 30-day mortality with placebo treatment minus the predicted probability of 30-day mortality with corticosteroid treatment (Figure 1).

**Figure 1:**
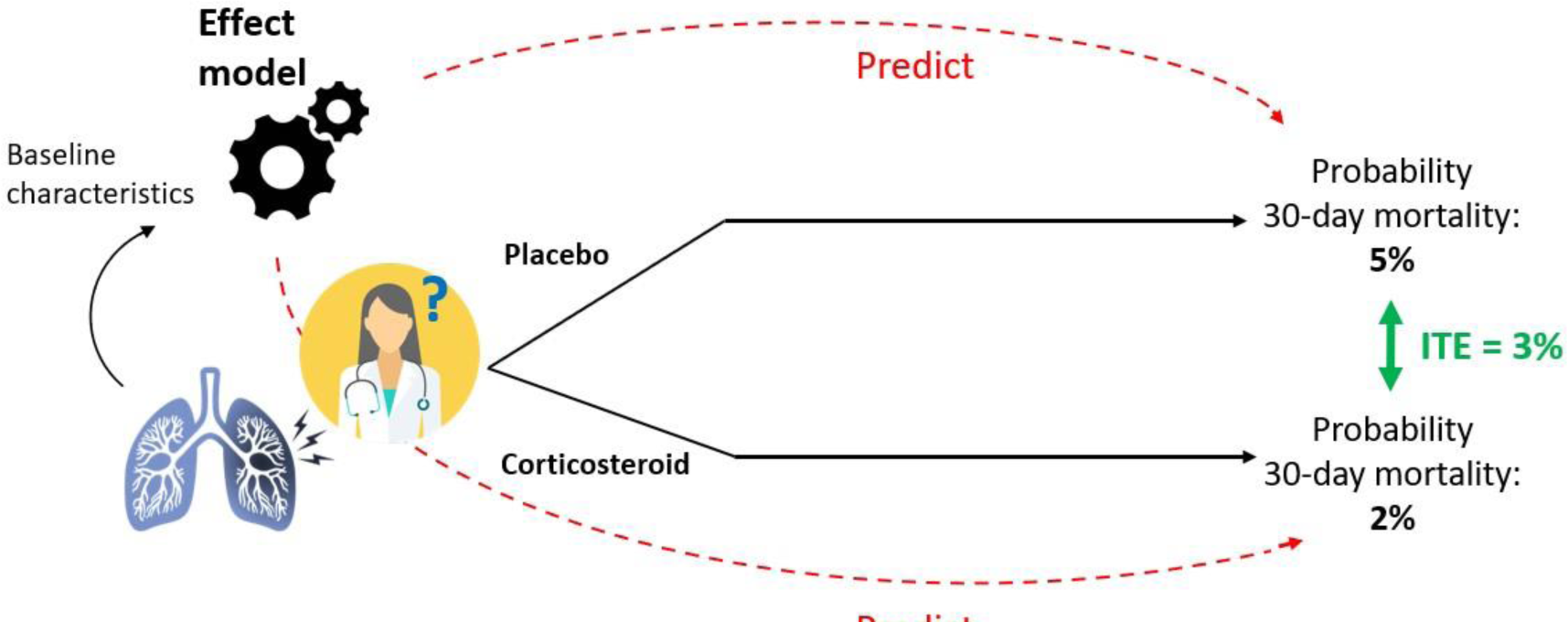
Schematic overview of individualized treatment effect (ITE) prediction using effect modelling.

### 2.3 Model Evaluation

We evaluated predicted ITEs in terms of discrimination for benefit and calibration for benefit, which assess the model’s ability to rank patients based on the benefit they would derive from adjuvant treatment with corticosteroids, and the agreement between predicted reduction in mortality and true reduction in mortality, respectively.^22^ Due to the unobservability of true ITEs (ie, the fundamental problem of causal inference^23^), conventional metrics for model discrimination and calibration cannot be used.

#### 2.3.1 Discrimination for benefit

To assess discrimination for benefit, we introduced the ‘area under the benefit curve’ (AUC-benefit). The AUC-benefit considers different ITE thresholds to divide patients into a lower ITE group (where ITE ≤ threshold) and a higher ITE group (where ITE > threshold, supplementary Figure B1). For each group, we measure the treatment effect in terms of *benefit*, ie, the mortality rate in patients who received placebo minus the mortality rate in patients who received adjuvant treatment with corticosteroids. The difference in benefit (ie, the ‘Δ-benefit’) is then calculated as the benefit in the higher ITE group minus the benefit in the lower ITE group. By calculating Δ-benefits for various thresholds, a ‘Δ-benefit-curve’ is created. In our analysis, we started with a threshold at the 25^th^ percentile, and increase the percentiles in ten equal steps until the 75^th^ percentile of the full ITE distribution. The AUC-benefit represents the trapezoidal area under the Δ-benefit-curve (supplementary Figure B2). Models with higher AUC-benefit indicate better discriminative performance (refer to Appendix B for details).

#### 2.3.2 Calibration for benefit

To evaluate calibration for benefit, we divided patients into four groups based on ascending ITE quartiles and plotted the ITE distributions (using violin plots) next to the observed mortality reductions in each quartile.

#### 2.3.3 Predicted harm vs Predicted benefit

Finally, assuming a *decision threshold* (ie, an ITE value above which treating patients is considered worthwhile) at 0, we divided patients in a group of those who would have been advised for treatment (with ITE > 0) and those who would have been advised against treatment (with ITE ≤ 0) by the model, referred to as the ‘predicted benefit’ and ‘predicted harm group’, respectively. For these groups, we presented observed mortality rates in the treatment arms, observed treatment effects in terms of odds ratios and mortality reduction, and the number needed to treat (NNT), defined as the average number of patients that need to be treated to prevent one additional death. Furthermore, we tested HTE between the predicted harm and predicted benefit group using an interaction test by fitting a logistic regression model for mortality, using group assignment, treatment assignment, and their interaction as covariates.

### 2.4 Train-Test split

We used so-called ‘leave-one-trial-out’ cross-validation (LOTO-CV) to validate the full modelling procedure (which we describe in the next section). That is, in six iterations (of ‘folds’), IPD from five trials formed the train cohort, and one trial is held out to form the test cohort (supplementary Figure F1). The ITEs predicted for patients in the test cohorts in each iteration are combined and then evaluated.

### 2.5 Modelling Procedure

We used effect modeling^8^ to directly predict 30-day mortality using IPD from the trials in the train cohort. Specifically, we utilized a method based on the least absolute shrinkage and selection operator (LASSO) regression, as proposed by Tian et al.^24^ The exact implementation for the LASSO penalty we used can be found in appendix C.

The modelling procedure comprised seven steps (supplementary Figure F1). Step one involved including variables based on availability: those available for at least two-thirds of patients in both train and test cohorts were included. In step two, missing values were imputed and data normalization was performed (described further in section 2.4). Step three encoded the treatment variable (placebo = −1, corticosteroids = 1). Step four involved creating interaction terms by multiplying the included variables with the encoded treatment variable and adding them, together with the encoded treatment variable, to the logistic regression model. Step five optimized the penalization strength (λ) through two grid searches using the train cohort only. The first grid search used a default wide grid (supplementary Table F1), and for each candidate λ, a ‘nested LOTO-CV’ was performed using the train cohort (supplementary Figure F2). Here, in five iterations, combined IPD from four trials formed the ‘inner train cohort’, and the held out trial the ‘inner test cohort’. the first four modelling steps were repeated using the inner train cohort, and a penalty term was added using the candidate λ, whereafter the model is trained and ITEs are predicted for the patients in the inner test cohort. Candidate λs resulting in zero weights for interaction terms and the treatment variable (ie, resulting in zero ITEs only) in at least one of the nested folds, were not considered. The predicted ITEs from the five iterations were then combined and we took 1000 bootstrap samples. For each bootstrap sample, we calculated the AUC-benefit, and the λ that yielded the highest median AUC-benefit (ie, the optimal λ) was used to define the center point of a finer grid for the second grid search (supplementary Table F1), and the nested LOTO-CV is repeated using this fine grid. In step six, the optimal λ found in step 5 is used to train the penalized logistic regression model using all data from the train cohort, penalizing both treatment variable and interaction terms. Finally (ie, step seven), this trained model is used to predict ITEs for patients in the test cohort. Hence, the outer LOTO-CV procedure will result in six trained logistic regression models.

### 2.6 Data Imputation and Normalization

We addressed missing values by the K-Nearest-Neighbour (KNN) imputation algorithm. This algorithm imputes missing values using values from the five nearest neighbours (i.e., the shortest Euclidean distance regarding the remaining predictors) that have a value for that variable, averaging these uniformly. For binary variables, after averaging, we mapped values < 0.5 to 0 and values ≥ 0.5 to 1. To accomplish this, we first normalized all variables in the train cohort and the data from the observational study, using centering and standard scaling. We fitted the imputer algorithm using the combined data of the train cohort with the observational study, and used it to fill in missing values in both the train and test cohorts. Subsequently, we transformed the imputed datasets back to their original scale. Lastly, we normalized the imputed train and test cohorts once again by centering and scaling each variable (ie, both continuous and binary variables) based on its standard deviation, ensuring that all variables in the training data are zero-mean and have unit variance before the model is trained.

### 2.7 Alternative Modelling Procedures

In our modelling procedure, several crucial choices were made: we opted for effect modelling instead of risk modelling and, as proposed by Tian and colleagues, we excluded main effects and the intercept term from the logistic regression model and encoded the treatment variable as ±1 (deviating from the more conventional 0/1 encoding).^8,24^ Furthermore, we utilized a LASSO penalty instead of alternative options, like a Ridge penalty. To demonstrate the impact of these choices on the resulting models and their performances, we conducted experiments with alternative modelling procedures resulting from different combinations of these choices (appendix D).

### 2.8 Addition of dichotomized variables

As our modelling approach only enables modelling of the variables as linear effects, we repeated our procedure (and all alternative procedures as outlined in the previous section), but with the introduction of additional dichotomized variables for each of the continuous variables used in the analysis. In particular, during step 2, after handling missing data through imputation, we introduced an extra variable for every continuous variable by splitting it based on the median value within the training cohort. For instance, apart from representing heart rate as a continuous variable, we also included a dichotomized variable, encoded as ‘1 if heart rate > 100, 0 otherwise,’ (in case of a median heart rate of 100 bpm in the training cohort). The remainder of the modelling procedure remained unchanged. We will refer to the modelling procedure without dichotomized variables as ‘procedure 1’ and with additional dichotomized variables as ‘procedure 2’.

### 2.9 Performance benchmark with pneumonia severity index

Under the assumption of homogeneous relative treatment effect of adjuvant treatment with corticosteroids among CAP patients, it is expected that patients with a higher initial risk of mortality would gain a greater absolute benefit from the treatment, a concept known as ‘risk magnification’.^25^ Assuming a homogeneous relative treatment effect, a model that accurately predicts the baseline mortality risk in CAP should be sufficient to identify patients who benefit more from adjuvant treatment with corticosteroids. Therefore, we benchmarked the discrimination for benefit of our proposed modelling procedures with the PSI, a well-established risk score for predicting adverse outcomes in CAP.^20^ For this score, we calculated the AUC-benefit in the same way as the other models, but using PSI thresholds instead of ITE thresholds. Assuming a decision threshold at a PSI score of 90, we divided patients in two groups (ie, class I–III vs. class IV-V), and compared the treatment effect in both groups and tested for HTE between these groups.

## 3. Results

### 3.1 Study and Patient Characteristics

Characteristics of the six included trials including a total of 1,896 patients are summarized in Table 1. Inclusion criteria were mostly based on the ATS/IDSA criteria for severe pneumonia, with some variations among the trials.^26^ Mortality rates varied among the trials (3.6-17.4%). The corticosteroid dose and duration varied, with a cumulative, hydrocortisone equivalent dose on day 7 ranging between approximately 500 and 2,000 mg (supplementary Figure F3). The maximum time between hospital admission and start of treatment ranged between 12 and 36 hours. In almost all included patients (97.5%), standard laboratory assessment was performed within 24 hours from hospital admission. Adherence to the study protocol was high in all the trials, with a ratio between the per-protocol and intention-to-treat population of 0.93. This ratio ranged between 0.86 and 1 among the included trials. The total 30-day mortality rate was 4.9%. Pooled baseline characteristics were similar between the corticosteroid and placebo group (Table 2). Baseline characteristic distributions were visualized per study and per treatment arm (supplementary Figure F4). Statistically significant (ie, a p value < 0.05) imbalance between the treatment arms (compared by the Fisher exact test for categorical variables and a two-sample t test for continuous variables, without adjusting for multiple testing) were observed with respect to urea levels in the trial by Torres et al.^18^, with respect to glucose levels by Blum et al.^16^, and with respect to CRP levels and occurrence of congestive heart failure in the trial by Snijders et al.^14^

**Table 1:**
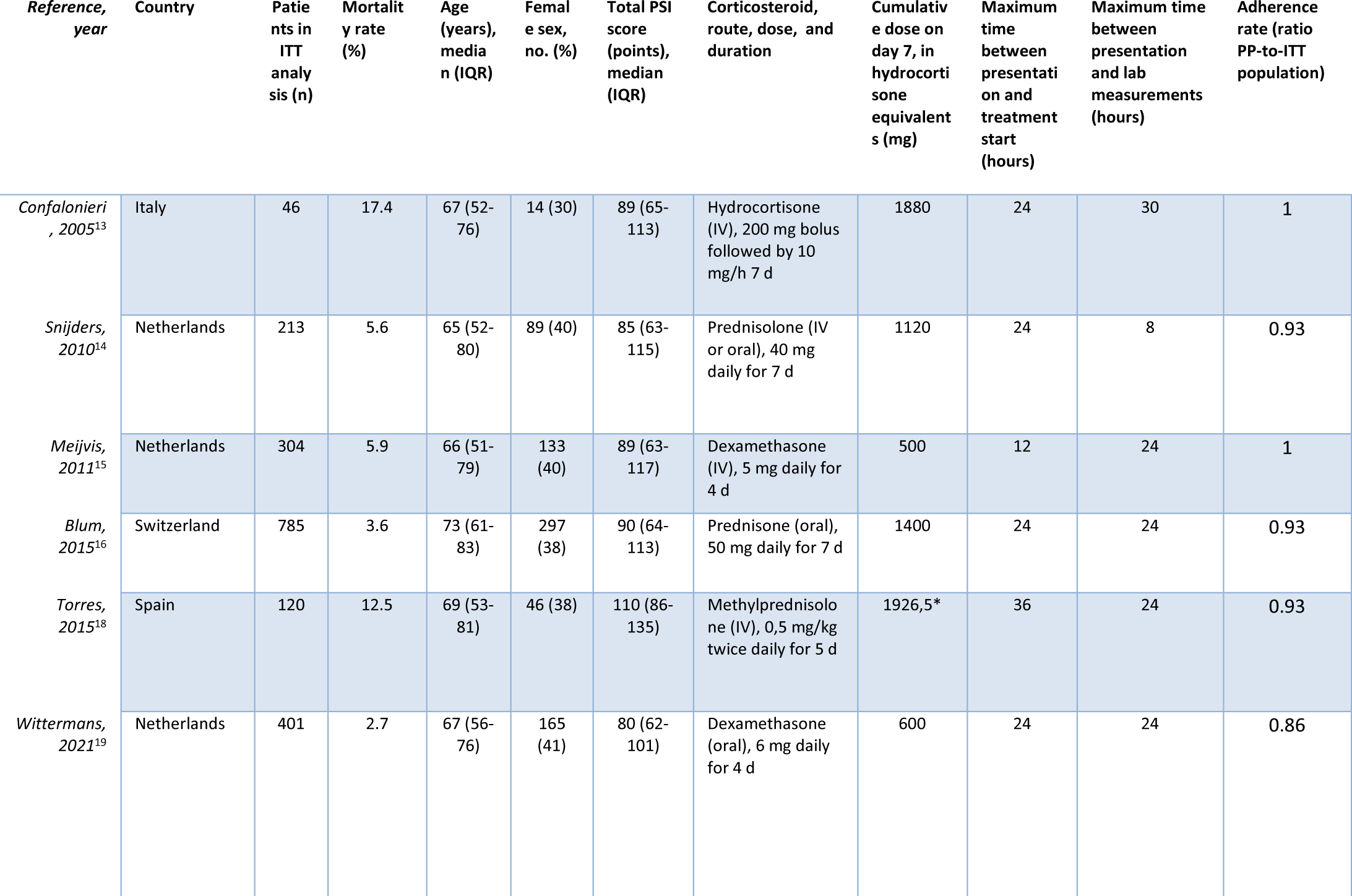
Characteristics of the included RCTs. IQR=interquartile range, NA=not available, PSI=pneumonia severity index, IV=intra-venous, ITT=intention-to-treat, PP=per-protocol. *To calculate the cumulative dose in the treatment regime of *Torres et al.*,^18^ which assigned patients in the treatment arm to 0.5 mg/kg per 12 hours of methylprednisolone, we assumed an average weight of 84 kg for male patients and 65.9 kg for female patients.^38^

**Table 2:** Baseline characteristics of the 1,869 patients from the included RCTs. Data are n (%) or median (IQR). *PSI values are missing for three (0.2%) patients, therefore the total numbers in the severity classes do not add up to total number of patients in treatment arm.

|  | Corticosteroid group<br>(N=934) | Placebo group<br>(N=935) |
| --- | --- | --- |
| **Demographics** |  |  |
| Female sex | 386 (41.3) | 359 (38.4) |
| Age, (years) | 70.0 (56.2-80.0) | 69.0 (55.0-80.0) |
| **Clinical parameters** |  |  |
| Resp. rate, (breaths/min) | 22.0 (18.0-27.0) | 22.0 (18.0-28.0) |
| Dias. blood pressure, (mmHg) | 70.0 (61.0-80.0) | 70.0 (61.0-80.0) |
| Syst. blood pressure, (mmHg) | 128.0 (112.0-142.0) | 126.0 (112.0-141.0) |
| Temperature, (°C) | 37.9 (37.2-38.7) | 38.0 (37.2-38.7) |
| Heart rate, (bpm) | 92.0 (80.0-106.0) | 92.0 (79.0-106.0) |
| SaO<sub>2</sub>, (%) | 94.0 (92.0-96.0) | 95.0 (92.0-97.0) |
| **Laboratory values** |  |  |
| Creatinine, (μmol/L) | 90.0 (70.7-120.2) | 89.0 (71.8-117.0) |
| Sodium, (mmol/L) | 136.0 (133.0-139.0) | 136.0 (133.0-138.0) |
| Urea, (mmol/L) | 6.8 (4.8-10.6) | 6.7 (4.7-9.8) |
| CRP, (mg/L) | 196.0 (98.0-300.0) | 188.1 (87.4-292.9) |
| Glucose, (mmol/L) | 7.1 (6.0-8.5) | 6.9 (6.0-8.4) |
| WBC count, (10<sup>9</sup> cells/L) | 12.8 (9.4-17.1) | 12.7 (9.1-16.9) |
| **Comorbidities** |  |  |
| Neoplastic disease | 59 (6.3) | 61 (6.5) |
| Liver disease | 22 (2.4) | 17 (1.8) |
| Congestive heart failure | 159 (17.0) | 152 (16.3) |
| Renal disease | 157 (16.8) | 139 (14.9) |
| Diabetes mellitus | 162 (17.3) | 176 (18.8) |
| Chronic obstructive pulmonary disease | 59 (6.3) | 61 (6.5) |
| **PSI\*** |  |  |
| Total score | 90.0 (64.0-115.0) | 87.0 (65.0-111.0) |
| Class I | 124 (13.3) | 114 (12.2) |
| Class II | 167 (17.9) | 159 (17.0) |
| Class III | 177 (19.0) | 224 (24.0) |
| Class IV | 335 (35.9) | 319 (34.2) |
| Class V | 130 (13.9) | 117 (12.5) |

### 3.2 Modelling Procedures

In the first step of both modelling procedures, some variables were excluded due to lack of availability in either the train or test cohort, depending on which trial formed the test cohort (supplementary Table F2).

The results of the first (wide) and second (fine) grid searches for λ optimization (ie, modelling step 5) in procedure 1 are depicted in supplementary Figure D1-o. The weights of the six trained models (step 6) in the different folds of the LOTO-CV procedure are visualized in supplementary Figure D3-x. The interaction term with CRP was selected in all LOTO-CV folds. For modelling procedure 2, the results of the grid searches and the weights of the corresponding trained models are depicted in supplementary Figure D2-o and D4-x, respectively. Here, the interaction term with CRP was selected in all LOTO-CV folds, and the interaction term with dichotomized glucose was selected in four of the six folds. Results of the grid searches and weight of the trained models for all alternative procedures (see section 2.7) are depicted in supplementary Figures D1-4.

### 3.3 Model Evaluation

#### 3.3.1 Discrimination for benefit

Overall, modelling procedure 2 yielded the highest discrimination for benefit, with an AUC-benefit of 184.9 (28.6 to 347.6, 95% CI), compared to 101.0 (−55.9 to 267.8, 95% CI) for procedure 1 and 118.0 (−55.1 to 299.1, 95% CI) for the PSI. Discriminative performances of the modelling procedure compared with the alternative procedures (see section 2.7) for procedure 1 and 2 are depicted in supplementary figures D5 and D6, respectively. Both procedure 1 and 2 yielded a higher AUC-benefit compared to all alternative procedures. Unlike procedure 1, procedure 2 also outperformed the PSI.

#### 3.3.2 Calibration for benefit

In both procedure 1 and 2, ITEs are underestimated in the lower ITE region and slightly overestimated ITEs in the higher ITE region (Figure 2). The underestimation in the lower ITE region was more pronounced in procedure 1 compared to procedure 2.

**Figure 2:**
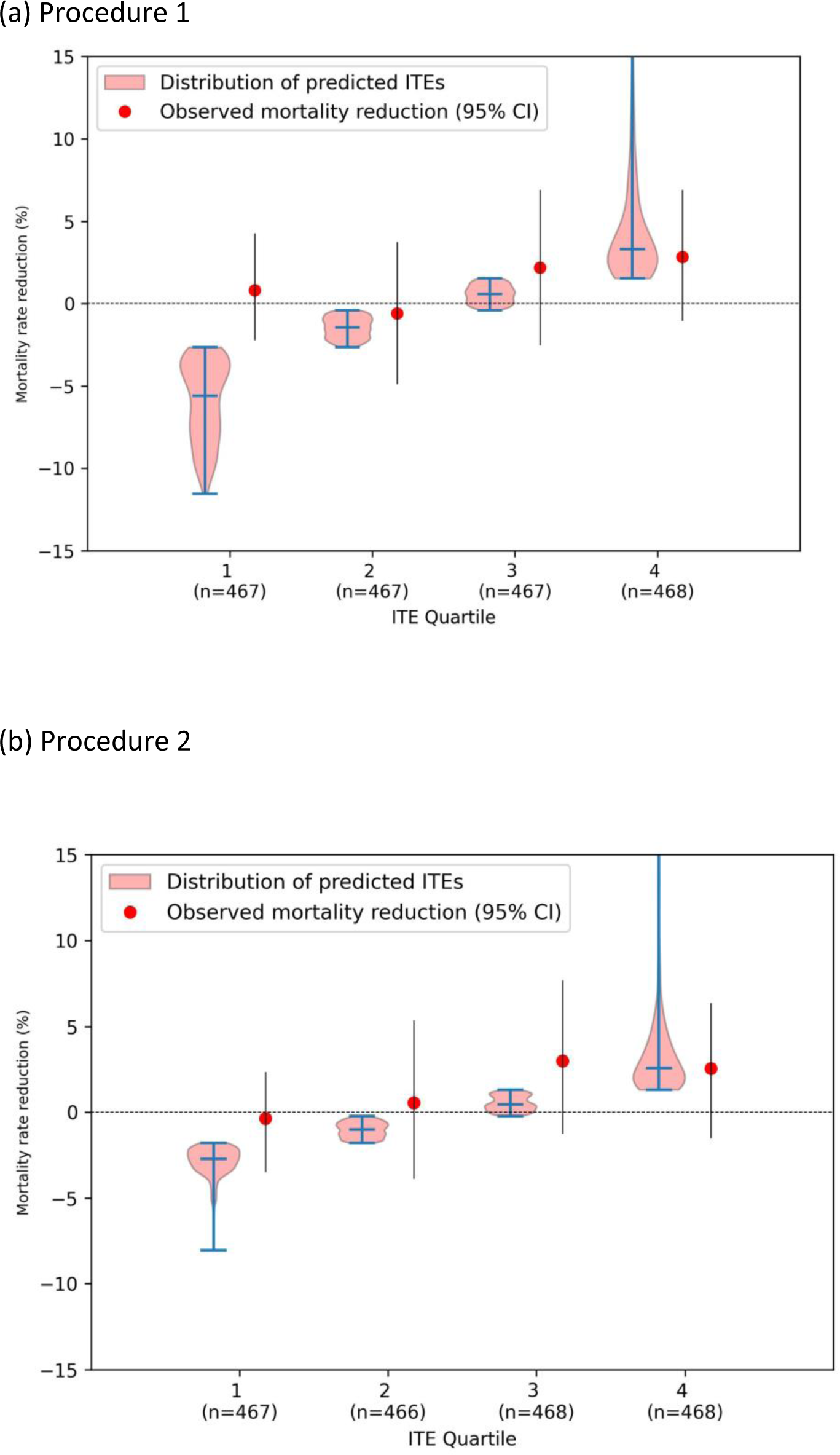
Calibration plots of the ITEs resulting from procedure 1 and 2. For four patient groups based on ascending ITE quartiles, the ITE distributions are using violin plots, next to the observed mortality reductions in each quartile.

#### 3.3.3 Predicted harm vs predicted benefit group

In the procedure that yielded the highest discrimination for benefit (ie, procedure 2), the models predicted harm (ie, ITE ≤ 0) in 1,004, and benefit (ie, ITE > 0) in 865 patients (Figure 3-b, Table 3). The observed benefit by adjuvant treatment with corticosteroids was greater for the predicted benefit group compared to the predicted harm group, with an odds ratio of 0.5 (0.3 to 0.9, 95% CI) versus 1.1 (0.7 to 1.8, 95% CI) and a mortality reduction of 3.2% (0.7 to 5.6, 95% CI) versus −0.3% (−2.6 to 1.8, 95% CI). Through the interaction test, we found a non-significant HTE between the predicted benefit and predicted harm group (p-value for interaction= 0.088). The NNT for the predicted benefit and predicted harm group were 31 and −293, respectively. These results indicate that, on average, 31 patients in the predicted benefit group must receive adjuvant treatment with corticosteroids to avert a single additional mortality case. Conversely, within the predicted harm group, the administration of corticosteroids to 293 patients, on average, is projected to cause one additional mortality case (Table 3).

**Figure 3:**
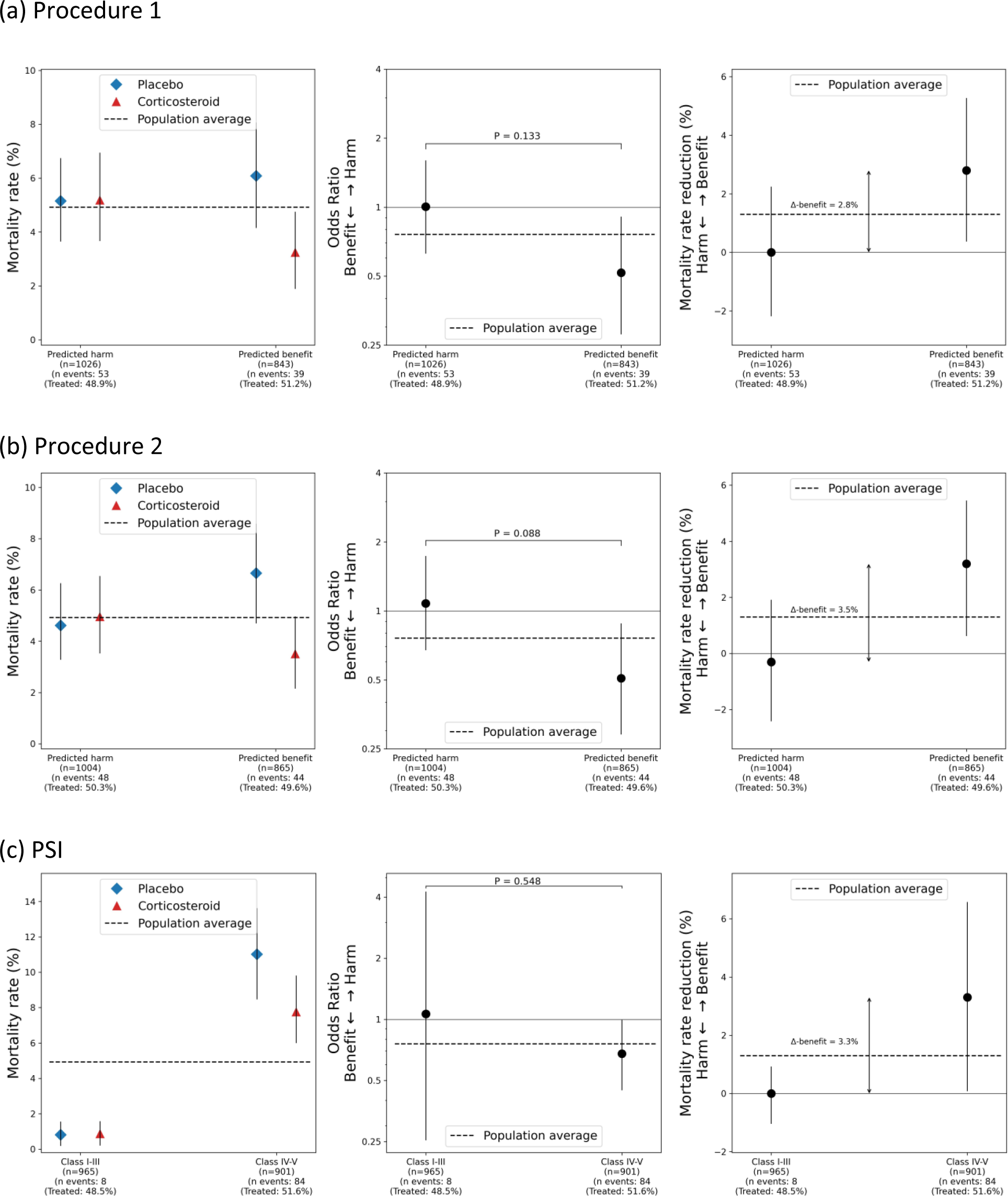
Observed treatment effects in the predicted harm group compared to the predicted benefit group resulting from procedure 1, procedure 2 and the PSI.^20^ Mortality rates (left) and treatment effect in terms of odds ratios (middle) and mortality reduction (right) are shown with 95% confidence intervals.

**Table 3:** Heterogeneity in treatment effect of corticosteroids among the subgroups identified in the LOTO-CV procedure by the PSI, and modelling procedures 1 and 2. OR=odds ratio, NNT=number of patients needed to treat. *Negative sign denotes net harm, hence representing the number of patients who need to be treated, on average, to cause 1 additional death.

|  | 30-day mortality rate |  | Relative effect (OR) | Mortality reduction (%) | NNT | P for Heterogeneity (Interaction) |
| --- | --- | --- | --- | --- | --- | --- |
|  | Placebo | Corticosteroid |  |  |  |  |
| **Overall** |  |  |  |  |  | - |
|  | 52/935 (5.6%) | 40/934 (4.3%) | 0.8 (0.5 to 1.1) | 1.3% (-0.4 to 2.7) | 78 |  |
| **PSI** |  |  |  |  |  | P = 0.548 |
| Class I-III (n=993) | 4/497 (0.8%) | 4/468 (0.9%) | 1.1 (0.2 to 4.6) | -0.0% (-1.1 to 1.0) | -2005 |  |
| Class IV-V (n=932) | 48/436 (11.0%) | 36/465 (7.7%) | 0.7 (0.5 to 1.0) | 3.3% (0.2 to 6.3) | 30 |  |
| **Procedure 1** |  |  |  |  |  | P = 0.113 |
| Predicted harm group (n=1,026) | 27/524 (5.2%) | 26/502 (5.2%) | 1.0 (0.7;1.6) | -0.0% (-2.3 to 1.9) | -3757 |  |
| Predicted benefit group (n=843) | 25/411 (6.1%) | 14/432 (3.2%) | 0.5 (0.3;0.9) | 2.8% (0.6 to 4.9) | 35 |  |
| **Procedure 2** |  |  |  |  |  | P = 0.088 |
| Predicted harm group (n=1,004) | 23/499 (4.6%) | 25/505 (5.0%) | 1.1 (0.7 to 1.8) | -0.3% (-2.6 to 1.8) | -293 |  |
| Predicted benefit group (n=865) | 29/436 (6.7%) | 15/429 (3.5%) | 0.5 (0.3 to 0.9) | 3.2% (0.7 to 5.6) | 31 |  |

These results for procedure 1 and the PSI can be found in Figure 3-a and 3-c, respectively, as well as in Table 3.

Supplementary Figures F5 and F6 display the results separately for each fold of the LOTO-CV in procedures 1 and 2, respectively. For both procedures, in most folds, HTE is observed in the expected direction (ie, more benefit is observed in the predicted benefit group compared to the predicted harm group), both in terms of odds ratio and mortality reduction. However, in both procedures, in the fold where the trial by Snijders et al.^14^ forms the test cohort, the model identified HTE in the opposite direction, with more benefit observed in the predicted harm group compared to the predicted benefit group, and vice versa.

#### 3.3.4 Group-defining variables

For a comprehensive view of the baseline characteristics disparity between the predicted harm and predicted benefit group identified in procedure 2, each continuous variable was standardized using z-scores relative to all 1,869 patients. Subsequently, we depicted the normalized distributions of each variable individually for both groups, arranged in ascending order of distinctiveness (Figure 4). Supplementary Table F3 presents the absolute differences in their original (non-normalized) values. Compared to the predicted harm group, the predicted benefit group are characterized by relatively high values for CRP (median CRP 299 vs 111 mg/L), glucose (median glucose 6.5 vs 7.6 mmol/L), and heart rate (median heart rate 88 vs 98 bpm), and relatively low values for sodium (median sodium 137 vs 134 mmol/L).

**Figure 4:**
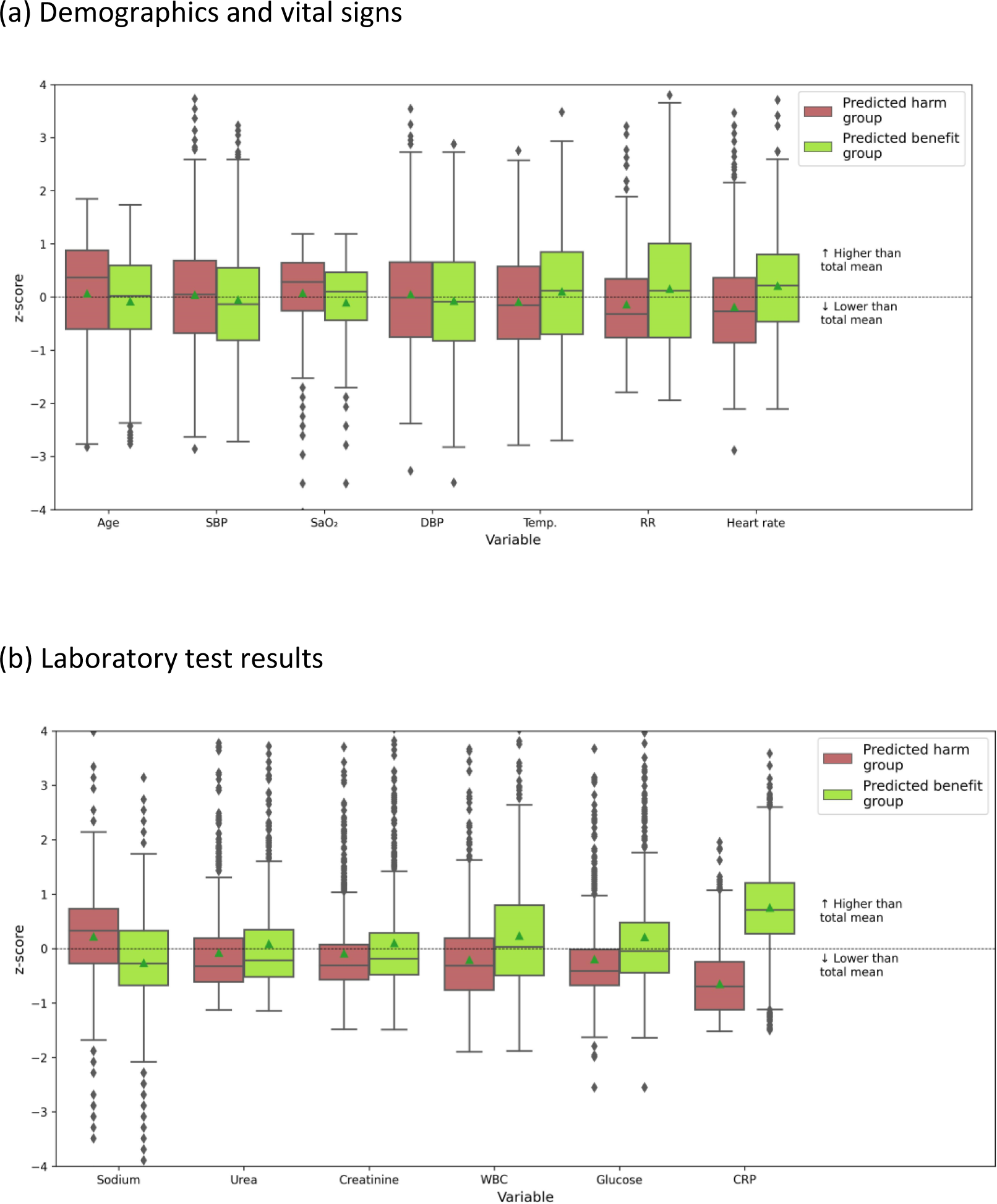
Normalized distributions of each continuous non-laboratory (a) and laboratory (b) variable between the predicted harm (red boxes) and predicted benefit group (green boxes), identified in procedure 2. The variables are sorted on the degree of separation between the groups. A value of +1 for the z-score (y-axis) would signify that the mean value for the subgroup was one standard deviation higher than the mean value in the cohort as a whole (ie, all 1,869 patients). SBP=systolic blood pressure, DBP=Diastolic blood pressure, WBC=White Cell Count, CRP=C-reactive protein. The green triangles represent the distribution means.

## 4. Discussion

### 4.1 Principal findings

Our findings suggest that the proposed modelling procedures are able to identify patients with CAP who would benefit from adjuvant treatment with corticosteroids based on their baseline characteristics, with procedure 2 leading to the best discriminative performance. In procedure 2, the interaction terms with CRP and dichotomized glucose were predominantly selected by the LASSO regression models, suggesting these being credible effect modifiers. Notably, despite the inclusion of various comorbidities, none of these were selected by the LASSO regression models in procedure 1 or 2. High glucose levels (i.e., > 7 mmol/L) were associated with greater benefit from adjuvant treatment with corticosteroids (Supplementary Figure F7-a). Although for CAP patients, elevated blood glucose level at baseline is associated with increased mortality,^27^ the association between elevated baseline blood glucose and more benefit from corticosteroids is surprising. Increased blood glucose may originate from several causes, such as increased hepatic gluconeogenesis and insulin resistance induced by inflammation^28^ or pre-existing diabetes mellitus (DM). Similar to the contrast between low and high baseline glucose levels, we also noted a greater advantage for patients with DM compared to those without DM (Supplementary Figure F7-b). This raises the question whether CAP patients with DM benefit more from adjuvant treatment with corticosteroids, and whether elevated baseline glucose levels merely serve as a proxy for identifying DM patients, or vice versa. Upon examining the treatment effects in patients with low (i.e., ≤ 7 mmol/L) and high (i.e., > 7 mmol/L) glucose levels, stratified by the presence of DM (supplementary Figure F7-c), we observed two conflicting phenomena. First, among patients without DM, a greater benefit was observed in those with high glucose levels compared to those with low glucose levels. Second, among patients with low glucose levels, a greater benefit was observed in those with DM compared to those without DM. Therefore, based on the currently available data, the observed HTE for corticosteroids cannot be solely attributed to glucose levels or the presence of DM. Further research, including data from the two most recent CAP trials,^11,12^ may provide deeper insights into this matter. High CRP levels being associated with treatment benefit, on the other hand, is less surprising. It has previously been hypothesized as an effect modifier by Briel et al.,^7^ and Torres et al.^18^ used CRP to ‘enrich’ the patient population in their trial.

### 4.2 Related works

In a previous IPDMA, Briel et al.^7^ noticed a tendency towards greater efficacy of corticosteroid treatment in patients with more severe CAP. However, the definition of ‘severe’ CAP lacks clarity. Briel et al.^7^ categorized CAP patients into severe and less severe groups based on univariate criteria, including CRP levels (<1 88 mg/L vs. ≥ 188 mg/L), initial admission to the intensive care unit (ICU), and Pneumonia Severity Index (PSI) (class I-III vs. class IV-V). However, they found no statistically significant effect modification. The two most recent trials investigating adjuvant treatment with corticosteroids in CAP only enrolled patients with severe CAP but yielded conflicting results.^11,12^ Meduri et al.^11^ selected patients with either one major or three minor modified ATS/IDSA criteria for severe pneumonia,^26^ whereas Dequin et al.^12^ used self-defined inclusion criteria, including the initiation of mechanical ventilation. Hence, the definition of severe CAP varies between studies, and some criteria used are based on (subjective) clinical decisions (such as the initiation of mechanical ventilation), leading to potential variation in classifying CAP severity across countries, hospitals, and physicians. In contrast, the predictive approach proposed here generates ITE predictions based on objective patient characteristics, providing an unambiguous way to identify the subgroup of CAP patients likely to benefit from adjuvant treatment with corticosteroids.

Wittermans et al.^29^ performed a post-hoc analysis on three trials,^15,16,19^ employing latent class modelling (LCA) to present two CAP ‘phenotypes’, with the ‘hyperinflammatory phenotype’ showing greater benefit in terms of length of hospital stay. However, we chose an effect modelling approach rather than LCA as it directly models the treatment effect, whereas in LCA, the model remains agnostic to treatment effects.

### 4.3 Modelling procedure

#### 4.3.1 Risk vs effect modelling

Risk modelling is generally preferred over effect modelling, as the latter is prone to overfitting and could lead to treatment mistargeting.^8,30^ Risk modelling relies on the concept of ‘risk magnification’, where patients with higher baseline mortality risk may experience a greater absolute treatment effect, even when the relative treatment effect is constant for all patients.^25^ It uses a multivariable model to predict outcome risk and stratifies patients based on their risk level to explore variations in treatment effects. In this study, risk modelling identified age, respiratory rate (RR), and urea as important predictors of mortality in patients with CAP (supplementary Figure D3-a and D4-a), which also play a big role in traditional CAP severity scores (ie, the PSI^20^ and the CURB-65^31^). However, the AUC-benefit obtained from risk modelling was only slightly higher than the AUC-benefit obtained from randomly generated ITEs (supplementary Figure D6), suggesting a non-constant relative treatment effect and therefore supporting the use of effect modelling. In contrast to the risk models, the PSI^20^ (in essence also a risk model) did show to be useful to identify HTE in CAP patients, with higher PSI scores (ie, class IV-V) showing more benefit from adjuvant treatment with corticosteroids in terms of absolute mortality risk reduction (Figure 3-c). This discrepancy may be explained by information incorporated in the PSI,^20^ but not in our risk modelling procedures (such as arterial pH or the presence of pleural effusion), although further research is needed in this regard.

#### 4.3.2 Influence of modelling choices

The effect modelling method we utilized, as proposed by Tian et al.^24^, is a variant of penalized logistic regression, in which some very specific choices are made: LASSO (opposed to Ridge) penalization, exclusion of an intercept term and main effects, and ±1 encoding of the treatment variable. Variations in these choices led to notably different model weights (appendix D). For example, the only difference between the ‘effect-7’ and ‘effect-8’ models (supplementary Table D1) is the encoding of the treatment variable (ie, 0/1 vs ±1). For the effect-7 model, the Lasso penalization primarily selected interaction terms with age, RR, and urea, while interaction terms with CRP and glucose were completely eliminated across all LOTO-CV folds (supplementary Figure D4-u). In contrast, in our proposed modelling procedure (i.e., effect-8), the Lasso penalization predominantly selected interaction terms with CRP and glucose, while interaction terms with age and RR were completely eliminated across all LOTO-CV folds (supplementary Figure D4-x). For a deeper understanding of how these modelling choices influence the model fitting, further research is needed.

#### 4.3.3 Credibility of effect modifiers

Considering the risk of overfitting with effect modelling, it is recommended to assess the credibility of treatment-variable interaction terms using comprehensive multidimensional criteria, such as whether the a priori hypothesized direction of (relative) effect modification for specific interactions matches the observed direction.^8,32^ Relative effect modification, however, can be a challenging concept to comprehend, necessitating expertise in both clinical and statistical domains for accurate judgment. In light of this, we chose a fully data-driven effect modelling approach, allowing the model to select interactions independent of subjective and potentially wrongly substantiated human judgements.

#### 4.3.4 Variation in performance of unpenalized models

In both modelling procedures with and without dichotomized variables, we observed large variations of the performance of the different LOTO folds considering (almost) no penalization (Supplementary Figures D1 and D2). For instance, in our modelling procedure (ie, ‘Effect-8 LASSO) with dichotomized variables, in the fold where the Wittermans trial^19^ forms the test cohort, the AUC-benefit is much higher in the small λ region compared to the other folds (supplementary Figure D2-o). Further research is warranted to understand the mechanisms behind these large variations in performance for unpenalized models in specific train and test cohort combinations.

### 4.4 Model evaluation

Van Klaveren et al.^33^ introduced the ‘c-for-benefit’ metric for evaluating the discriminative performance of ITE modelling. However, this metric requires one-to-one patient matching, which remains a challenging task. To avoid biasing our model towards a specific decision threshold, we did not adopt the ‘population benefit’ metric proposed by Efthimiou et al.,^22^ which uses a decision threshold on the absolute ITE scale to group patients. Instead, we introduced the AUC-benefit metric, which quantifies the discriminative performance of the model without relying on one-to-one matching or a fixed decision threshold. This metric is closely related to the (area under the) ‘Qini’ or ‘Uplift’ curve, as the Δ-benefit curve is a special case of the Qini/Uplift curve where treated and untreated patients are ranked jointly and the volumes are expressed in relative numbers (ie, percentiles).^34,35^

Finally, we also presented the results assuming a decision threshold of 0, as we argue that treatment would be beneficial in any patient where a reduction in mortality is expected. However, a nonzero decision threshold may also be justified, considering the potential side effects associated with adjuvant treatment with corticosteroids.

### 4.5 Study strengths and limitations

The design of our modelling procedure ensured that no information from test sets was used to influence the modelling choices, thus minimizing the risk of overfitting. The presented model’s ITE predictions offer an unambiguous way to divide patients into subgroup which will likely benefit from corticosteroids in terms of reduced mortality, as the model relies on objective patient measurements. Our ‘final models’ to be prospectively validated (see section 5) utilize only one (ie, CRP, final model 1) or two (ie, CRP, and glucose, Final model 3) measurements, which are typically routinely taken at hospital admission, meaning that no extra measurements would be required if the models would be used in clinical practice.

Our study has limitations. First, the proposed modelling procedures predicted benefit for patients where harm was observed and vice versa in the fold where the trial by Snijders et al.^14^ formed the test cohort. Nevertheless, given the small sample sizes of (most) individual trials, it is hard to draw definitive conclusions on model performance in individual trials. For the same reason, we opted to pool ITEs predicted by different models (from the different LOTO-CV folds) to evaluate each modelling procedure in terms of discrimination and calibration for benefit, rather than evaluating the performance in individual trials. Second, the included trials show some notable differences in terms of treatment dose and duration among potential other differences, which make the pooling of all these trials questionable. Finally, other patient information may have improved ITE prediction, such as chest X-ray examinations, but were not available for this study. Cytokine data was available for some of the included trials, but these data were found to be incomparable between trials due to differences in used quantification techniques.

### 4.6 Conclusion

Our modelling approach has potential to identify patients with CAP, at hospital admission, who would benefit from the investigated adjuvant treatment with corticosteroids and those who would not. We will prospectively validate the proposed modelling procedures in the two most recent trials,^11,12^ as described in the following section.

## 5. Protocol for prospective validation

### 5.1 Training the final models

To minimize the risk of overfitting, our modelling procedure was designed to prevent the use of any information of the test cohort to train the models. However, in order to determine the best-performing modelling procedure among various alternative procedures (as outlined in appendix D), we selected the one that yielded the best overall performance in the six included trials. Hence, risk of overfitting due to modelling procedure selection based on already available data, ie, the six included trials, still exists. To address this, we repeated procedure 1 and 2 (ie, with and without dichotomized variables) until step 6 (supplementary Figure F1), in which data from all six trials formed the train cohort (see appendix E). The ‘final models’ resulting from these folds will be prospectively validated in the trials conducted by Meduri et al.^11^ and Dequin et al. (ie, together forming the test cohort),^12^ whose IPD were not available to the authors at the time of selecting the modelling procedure was chosen and the final models were trained.

The final model resulting from the procedure 1 (ie, without dichotomized variables), referred to as ‘final model 1’, resulted in a model with only one non-zero weight for the interaction term with CRP (supplementary Figure E2 and Table E1). The final model resulting from the procedure 2 (ie, with dichotomized variables), referred to as ‘final model 2’, resulted in a model with four non-zero weights: for the interaction terms with CRP, dichotomized glucose (ie, glucose > 7 mmol/L), creatinine and sex (supplementary Figure E4 and Table E2). As models that require fewer variables are preferred in clinical practise, we fitted an extra final model (ie, ‘final model 3’), by again repeating procedure 2 until step 6 where all six trials formed the train cohort, but with an extra constraint. Namely, instead of selecting the optimal λ, we selected the optimal λ among λs which resulted in a final model with maximally two non-zero weights (supplementary Figure E3). This resulted in a model with two non-zero weights for the interactions with CRP and dichotomized glucose (ie, glucose > 7 mmol/L, supplementary Figure E5). To test the effect of this extra constraint on model performance, we also repeated the full LOTO-CV of procedure 2, using this constraint in each fold. This led to similar results in terms of AUC-benefit as in procedure 2 without this constraint (supplementary Figure F8).

Assuming a decision threshold (ie, an ITE value above which treating patients is considered worthwhile) of 0, final models 1 and 3 simplify to one or two (absolute) CRP thresholds (see appendix E). As such, we presented final models 1 and 3, assuming a decision threshold of 0, as simple flow charts (Figure 5).

**Figure 5:**
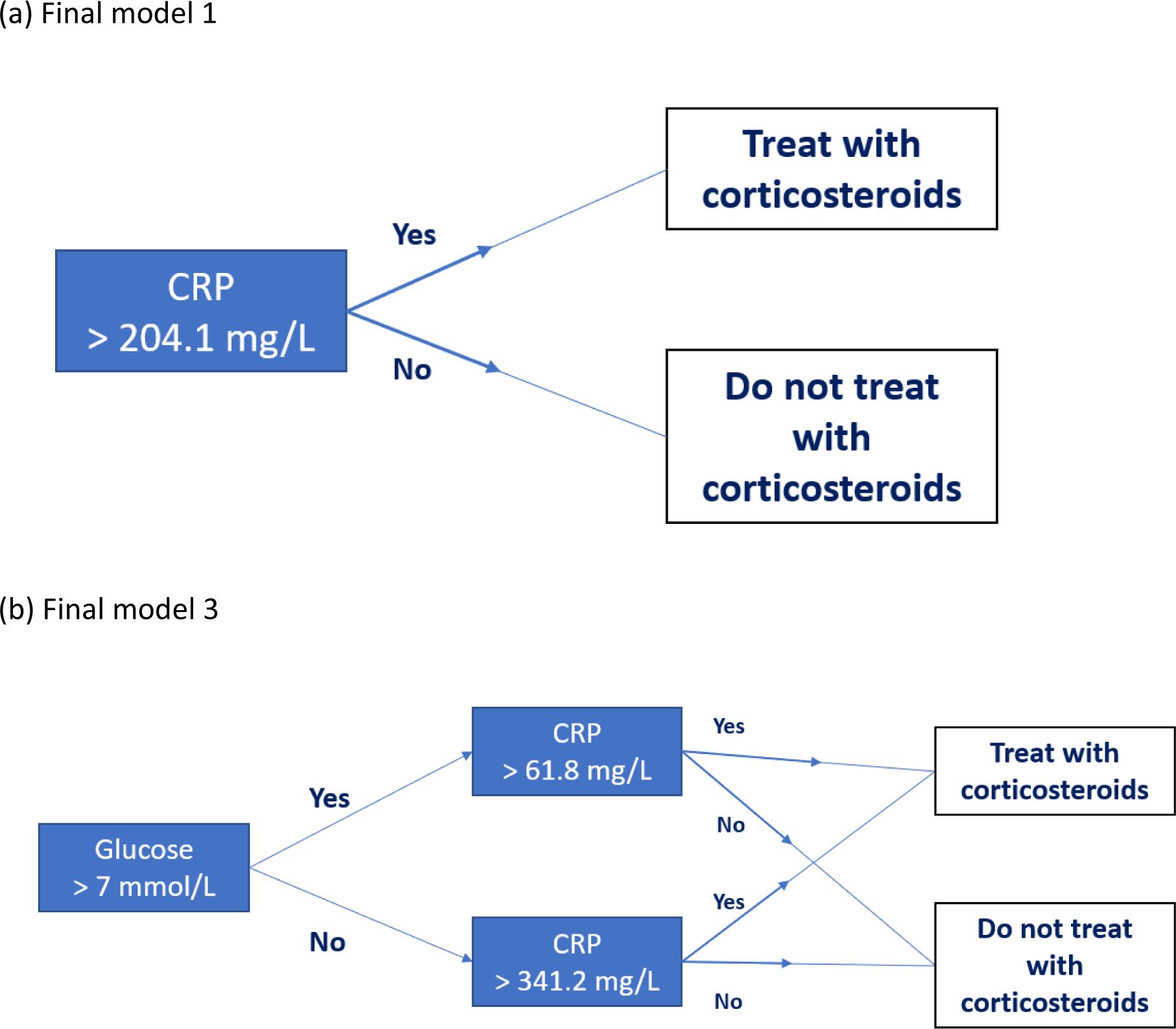
Simple flowcharts representing final models 1 and 3, assuming a decision threshold of 0.

The resulting final models, as well as the corresponding trained imputers and scalers are available on Github.^36^

### 5.2 Overview of prospective validation

Using data from the two recent trials,^11,12^ we will prospectively validate:

- Final model 1 (as depicted in supplementary Figure E2 and Table E1)
- Final model 2 (as depicted in supplementary Figure E4 and Table E2)
- Final model 3 (as depicted in supplementary Figure E5 and Table E3)
- the PSI.^20^

The performance of these models will be evaluated:

- in terms of discrimination for benefit, using the AUC-benefit (as described in section 2.3.1)
- in terms of calibration for benefit (as described in section 2.3.2)
- by splitting the patients in two groups (ITE ≤ 0 vs ITE > 0 for Final models 1-3, and Class I-III vs IV-V for the PSI). For these groups, we will present observed mortality rates in the treatment arms, observed treatment effects in terms of odds ratios and mortality reduction, and the NNT. And we will test for HTE between the groups using an interaction test by fitting a logistic regression model for mortality, using group assignment, treatment assignment, and their interaction as covariates (as described in section 2.3.3). Please note that, for final models 1 and 3, this evaluation is exactly the evaluation of the corresponding flowcharts (Figure 5).

### 5.3 Results in training data

For now, we evaluated the performance of the final models using the data on which the models were trained (ie, the six included trials). Final model 1, 2 and 3 yielded an AUC-benefit of 137.4 (−25.6 to 303.4, 95% CI), 252.0 (69.7 to 429.9, 95% CI) and 254.9 (80.6 to 426.9, 95% CI), respectively. Performances in terms of calibration for benefit for the final models are depicted in Figure 6. Comparisons between the predicted harm and predicted benefit groups identified by the final models, assuming a decision threshold of 0, are depicted in Figure 7 and Table 4. Depending on the extend of overfitting, these performances are expected to deteriorate to a certain extent when evaluated using the unseen data in the prospective validation.

**Figure 6:**
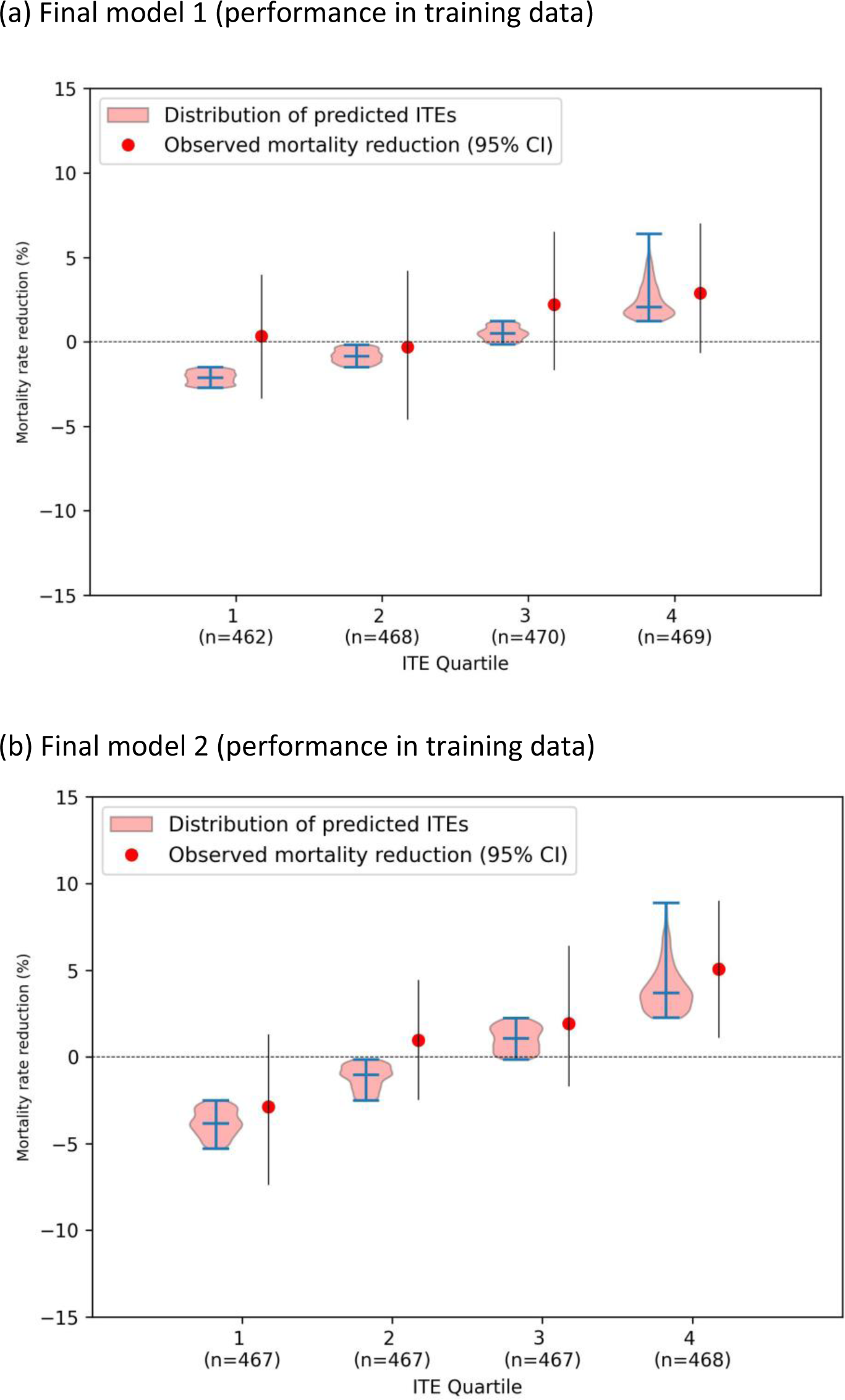
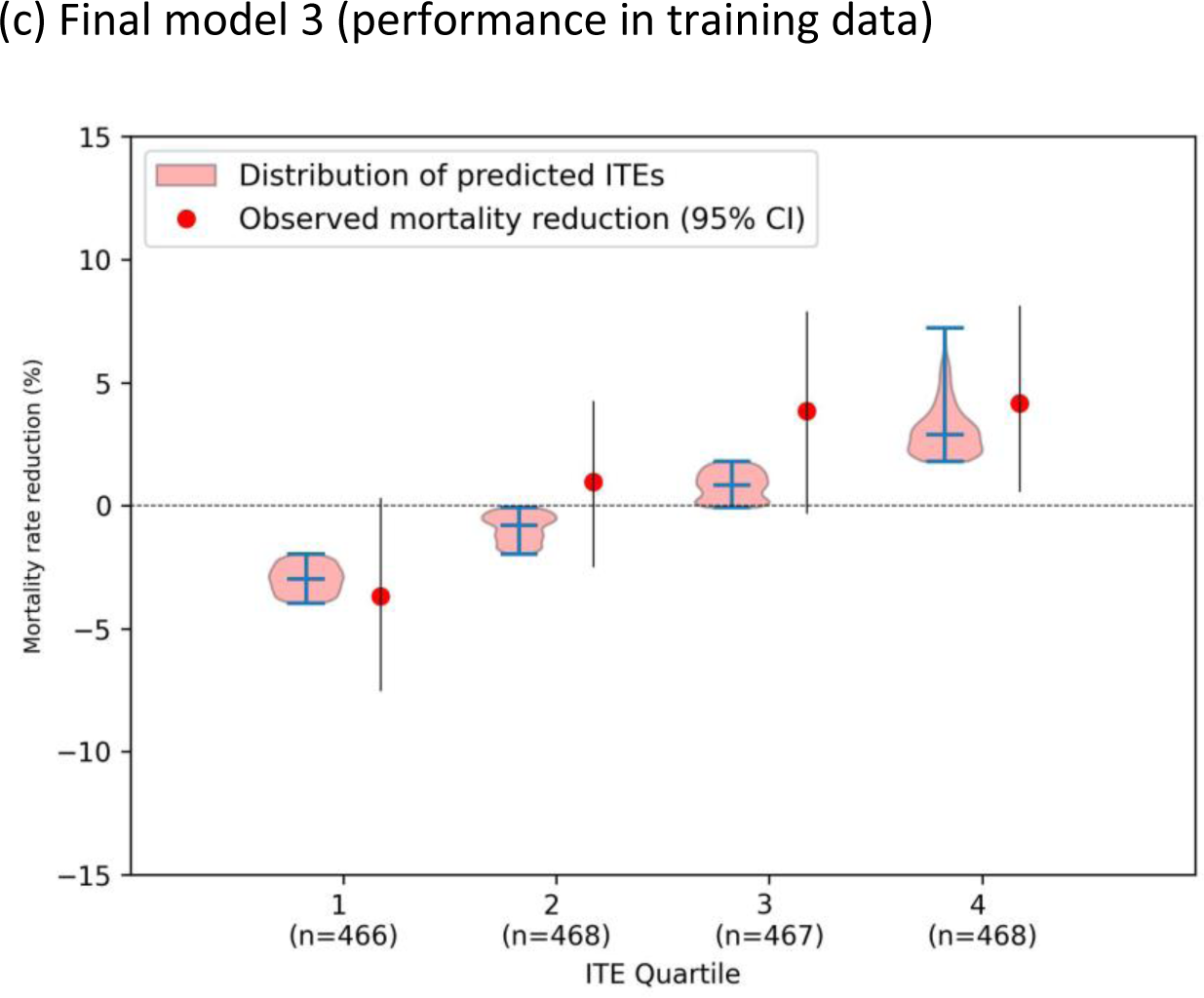
Calibration plots of the ITEs resulting from the ITEs predicted by the final models in the training data. For four patient groups based on ascending ITE quartiles, the ITE distributions are using violin plots, next to the observed mortality reductions in each quartile.

**Figure 7:**
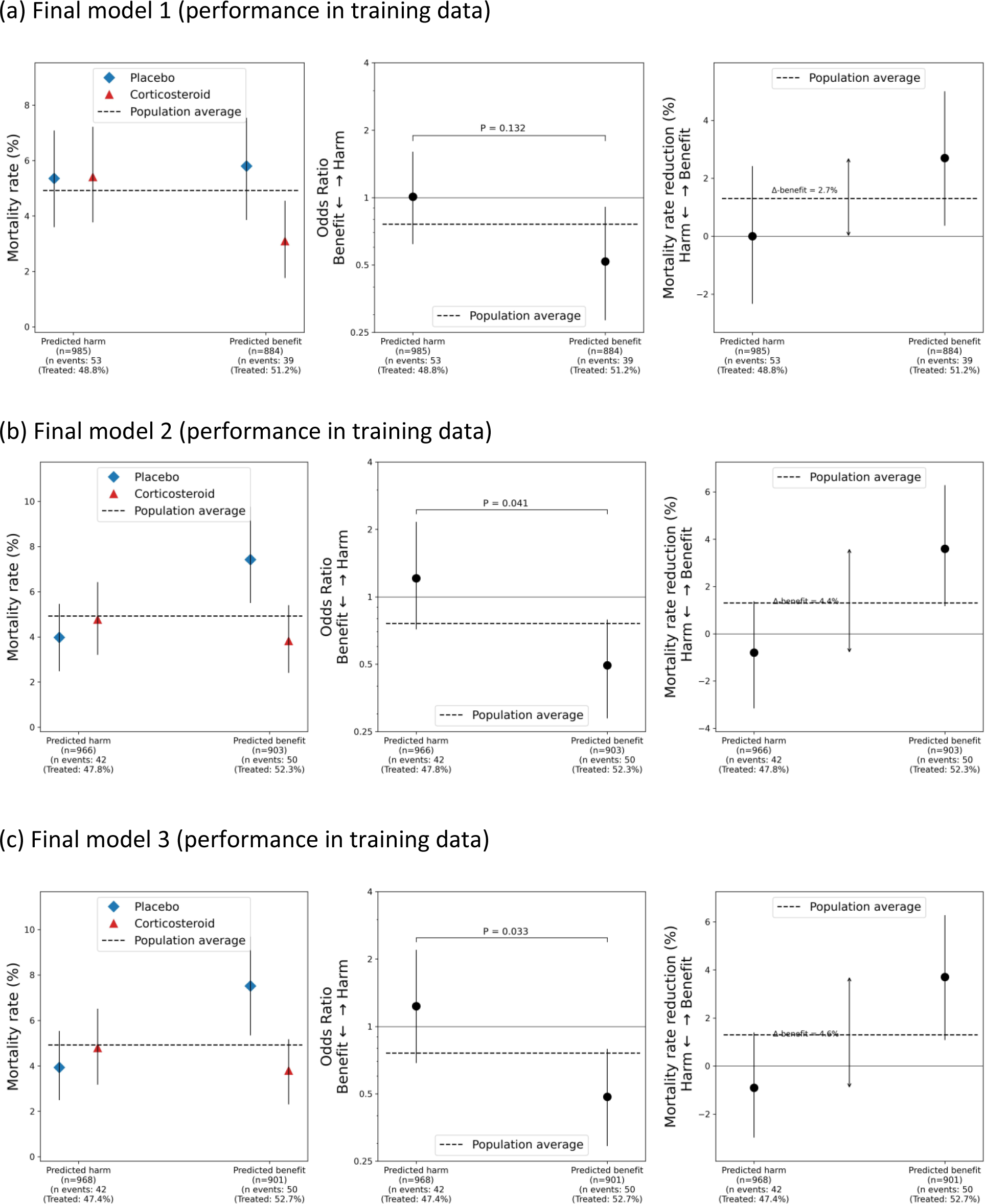
Observed treatment effects in the predicted harm group compared to the predicted benefit group resulting from the ITEs predicted by the final models in the training data. Mortality rates (left) and treatment effect in terms of odds ratios (middle) and mortality reduction (right) are shown with 95% confidence intervals.

**Table 4:** Heterogeneity in treatment effect of corticosteroids among the subgroups identified by the final models in the data on which these were trained. OR=odds ratio, NNT=number of patients needed to treat. *Negative sign denotes net harm, hence representing the number of patients who need to be treated, on average, to cause 1 additional death.

|  | 30-day mortality rate |  | Relative effect (OR) | Mortality reduction (%) | NNT | P for Heterogeneity (Interaction) |
| --- | --- | --- | --- | --- | --- | --- |
|  | Placebo | Corticosteroid |  |  |  |  |
| **Final model 1** |  |  |  |  |  | P = 0.132 |
| *Predicted harm group (n=985)* | 27/504 (5.4%) | 26/481 (5.4%) | 1.0 (0.6 to 1.6) | -0.0% (-2.4 to 2.4) | -2072 |  |
| *Predicted benefit group (n=884)* | 25/431 (5.8%) | 14/453 (3.1%) | 0.5 (0.3 to 0.9) | 2.7% (0.3 to 4.9) | 36 |  |
| **Final model 2** |  |  |  |  |  | P = 0.041 |
| *Predicted harm group (n=966)* | 20/504 (4.0%) | 22/462 (4.8%) | 1.2 (0.7 to 2.0) | -0.8% (-3.0 to 1.5) | -126 |  |
| *Predicted benefit group (n=903)* | 32/431 (7.4%) | 18/472 (3.8%) | 0.5 (0.3 to 0.8) | 3.6% (0.9 to 6.0) | 27 |  |
| **Final model 3** |  |  |  |  |  | P = 0.033 |
| *Predicted harm group (n=968)* | 20/509 (3.9%) | 22/459 (4.8%) | 1.2 (0.7 to 2.1) | -0.9% (-3.1 to 1.5) | -115 |  |
| *Predicted benefit group (n=901)* | 32/426 (7.5%) | 18/475 (3.8%) | 0.5 (0.3 to 0.8) | 3.7% (1.2 to 6.4) | 26 |  |

### 5.4 Hypothesis for effect modifiers

Based on our results, we hypothesize CRP and dichotomized glucose (ie, ≤ 7 mmol/L vs > 7 mmol/L) to act as relative effect modifiers. In contrast, we attribute the observed HTE between PSI class I-III and IV-V on the absolute scale (Figure 3-c) to risk magnification.^25^ And hence, we hypothesize PSI not to be a relative effect modifier. Once the data of the two recent trials^11,12^ is collected, we will test these hypotheses by testing for interaction by CRP, dichotomized glucose, and the PSI score using interaction tests. That is, we will fit a mixed effects logistic regression model (with random intercepts and slopes per trial) for mortality, using the variable of interest (ie, either CRP, dichotomized glucose or the PSI), treatment assignment, and their interaction as covariates, using data of all eight trials.^11–16,18,19^

### 5.5 Extra validation in other trials

Finally, two trials were judged ineligible for this review, although we still consider extra validation of our models in these trials useful. The trial by Fernandez-Serrano et al.^17^ was excluded because of the high used dose of corticosteroids (supplementary Figure F3). We excluded the trial by Lloyd et al.^37^ because they randomized a bundle of treatments, among which adjuvant corticosteroids therapy, which does not allow for the identification of the isolated effect of corticosteroids on mortality. As these trials are both close to eligible, we will also validate the final models and decision rules in these trials as an extra analysis.

## Data Availability

The proposed model (and corresponding dependencies), as well as the supplementary material, are available online on Github: https://github.com/jimmsmit/HTE_CAP. Individual participant data that underlie the results reported in this article are not publicly available. Requests should be directed to the corresonding authors of the included trials; data requestors will need to sign a data access agreement.

https://github.com/jimmsmit/HTE_CAP/tree/main

## References

1 Martin-Loeches I, Torres A. New guidelines for severe community-acquired pneumonia. Curr Opin Pulm Med 2021; 27: 210–5.

2 Global burden of 369 diseases and injuries in 204 countries and territories, 1990-2019: a systematic analysis for the Global Burden of Disease Study 2019. Lancet (London, England) 2020; 396: 1204–22.

3 Kellum JA, Kong L, Fink MP, et al. Understanding the inflammatory cytokine response in pneumonia and sepsis: results of the Genetic and Inflammatory Markers of Sepsis (GenIMS) Study. Arch Intern Med 2007; 167: 1655–63.

4 Remmelts HHF, Meijvis SCA, Biesma DH, et al. Dexamethasone downregulates the systemic cytokine response in patients with community-acquired pneumonia. Clin Vaccine Immunol 2012; 19: 1532–8.

5 Pitre T, Abdali D, Chaudhuri D, et al. Corticosteroids in Community-Acquired Bacterial Pneumonia: a Systematic Review, Pairwise and Dose-Response Meta-Analysis. J Gen Intern Med 2023. DOI:10.1007/s11606-023-08203-6.

6 Metlay JP, Waterer GW, Long AC, et al. Diagnosis and treatment of adults with community-acquired pneumonia. Am J Respir Crit Care Med 2019; 200: E45–67.

7 Briel M, Spoorenberg SMC, Snijders D, et al. Corticosteroids in Patients Hospitalized with Community-Acquired Pneumonia: Systematic Review and Individual Patient Data Metaanalysis. Clin Infect Dis 2018; 66: 346–54.

8 Kent DM, Paulus JK, Van Klaveren D, et al. The Predictive Approaches to Treatment effect Heterogeneity (PATH) statement. Ann Intern Med 2020; 172: 35–45.

9 Hoogland J, IntHout J, Belias M, et al. A tutorial on individualized treatment effect prediction from randomized trials with a binary endpoint. Stat Med 2021; 40: 5961–81.

10 Riley RD, Debray TPA, Fisher D, et al. Individual participant data meta-analysis to examine interactions between treatment effect and participant-level covariates: Statistical recommendations for conduct and planning. Stat Med 2020; 39: 2115–37.

11 Meduri GU, Shih M-C, Bridges L, et al. Low-dose methylprednisolone treatment in critically ill patients with severe community-acquired pneumonia. Intensive Care Med 2022; 48: 1009–23.

12 Dequin P-F, Meziani F, Quenot J-P, et al. Hydrocortisone in Severe Community-Acquired Pneumonia. N Engl J Med 2023; 388: 1931–41.

13 Confalonieri M, Urbino R, Potena A, et al. Hydrocortisone infusion for severe community-acquired pneumonia: A preliminary randomized study. Am J Respir Crit Care Med 2005; 171: 242–8.

14 Snijders D, Daniels JMA, De Graaff CS, Van Der Werf TS, Boersma WG. Efficacy of corticosteroids in community-acquired pneumonia: A randomized double-blinded clinical trial. Am J Respir Crit Care Med 2010; 181: 975–82.

15 Meijvis SCA, Hardeman H, Remmelts HHF, et al. Dexamethasone and length of hospital stay in patients with community-acquired pneumonia: A randomised, double-blind, placebo-controlled trial. Lancet 2011; 377: 2023–30.

16 Blum CA, Nigro N, Briel M, et al. Adjunct prednisone therapy for patients with community-acquired pneumonia: A multicentre, double-blind, randomised, placebo-controlled trial. Lancet 2015; 385: 1511–8.

17 Fernández-Serrano S, Dorca J, Garcia-Vidal C, et al. Effect of corticosteroids on the clinical course of community-acquired pneumonia: A randomized controlled trial. Crit Care 2011; 15. DOI:10.1186/cc10103.

18 Torres A, Sibila O, Ferrer M, et al. Effect of corticosteroids on treatment failure among hospitalized patients with severe community-acquired pneumonia and high inflammatory response: A randomized clinical trial. JAMA 2015; 313: 677–86.

19 Wittermans E, Vestjens SMT, Spoorenberg SMC, et al. Adjunctive treatment with oral dexamethasone in non-ICU patients hospitalised with community-acquired pneumonia: A randomised clinical trial. Eur Respir J 2021; 58: 1–10.

20 Fine MJ, Auble TE, Yealy DM, et al. A Prediction Rule to Identify Low-Risk Patients with Community-Acquired Pneumonia. N Engl J Med 1997; 336: 243–50.

21 Endeman H, Meijvis SCA, Rijkers GT, et al. Systemic cytokine response in patients with community-acquired pneumonia. Eur Respir J 2011; 37: 1431–8.

22 Efthimiou O, Hoogland J, Debray TPA, et al. Measuring the performance of prediction models to personalize treatment choice. Stat Med 2023;: 1188–206.

23 Holland PW. Statistics and Causal Inference. J Am Stat Assoc 1986; 81: 945–60.

24 Tian L, Alizadeh AA, Gentles AJ, Tibshirani R. A Simple Method for Estimating Interactions between a Treatment and a Large Number of Covariates. J Am Stat Assoc 2014; 109: 1517–32.

25 Harrell F. Viewpoints on Heterogeneity of Treatment Effect and Precision Medicine. 2018. https://www.fharrell.com/post/hteview/.

26 Mandell LA, Wunderink RG, Anzueto A, et al. Infectious Diseases Society of America/American Thoracic Society consensus guidelines on the management of community-acquired pneumonia in adults. Clin Infect Dis 2007; 44: S27–72.

27 Barmanray RD, Cheuk N, Fourlanos S, Greenberg PB, Colman PG, Worth LJ. In-hospital hyperglycemia but not diabetes mellitus alone is associated with increased in-hospital mortality in community-acquired pneumonia (CAP): a systematic review and meta-analysis of observational studies prior to COVID-19. BMJ open diabetes Res care 2022; 10: 1–9.

28 Baker EH, Archer JRH, Srivastava SA. Hyperglycemia, Lung Infection, and Inflammation. Clin Pulm Med 2009; 16: 258–64.

29 Wittermans E, van der Zee PA, Qi H, et al. Community-acquired pneumonia subgroups and differential response to corticosteroids: a secondary analysis of controlled studies. ERJ Open Res 2022; 8: 00489–2021.

30 van Klaveren D, Balan TA, Steyerberg EW, Kent DM. Models with interactions overestimated heterogeneity of treatment effects and were prone to treatment mistargeting. J Clin Epidemiol 2019; 114: 72–83.

31 Lim WS, Van Der Eerden MM, Laing R, et al. Defining community acquired pneumonia severity on presentation to hospital: An international derivation and validation study. Thorax 2003; 58: 377–82.

32 Schandelmaier S, Briel M, Varadhan R, et al. Development of the Instrument to assess the Credibility of Effect Modification Analyses (ICEMAN) in randomized controlled trials and meta-analyses. Can Med Assoc J 2020; 192: E901–6.

33 van Klaveren D, Steyerberg EW, Serruys PW, Kent DM. The proposed ‘concordance-statistic for benefit’ provided a useful metric when modeling heterogeneous treatment effects. J Clin Epidemiol 2018; 94: 59–68.

34 Radcliffe N. Using control groups to target on predicted lift: Building and assessing uplift model. Direct Mark Anal J 2007;: 14–21.

35 Devriendt F, Van Belle J, Guns T, Verbeke W. Learning to Rank for Uplift Modeling. IEEE Trans Knowl Data Eng 2020. DOI:10.1109/TKDE.2020.3048510.

36 Smit J. Predicting individualized treatment effects of corticosteroids in community-acquired-pneumonia. https://github.com/jimmsmit/HTE_CAP.

37 Lloyd M, Karahalios A, Janus E, et al. Effectiveness of a Bundled Intervention Including Adjunctive Corticosteroids on Outcomes of Hospitalized Patients with Community-Acquired Pneumonia: A Stepped-Wedge Randomized Clinical Trial. JAMA Intern Med 2019; 179: 1052–60.

38 Average sizes of men and women. https://www.worlddata.info/average-bodyheight.php.

