## Supplementary material for "Predicting individualized treatment effects of corticosteroids in community-acquired-pneumonia: a data-driven analysis of randomized controlled trials"

[1] Department of Intensive Care, Erasmus University Medical Center, Rotterdam, The Netherlands
[2] Pattern Recognition & Bioinformatics group, Delft University of Technology, Delft, The Netherlands
[3] Department of Pulmonary Medicine, Erasmus University Medical Center, The Netherlands
[4] Department of Pulmonary Medicine, Spaarne Gasthuis, Haarlem, The Netherlands
[5] Department of Pulmonary Medicine, Noordwest Hospital, Alkmaar, The Netherlands
[6] Department of Pulmonary Medicine, University Hospital of Cattinara, Trieste, Italy
[7] Cooperative Studies Program Coordinating Center, VA Palo Alto, Palo Alto, USA
[8] Department of Pharmaceutical Sciences, University of Tennessee Health Science Center, Memphis, USA
[9] Medecine intensive reanimation, Chru Hôpitaux De Tours, Hospital Bretonneau, Tours, France
[10] INSERM CIC1415, CHRU de Tours, Tours, France.
[11] Centre for Medicine Use and Safety, Monash University, Melbourne, Australia.
[12] Department of Medicine, The Western Health Chronic Disease Alliance and the University of Melbourne, Melbourne, Australia
[13] Pulmonary Medicine, Badalona Serveis Assistencials S.A., Badalona, Spain
[14] Pulmonary Medicine, Bellvitge University Hospital, L'Hospitalet de Llobregat, Spain
[15] Predictive Analytics and Comparative Effectiveness Center, Institute for Clinical Research and Health Policy Studies, Tufts Medical Center, Boston, USA
[16] Department of Biomedical Data Sciences, Leiden University Medical Center, Leiden, Netherlands
[17] CLEAR Methods Center, Division of Clinical Epidemiology, Department of Clinical Research, University Hospital Basel, University of Basel, Basel, Switzerland
[18] Department of Clinical Epidemiology & Biostatistics, McMaster University, Ontario, Canada
[19] Department of Medical Microbiology and Infectious Diseases and the Department of Internal Medicine, Section of Infectious Diseases , Erasmus University Medical Center, Rotterdam, The Netherlands
[20] Department of Internal Medicine, St. Antonius Hospital, Nieuwegein, Netherlands
[21] Department of Internal Medicine, Leiden University Medical Center, Leiden, Netherlands
[22] Department of Pharmacy, St. Antonius Hospital, Utrecht/Nieuwegein, Netherlands
[23] Department of Pulmonary Medicine, St. Antonius Hospital, Nieuwegein, Netherlands
[24] Hormonpraxis Aarau, Aarau, Switzerland
[25] Division of Endocrinology, Diabetes and Metabolism, Department of Clinical Research, Universitätsspital Basel, Basel, Switzerland
[26] Department of Pulmonology, Hospital Clinic. University of Barcelona, Barcelona, Spain
[27] Centro de Investigación Biomedica En Red-Enfermedades Respiratorias (CIBERES), Institut d'Inve, Universitat de Barcelona, Barcelona, Spain

### Appendix A: Literature search

Methods

Randomized controlled trials (RCTs) eligible for this study compared placebo with low-dose oral or intravenous corticosteroid therapy as adjunctive therapy in community-acquired pneumonia (CAP) patients. We excluded studies with pseudo randomization or with treatment combinations that did not allow investigation of an independent corticosteroid effect. We updated the systematic search by Briel and colleagues, which identified eligible studies up to July 2017. As such, we electronically searched Medline, Embase, and the Cochrane Controlled Trials Registry from July 2017 to November 2022 using medical subject headings based on the terms 'pneumonia' and 'corticosteroid'. Table A1 contains the detailed search strategies. Two reviewers (JS and PvdZ) independently assessed trial eligibility based on title and abstracts, full-texts, and further information from investigators if needed.

From all eligible trials, individual patient data (IPD), including demographic, clinical, and laboratory data, were requested by the authors. The data were verified against the reported results and inconsistencies were resolved with the corresponding authors. The risk of bias (ROB) arising from the randomization process, deviations from intended interventions, missing outcome data, measurement of the outcome, and selection of the reported result in included trials was assessed independently by two reviewers (JS and PvdZ), using the updated version of the Cochrane ROB assessment tool.[1]

Results

From the literature search, we identified 10 eligible trials. The trial by Dequin et al.[2] was published after the literature search, and was added post-hoc as an eligible trial (figure A1). We contacted the corresponding authors of all eligible trials. The authors of Nafae et al.[3] did not respond, the authors of Sabry et al.[4] responded that the dataset was lost, and authors of Meduri et al.[5] and Dequin et al.[2] responded, but did not share IPD yet (upon our own request to wait with this for prospective validation). Hence, IPD of six trials was included, totalling 1,869 CAP patients with randomized corticosteroid or placebo treatment. Five studies were judged as having overall low ROB, while for the remaining studies, some concerns were raised (table A2). Concerns were raised for bias arising from the randomization process, the selection of the reported result or both.

Table A1: Search query per database.

| **Database** | **Records after duplicates removed** | **Search Query** |
|---|---|---|
| Medline ALL through Ovid | 1870 | (exp Pneumonia / OR (cap OR hap OR pneumon*).ab,ti.) AND (exp Steroids / OR exp Adrenal Cortex Hormones / OR (prednison* OR prednisolon* OR methylprednisolon* OR betamethason* OR dexamethason* OR triamcinolone OR hydrocortison* OR alclometason* OR algeston* OR amcinonid* OR amelometason* OR beclometason* OR budesonid* OR butixocort* OR chloroprednison* OR ciclesonid* OR ciprocinonid* OR clobetasol* OR clobetason* OR clocortolon* OR cloprednol* OR cortivazol* OR deflazacort* OR diflorason* OR diflucortolon* OR difluprednat* OR domoprednat* OR drocinonid* OR dutimelan* OR etiprednol-dicloacetat* OR fluclorolon* OR fludrocortison* OR fludroxycortid* OR flumetason* OR flumoxonid* OR flunisolid* OR fluocinolon* OR fluocinonid* OR fluocortin* OR fluocortolon* OR fluorometholon* OR flupredniden* OR fluprednisolon* OR fluticason* OR formocortal* OR halcinonid* OR halometason* OR halopredon* OR hydrocortison* OR icometasone-enbutat* OR isoflupredon* OR itrocinonid* OR locicortolone-dicibat* OR lorinden-a* OR lorinden-t* OR loteprednol* OR mazipredon* OR medryson* OR meprednison* OR mometasone-furoat* OR nicocortonid* OR nivacortol* OR oropivalon* OR paramethason* OR prednisolon* OR prednison* OR pregnenolon* OR procinonid* OR promestrien* OR resocortol* OR rimexolon* OR rofleponid* OR ticabeson* OR timobeson* OR tipredan* OR tixocortol* OR triamcinolon* OR ulobetasol-propionat* OR uniderm* OR vamorolon* OR zoticason* OR steroid* OR corticosteroid* OR Adrenal-Cortex-Hormone* OR glucocorticoid* OR hydroxycorticosteroid*).ab,ti.) AND (Exp Controlled clinical trial/ OR "Double-Blind Method"/ OR "Single-Blind Method"/ OR "Random Allocation"/ OR (random* OR factorial* OR crossover* OR cross over* OR placebo* OR ((doubl* OR singl*) ADJ blind*) OR assign* OR allocat* OR volunteer* OR trial OR groups).ab,ti,kf.) NOT (exp Animals/ NOT Humans/) NOT ((exp child/ OR exp infant/ OR pediatrics/ OR adolescent/) NOT exp adult/) AND 2017:2030.(sa_year). |
| Embase through Embase.com | 1180 | (pneumonia/exp OR (cap OR hap OR pneumon*):Ab,ti) AND ('steroid'/de OR 'corticosteroid'/exp OR (prednison* OR prednisolon* OR methylprednisolon* OR betamethason* OR dexamethason* OR triamcinolone OR hydrocortison* OR alclometason* OR algeston* OR amcinonid* OR amelometason* OR beclometason* OR budesonid* OR butixocort* OR chloroprednison* OR ciclesonid* OR ciprocinonid* OR clobetasol* OR clobetason* OR clocortolon* OR cloprednol* OR cortivazol* OR deflazacort* OR diflorason* OR diflucortolon* OR difluprednat* OR domoprednat* OR drocinonid* OR dutimelan* OR etiprednol-dicloacetat* OR fluclorolon* OR fludrocortison* OR fludroxycortid* OR flumetason* OR flumoxonid* OR flunisolid* OR fluocinolon* OR fluocinonid* OR fluocortin* OR fluocortolon* OR fluorometholon* OR flupredniden* OR fluprednisolon* OR fluticason* OR formocortal* |

| | | |
|---|---|---|
| | | OR halcinonid* OR halometason* OR halopredon* OR hydrocortison* OR icometasone-enbutat* OR isoflupredon* OR itrocinonid* OR locicortolone-dicibat* OR lorinden-a* OR lorinden-t* OR loteprednol* OR mazipredon* OR medryson* OR meprednison* OR mometasone-furoat* OR nicocortonid* OR nivacortol* OR oropivalon* OR paramethason* OR prednisolon* OR prednison* OR pregnenolon* OR procinonid* OR promestrien* OR resocortol* OR rimexolon* OR rofleponid* OR ticabeson* OR timobeson* OR tipredan* OR tixocortol* OR triamcinolon* OR ulobetasol-propionat* OR uniderm* OR vamorolon* OR zoticason* OR steroid* OR corticosteroid* OR Adrenal-Cortex-Hormone* OR glucocorticoid* OR hydroxycorticosteroid*):ab,ti) AND ('randomised controlled trial'/exp OR 'single blind procedure'/exp OR 'double blind procedure'/exp OR 'crossover procedure'/exp OR (random* OR placebo* OR factorial* OR crossover* OR 'cross-over' OR 'cross over' OR assign* OR allocat* OR volunteer* OR ((singl* OR doubl*) NEAR/2 (blind* OR mask*))):ab,ti) NOT [conference abstract]/lim NOT ([animals]/lim NOT [humans]/lim) NOT (juvenile/exp NOT adult/exp) AND [2017-07-01]/sd |
| Cochrane Central Register of Controlled Trials through Wiley | 894 | ((cap OR hap OR pneumon*):Ab,ti) AND ((prednison* OR prednisolon* OR methylprednisolon* OR betamethason* OR dexamethason* OR triamcinolone OR hydrocortison* OR alclometason* OR algeston* OR amcinonid* OR amelometason* OR beclometason* OR budesonid* OR butixocort* OR chloroprednison* OR ciclesonid* OR ciprocinonid* OR clobetasol* OR clobetason* OR clocortolon* OR cloprednol* OR cortivazol* OR deflazacort* OR diflorason* OR diflucortolon* OR difluprednat* OR domoprednat* OR drocinonid* OR dutimelan* OR etiprednol-dicloacetat* OR fluclorolon* OR fludrocortison* OR fludroxycortid* OR flumetason* OR flumoxonid* OR flunisolid* OR fluocinolon* OR fluocinonid* OR fluocortin* OR fluocortolon* OR fluorometholon* OR flupredniden* OR fluprednisolon* OR fluticason* OR formocortal* OR halcinonid* OR halometason* OR halopredon* OR hydrocortison* OR icometasone-enbutat* OR isoflupredon* OR itrocinonid* OR locicortolone-dicibat* OR lorinden-a* OR lorinden-t* OR loteprednol* OR mazipredon* OR medryson* OR meprednison* OR mometasone-furoat* OR nicocortonid* OR nivacortol* OR oropivalon* OR paramethason* OR prednisolon* OR prednison* OR pregnenolon* OR procinonid* OR promestrien* OR resocortol* OR rimexolon* OR rofleponid* OR ticabeson* OR timobeson* OR tipredan* OR tixocortol* OR triamcinolon* OR ulobetasol-propionat* OR uniderm* OR vamorolon* OR zoticason* OR steroid* OR corticosteroid* OR Adrenal-Cortex-Hormone* OR glucocorticoid* OR hydroxycorticosteroid*):ab,ti) |

Table A2: Results of risk of bias assessment for each eligible study. The updated version of the Cochrane ROB assessment tool[1] was used to assess bias arising from the randomization process (R), deviations from intended interventions (D), missing outcome data (Mi), measurement of the outcome (Me), and selection of the reported result (S). Overall ROB was judged to be low if ROB was judged to be low in all domains. Overall ROB (O) was judged as 'some concerns' if some concerns were raised in at least one domain, but no domain was judged as high ROB.

| ***First author, year (reference)*** | **R** | **D** | **Mi** | **Me** | **S** | **O** |
|---|---|---|---|---|---|---|
| *Confalonieri, 2005* | SOME CONCERNS | LOW RISK | LOW RISK | LOW RISK | SOME CONCERNS | SOME CONCERNS |
| *Snijders, 2010* | LOW RISK | LOW RISK | LOW RISK | LOW RISK | SOME CONCERNS | SOME CONCERNS |
| *Meijvis, 2011* | LOW RISK | LOW RISK | LOW RISK | LOW RISK | LOW RISK | LOW RISK |
| *Sabry, 2011* | SOME CONCERNS | LOW RISK | LOW RISK | LOW RISK | SOME CONCERNS | SOME CONCERNS |
| *Nafae, 2013* | SOME CONCERNS | LOW RISK | LOW RISK | LOW RISK | SOME CONCERNS | SOME CONCERNS |
| *Torres, 2015* | LOW RISK | LOW RISK | LOW RISK | LOW RISK | LOW RISK | LOW RISK |
| *Blum, 2015* | LOW RISK | LOW RISK | LOW RISK | LOW RISK | LOW RISK | LOW RISK |
| *Wittermans, 2021* | LOW RISK | LOW RISK | LOW RISK | LOW RISK | LOW RISK | LOW RISK |
| *Meduri, 2022* | LOW RISK | LOW RISK | LOW RISK | LOW RISK | SOME CONCERNS | SOME CONCERNS |
| *Dequin, 2023* | LOW RISK | LOW RISK | LOW RISK | LOW RISK | LOW RISK | LOW RISK |

Figure A1: Flow diagram of study selection.

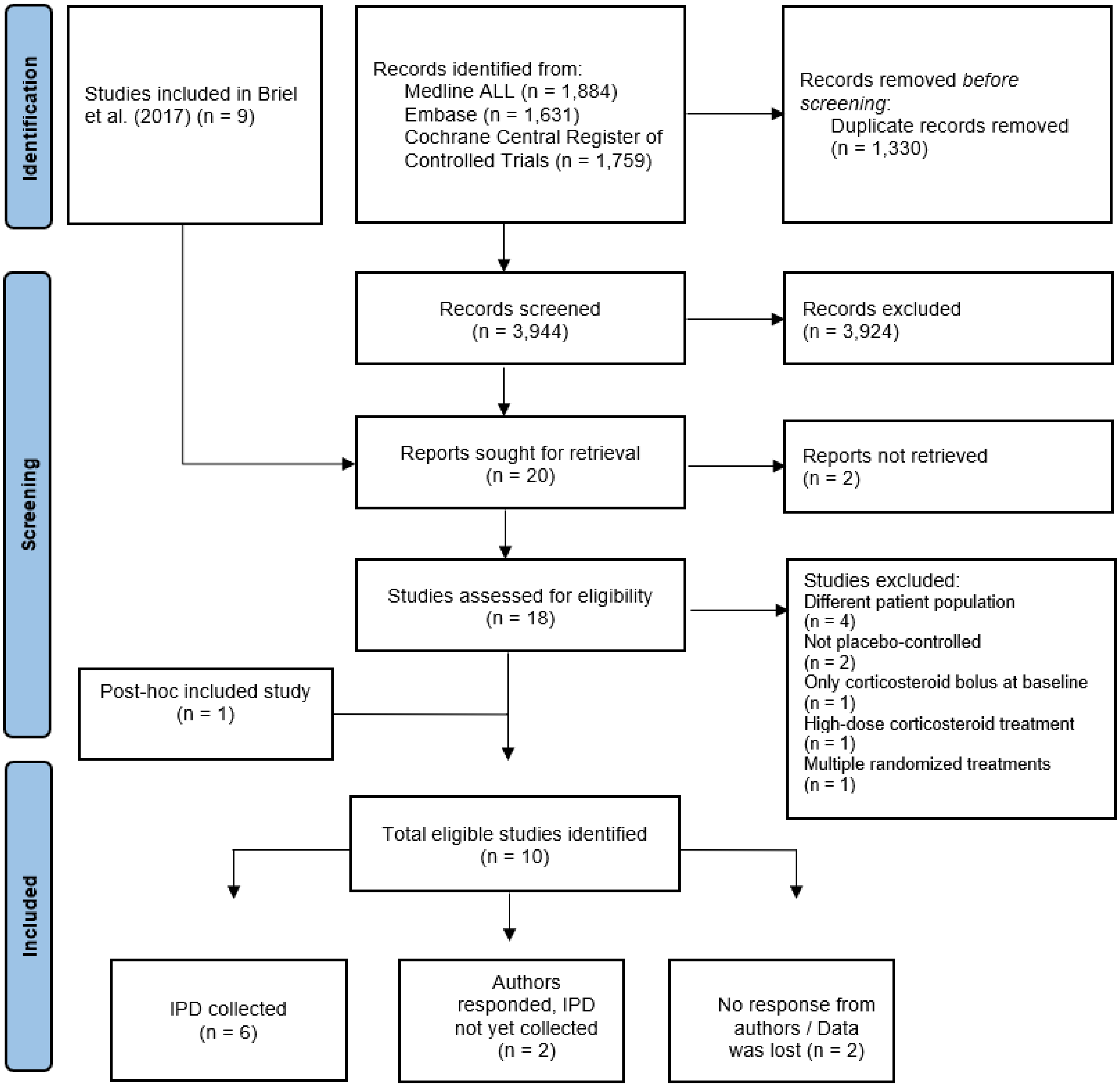

**Appendix B: Definition of the 'Area under the benefit curve' (AUC-benefit)**

The AUC-benefit involves considering different ITE thresholds to divide patients into two groups: a predicted harm group (where ITE ≤ threshold) and a predicted benefit group (where ITE > threshold). Both groups are further divided into those who received corticosteroids ($G_1$ and $G_3$) and those who received placebo ($G_2$ and $G_4$, Figure B1).

The Δ-benefit is defined as follows:

$$\Delta_{benefit} = \left[\frac{\sum_{i \subseteq G_2} y_i}{n_2} - \frac{\sum_{i \subseteq G_1} y_i}{n_1}\right] - \left[\frac{\sum_{i \subseteq G_4} y_i}{n_4} - \frac{\sum_{i \subseteq G_3} y_i}{n_3}\right]$$

where i indexes the patient, $y_i$ equals 1 in case 30-day mortality and 0 otherwise, and $n_{1\text{-}4}$ denote the number of patients in $G_{1\text{-}4}$.

The Δ-benefit is calculated considering a range of ten thresholds, starting with a threshold at the 25$^{th}$ percentile, and increase the percentiles in ten equal steps until the 75$^{th}$ percentile of the full ITE distribution. The calculated Δ-benefits for the different thresholds forms the 'Δ-benefit-curve', and the area under the Δ-benefit-curve (AUC-benefit) is calculated as the trapezoidal area under this curve (Figure B2). We used Sklearn's 'metrics.auc' function to calculate the AUC-benefit.

Figure B1: Schematic overview of patient grouping according to a certain ITE threshold.

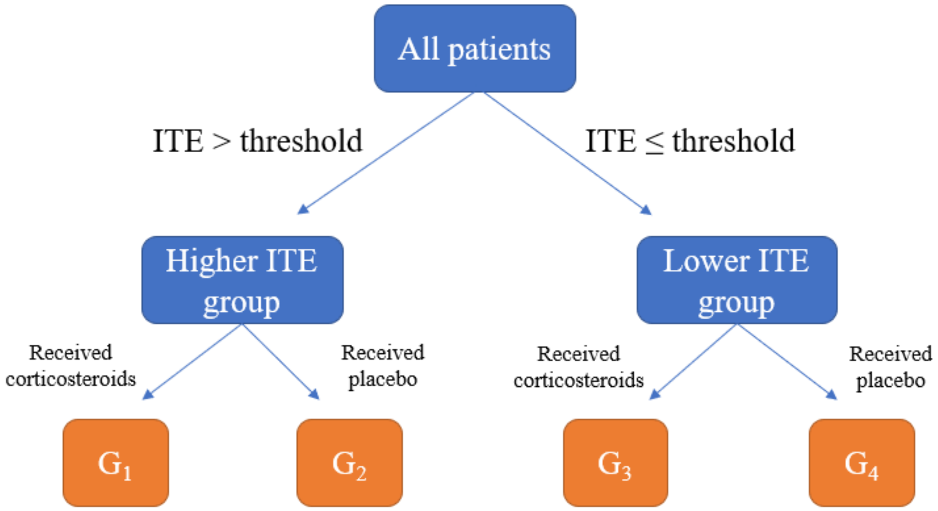

Figure B2: Schematic overview of the area under the benefit-curve (AUC-benefit).

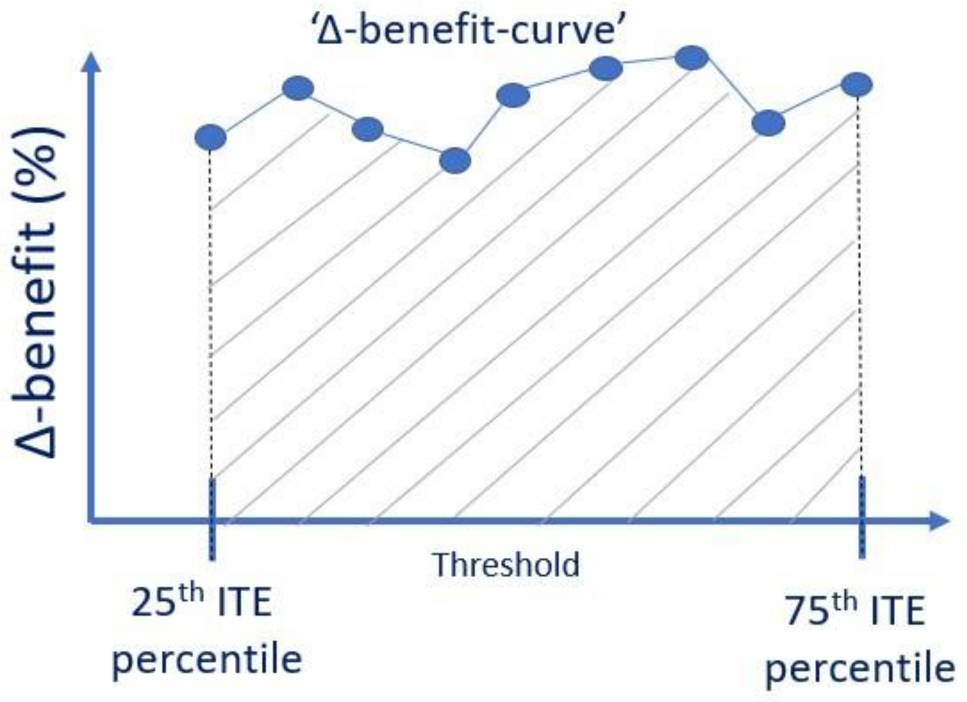

### Appendix C: Implementation of the LASSO penalty

We used Statsmodels' 'fit_regularized' function to implement penalized logistic regression,[6] which minimizes the following loss function:

$$\frac{LL}{2 * n_{train}} + \lambda\Big((1 - L1_{wt})\frac{||x||_2^2}{2} + L1_{wt}||x||_1\Big)$$

where LL represents the logistic loss, $n_{train}$ the number of obervations (ie, patients) used to train the model, λ the penalization strength, $L1_{wt}$ the fraction of the penalty given to the L1 penalty term (ie, $L1_{wt}$=0 results in a Ridge fit, $L1_{wt}$=1 results in a LASSO fit), and $||x||_2$ and $||x||_1$ the L2 and L1 norms, respectively.

In this particular implementation, the penalization strength is influenced by the training data's size ($n_{train}$). In our study, we employed a leave-one-trial-out cross-validation, which resulted in variations in the size of the training set across different folds. For instance, the training set was notably smaller in the cross-validation fold where the study conducted by Blum et al.,[7] which accounted for 42% of all patients included in our study, formed the test cohort. To enhance the robustness of the penalization approach against changes in training set size across the cross-validation folds, we modified the loss function as follows:

$$\frac{LL}{2 * n_{train}} + \frac{\lambda}{n_{train}}\Big((1 - L1_{wt})\frac{||x||_2^2}{2} + L1_{wt}||x||_1\Big)$$

We implemented this by dividing λ of the 'fit_regularized' function (in the Statsmodels' implementation this argument is called 'alpha') by the size of the train set.

### Appendix D: Alternative modelling procedures

Methods

We evaluated the performance of alternative modelling procedures varying choices regarding:

- the **modelling strategy** for heterogeneity of treatment effect (ie, risk vs effect modelling[8])
- for the effect modelling strategy:
  - the inclusion of **main effects** in the logistic regression model
  - the inclusion of an **intercept term** in the logistic regression model
  - the **encoding** of the treatment variable (ie, {0, 1} vs {-1, 1}

yielding a total of nine different models (table D1). For all the models in table D1, except the risk modelling procedure, we performed:

- unpenalized logistic regression
- penalized logistic regression with Lasso penalty
- penalized logistic regression with Ridge penalty.

For the risk modelling procedure, we only performed unpenalized logistic regression.

This yielded a total of 25 unique modelling procedures. We evaluated each procedure, without and with additional dichotomized variables, in the same leave-one-trial-out cross-validation (LOTO-CV) using the six trials (as described in the main text). We compared the performance of the different procedures in terms of discrimination for benefit (using AUC-benefit) and calibration for benefit.

For all the models summed up in table D1, referred to as 'risk' and 'effect 1-8', the differences compared to the modelling procedure described in the main text (ie, 'procedure 1 and 2) are summarized below. Please note that 'effect-8' with LASSO penalization is exactly procedure 1 (when modelled without additional dichotomized variables) and procedure 2 (when modelled with additional dichotomized variables) described in the main text.

*Risk*

This procedure is identical to the one described in the main text, until step 2. In step 3, the treatment variable is encoded as 0=placebo, 1=corticosteroids. Then, no interaction terms are introduced in the logistic regression model, but only the included variables as main effects, the treatment variable, and an intercept term. This model is trained without penalization. In risk modelling, because only the treatment variable (and no interaction terms) is added to the model, a constant relative treatment effect is assumed. Hence, all predicted ITEs will be either positive or negative (depending on the direction of the overall treatment effect).

*Effect-1*

This procedure is identical to the one described in the main text, until step 2. In step 3, the treatment variable is encoded as 0=placebo, 1=corticosteroids. Then, the interaction terms are created, and are added to the model together with the treatment variable, the selected variables as main effects and an intercept term. The optimization of λ (for the penalized models) is again identical to the procedure described in the main text.

*Effect-2*

This procedure is identical to the one described in the main text, until step 3. Then, the interaction terms are created, and are added to the model together with the treatment variable, the selected variables as main effects and an intercept term. The optimization of λ (for the penalized models) is again identical to the procedure described in the main text.

*Effect-3*

This procedure is identical to the one described in the main text, until step 2. In step 3, the treatment variable is encoded as 0=placebo, 1=corticosteroids. Then, the interaction terms are created, and are added to the model together with the treatment variable and the selected variables as main effects. The optimization of λ (for the penalized models) is again identical to the procedure described in the main text.

*Effect-4*

This procedure is identical to the one described in the main text, until step 3. Then, the interaction terms are created, and are added to the model together with the treatment variable and the selected variables as main effects. The optimization of λ (for the penalized models) is again identical to the procedure described in the main text.

*Effect-5*

This procedure is identical to the one described in the main text, until step 2. In step 3, the treatment variable is encoded as 0=placebo, 1=corticosteroids. Then, the interaction terms are created, and are added to the model together with the treatment variable and an intercept term. The optimization of λ (for the penalized models) is again identical to the procedure described in the main text.

*Effect-6*

This procedure is identical to the one described in the main text, until step 3. Then, the interaction terms are created, and are added to the model together with the treatment variable and an intercept term. The optimization of λ (for the penalized models) is again identical to the procedure described in the main text.

*Effect-7*

This procedure is identical to the one described in the main text, until step 2. In step 3, the treatment variable is encoded as 0=placebo, 1=corticosteroids. Then, the interaction terms are created, and are added to the model together with the treatment variable. The optimization of λ (for the penalized models) is again identical to the procedure described in the main text.

Table D1: Description of the different modelling procedures, where i indexes the patients, Y is the mortality, T is the treatment (ie, corticosteroids or placebo), X is the set of included variables, $\beta_0$ is the intercept term, $\beta_t$ is the coefficient for the treatment variable, $\beta_m$ includes the coefficients for the main effects ($x_i$) and $\beta_z$ includes the coefficients for the treatment-variable interaction terms ($x_i$ $t_i$) for an individual patient.

| **Modelling strategy** | **Model name** | **Main effects** | **Intercept term** | **Encoding treatment variable** | **Formula** |
|---|---|---|---|---|---|
| **Risk modelling** | Risk | ✓ | ✓ | {0, 1} | $Logit\,[P(Y_i = 1 \vert T = t_i, X = x_i)] = \beta_0 + \beta_t \underbrace{t_i}_{0,1} + \beta_m x_i$ |
| **Effect modelling** | Effect-1 | ✓ | ✓ | {0, 1} | $Logit\,[P(Y_i = 1 \vert T = t_i, X = x_i)] = \beta_0 + \beta_t \underbrace{t_i}_{0,1} + \beta_m x_i + \beta_z x_i \underbrace{t_i}_{0,1}$ |
| | Effect-2 | ✓ | ✓ | {-1, 1} | $Logit\,[P(Y_i = 1 \vert T = t_i, X = x_i)] = \beta_0 + \beta_t \underbrace{t_i}_{-1,1} + \beta_m x_i + \beta_z x_i \underbrace{t_i}_{-1,1}$ |
| | Effect-3 | ✓ | ✗ | {0, 1} | $Logit\,[P(Y_i = 1 \vert T = t_i, X = x_i)] = \beta_t \underbrace{t_i}_{0,1} + \beta_m x_i + \beta_z x_i \underbrace{t_i}_{0,1}$ |
| | Effect-4 | ✓ | ✗ | {-1, 1} | $Logit\,[P(Y_i = 1 \vert T = t_i, X = x_i)] = \beta_t \underbrace{t_i}_{-1,1} + \beta_m x_i + \beta_z x_i \underbrace{t_i}_{-1,1}$ |
| | Effect-5 | ✗ | ✓ | {0, 1} | $Logit\,[P(Y_i = 1 \vert T = t_i, X = x_i)] = \beta_0 + \beta_t \underbrace{t_i}_{0,1} + \beta_z x_i \underbrace{t_i}_{0,1}$ |
| | Effect-6 | ✗ | ✓ | {-1, 1} | $Logit\,[P(Y_i = 1 \vert T = t_i, X = x_i)] = \beta_0 + \beta_t \underbrace{t_i}_{-1,1} + \beta_z x_i \underbrace{t_i}_{-1,1}$ |
| | Effect-7 | ✗ | ✗ | {0, 1} | $Logit\,[P(Y_i = 1 \vert T = t_i, X = x_i)] = \beta_t \underbrace{t_i}_{0,1} + \beta_z x_i \underbrace{t_i}_{0,1}$ |
| | Effect-8 (main text) | ✗ | ✗ | {-1, 1} | $Logit\,[P(Y_i = 1 \vert T = t_i, X = x_i)] = \beta_t \underbrace{t_i}_{-1,1} + \beta_z x_i \underbrace{t_i}_{-1,1}$ |

Results

The results of the grid searches for all modelling procedures without, and with additional dichotomized variables that used penalization, are visualized in Figures D1 and D2, respectively. The resulting weights of the trained model of the different modelling procedures without, and with additional dichotomized variables, are visualized in Figure D3 and D4, respectively. Both with and without additional dichotomized variables, the effect-8 model using a Lasso penalty resulted in the highest discriminative performance and are therefore selected as the modelling procedures presented in the main text (Figures D5 and D6).

Figure D1: Results of the initial (wide) and (secondary) fine grid searches for λ optimalization in each LOTO-CV fold, resulting from the different variations of the modelling procedures without additional dichotomized variables.

(a) Effect-1, Lasso

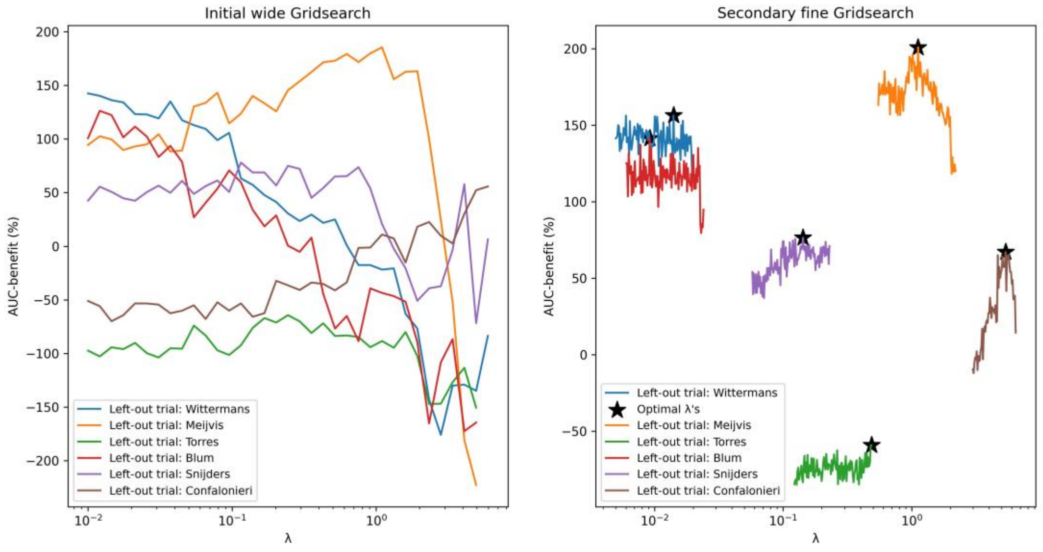

(b) Effect-1, Ridge

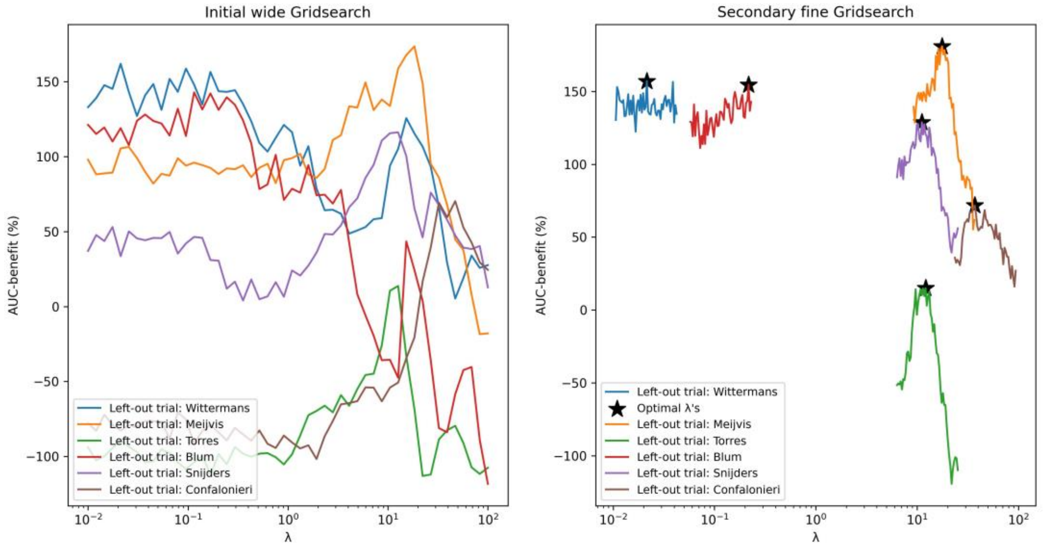

(c) Effect-2, Lasso

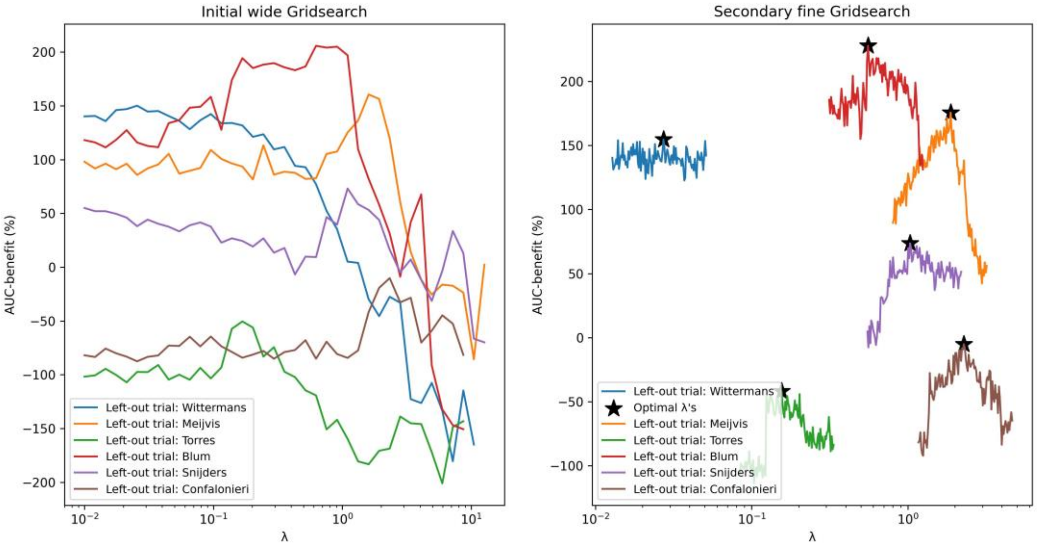

(d) Effect-2, Ridge

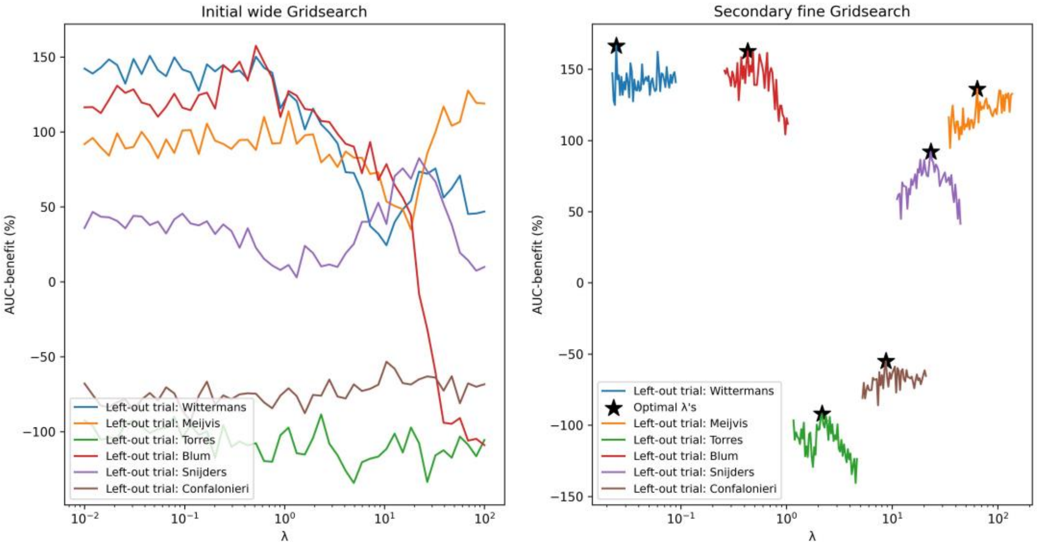

(e) Effect-3, Lasso

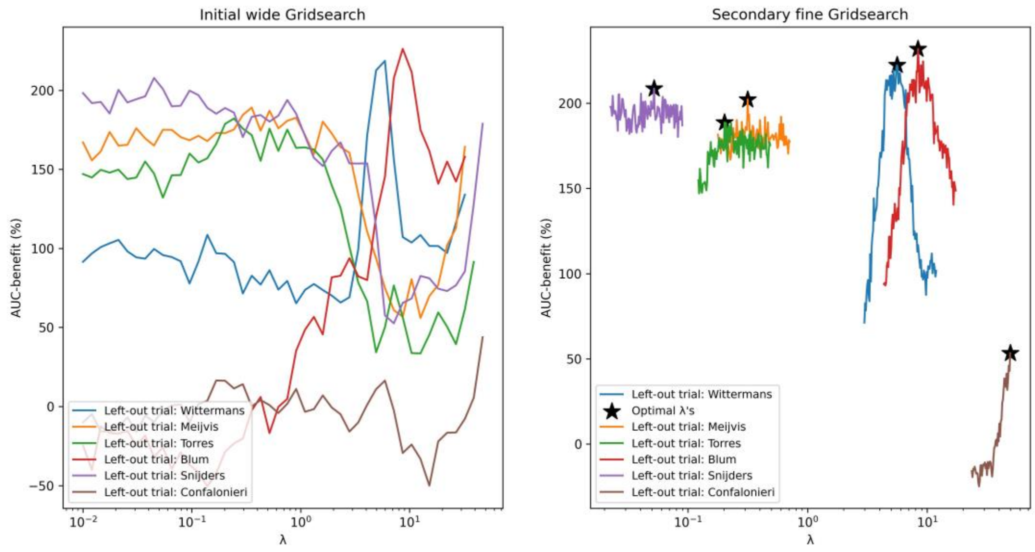

(f) Effect-3, Ridge

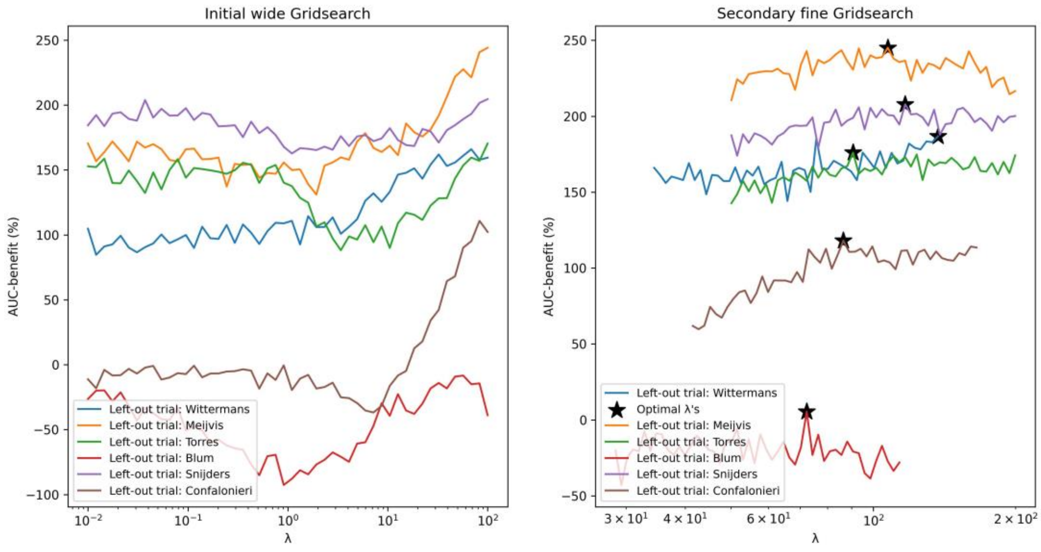

(g) Effect-4, Lasso

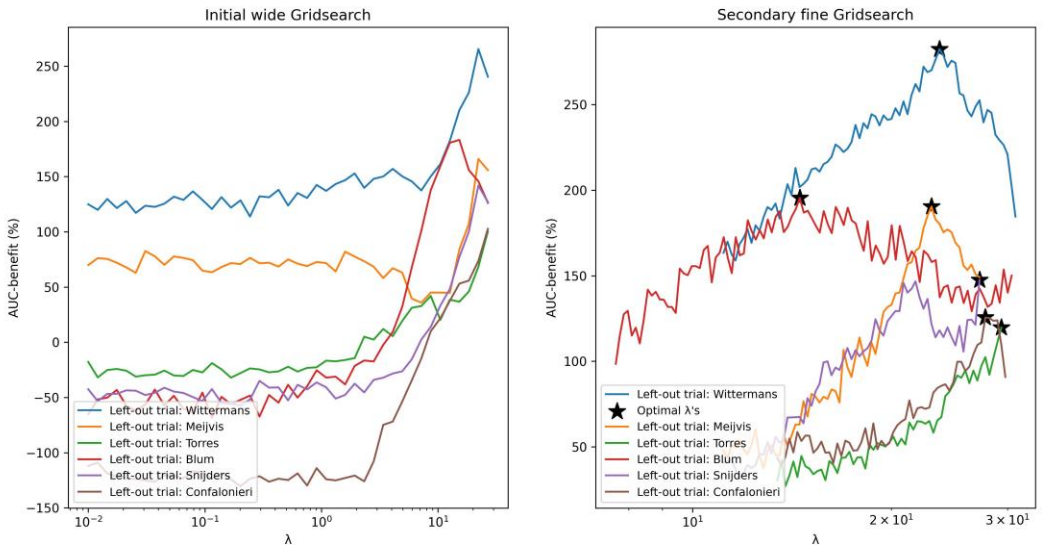

(h) Effect-4, Ridge

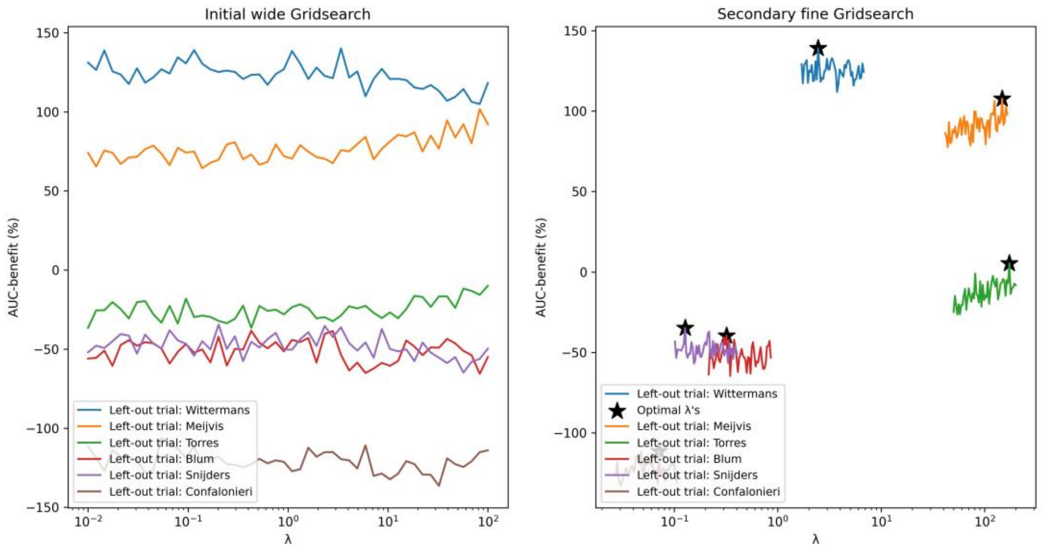

(i) Effect-5, Lasso

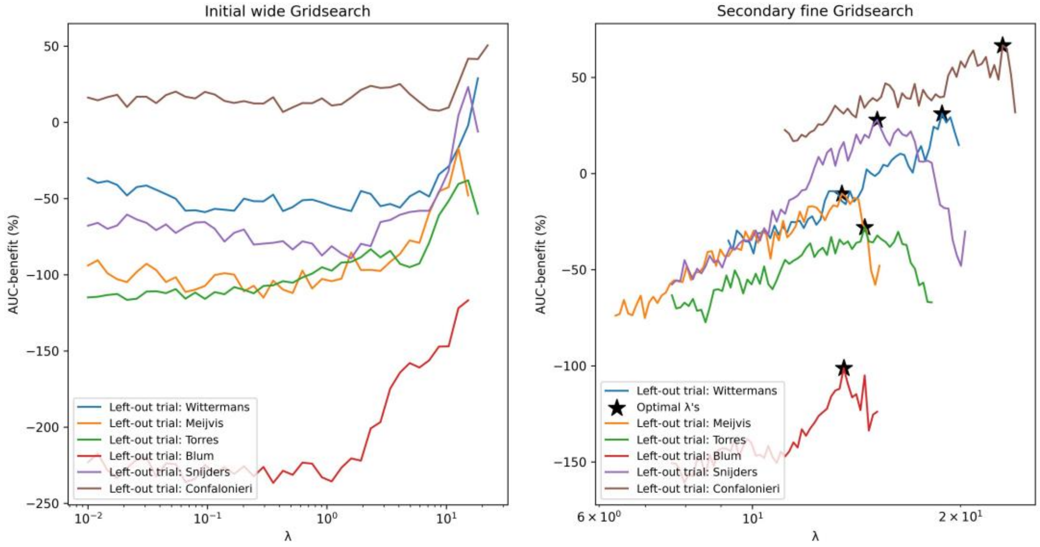

(j) Effect-5, Ridge

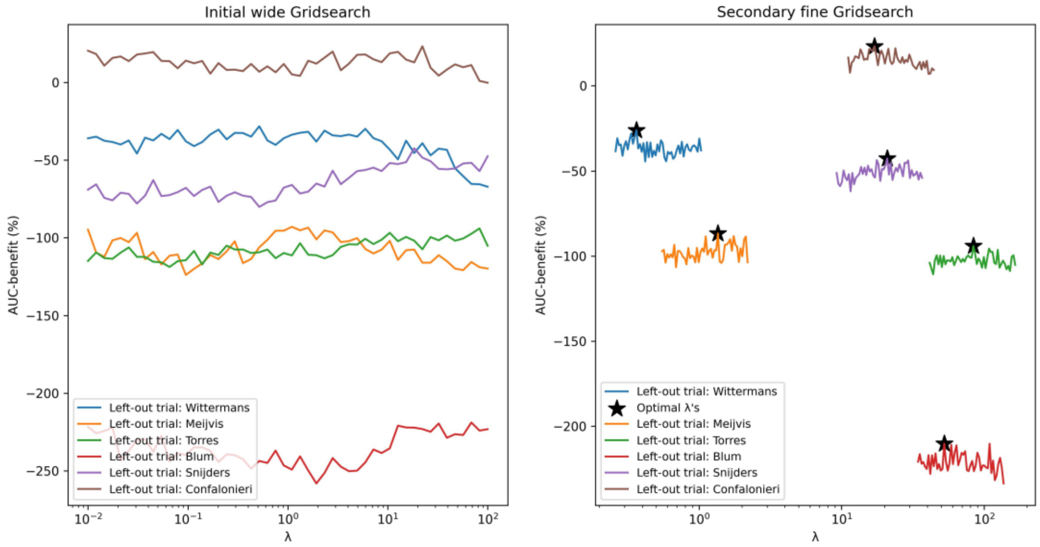

(k) Effect-6, Lasso

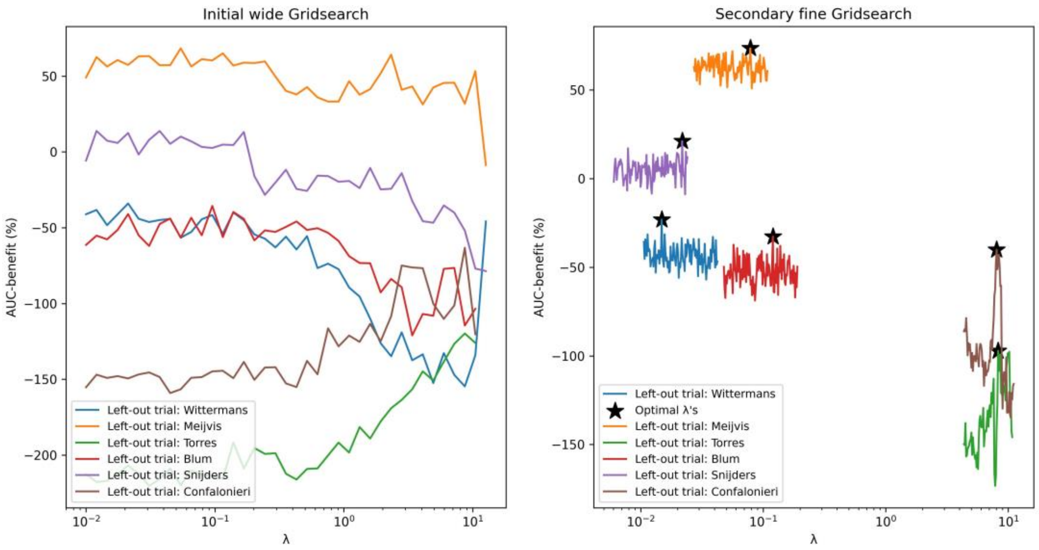

(l) Effect-6, Ridge

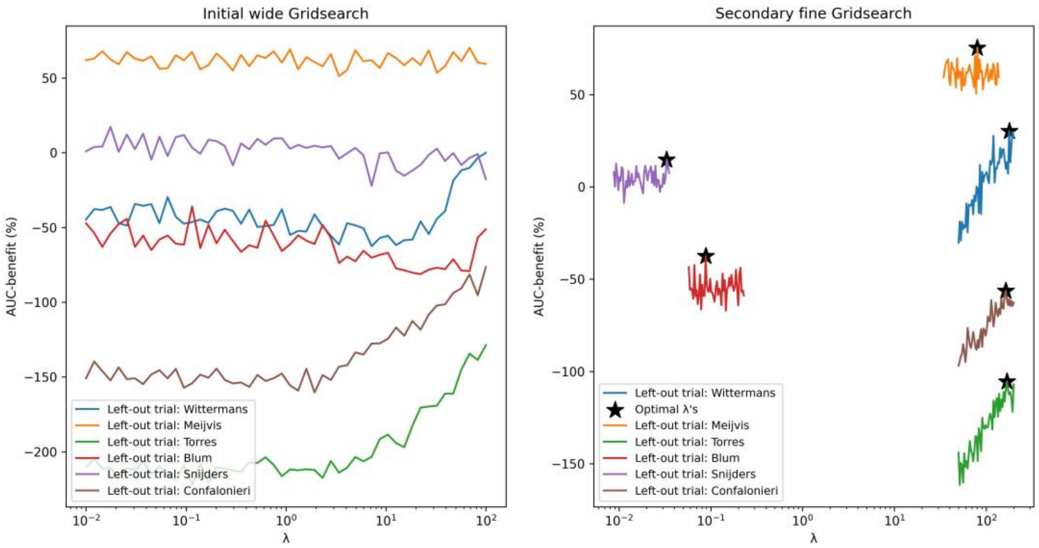

(m) Effect-7, Lasso

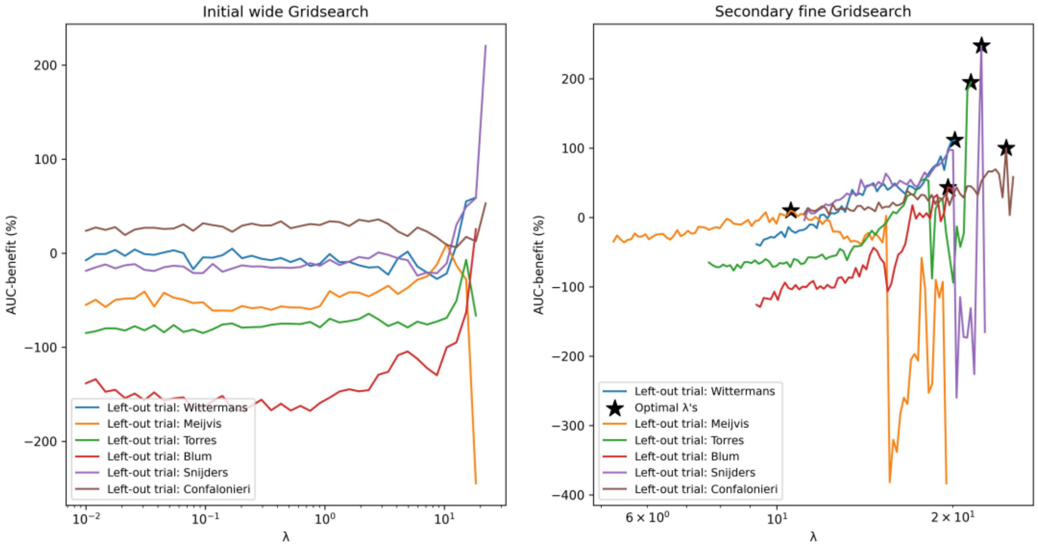

(n) Effect-7, Ridge

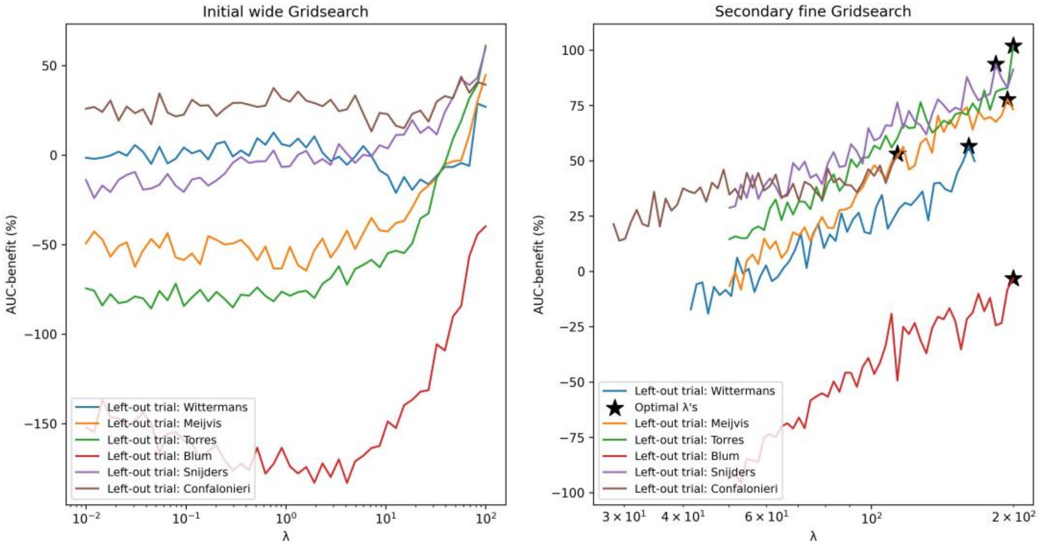

(o) Effect-8, Lasso ( ‘procedure 1’ in the main text)

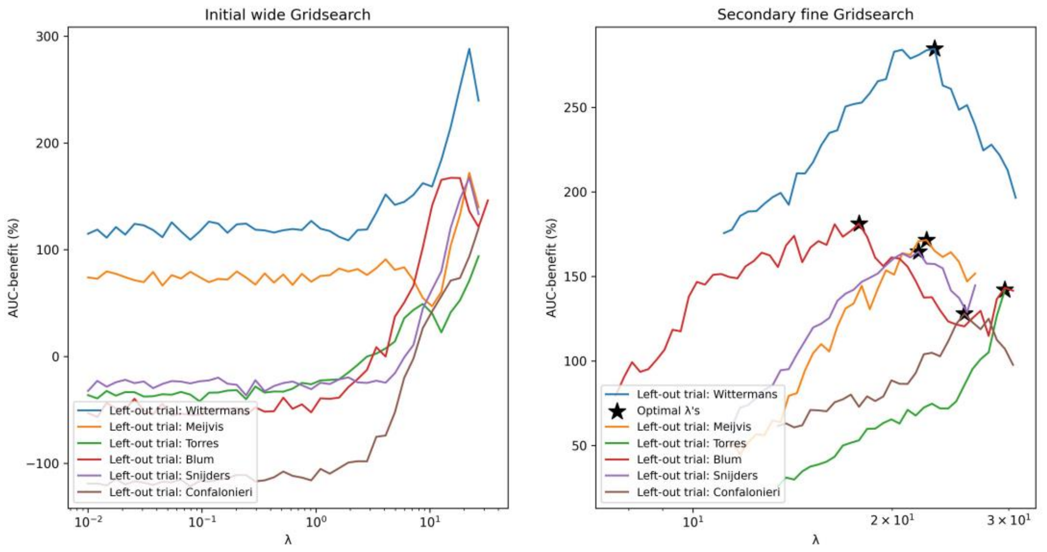

(p) Effect-8, Ridge

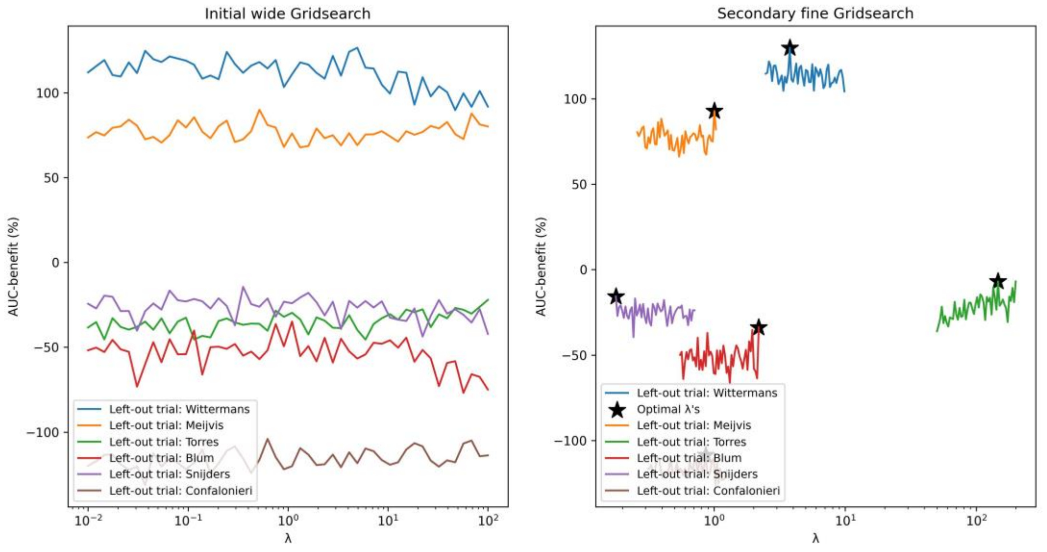

Figure D2: Results of the initial (wide) and (secondary) fine grid searches for λ optimalization in each LOTO-CV fold, resulting from the different variations of modelling procedures with additional dichotomized variables.

(a) Effect-1, Lasso

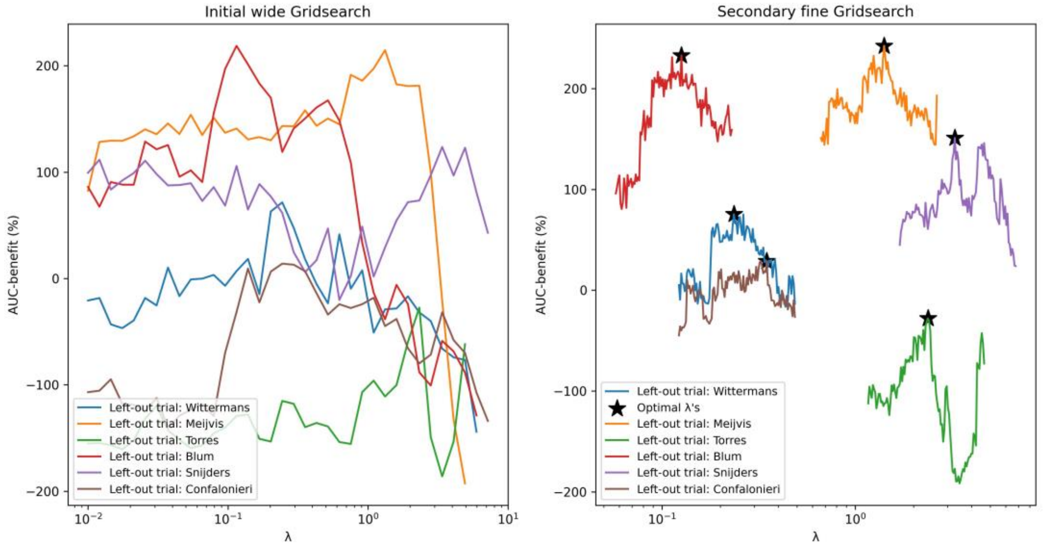

(b) Effect-1, Ridge

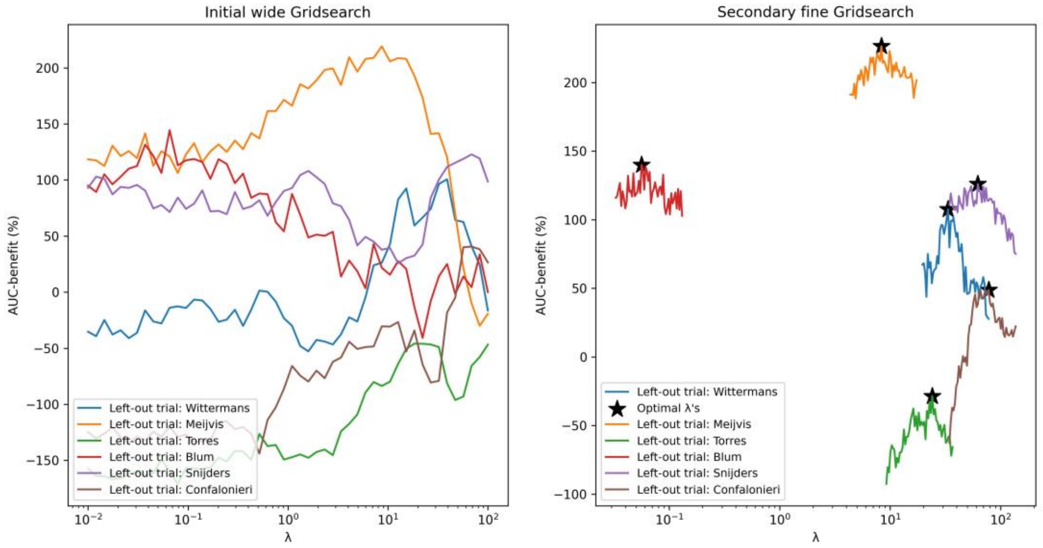

(c) Effect-2, Lasso

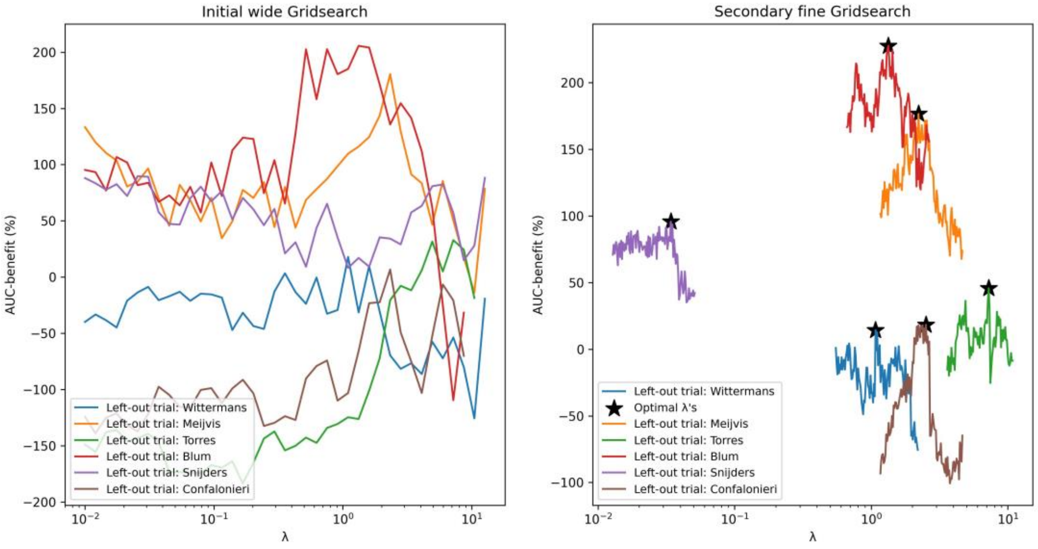

(d) Effect-2, Ridge

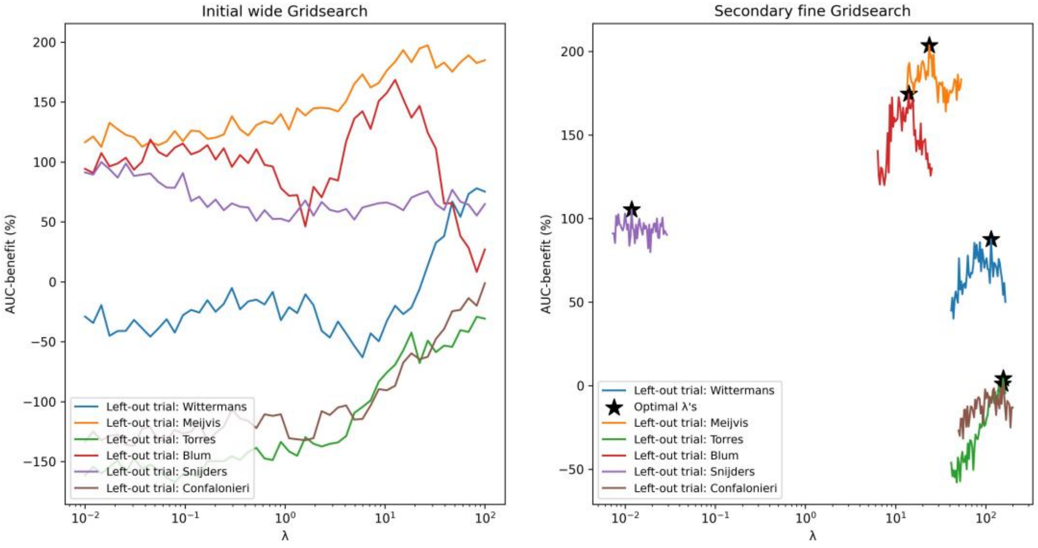

(e) Effect-3, Lasso

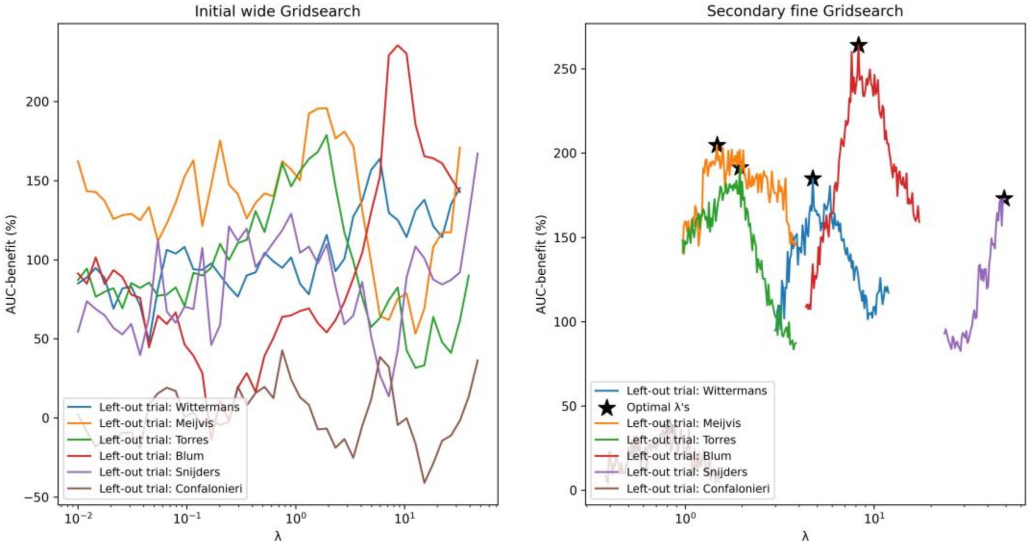

(f) Effect-3, Ridge

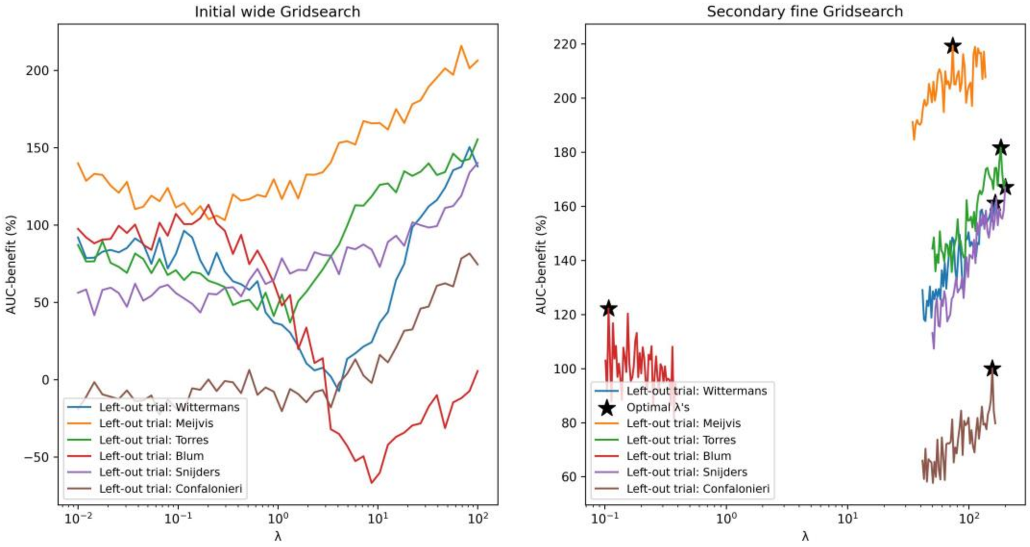

(g) Effect-4, Lasso

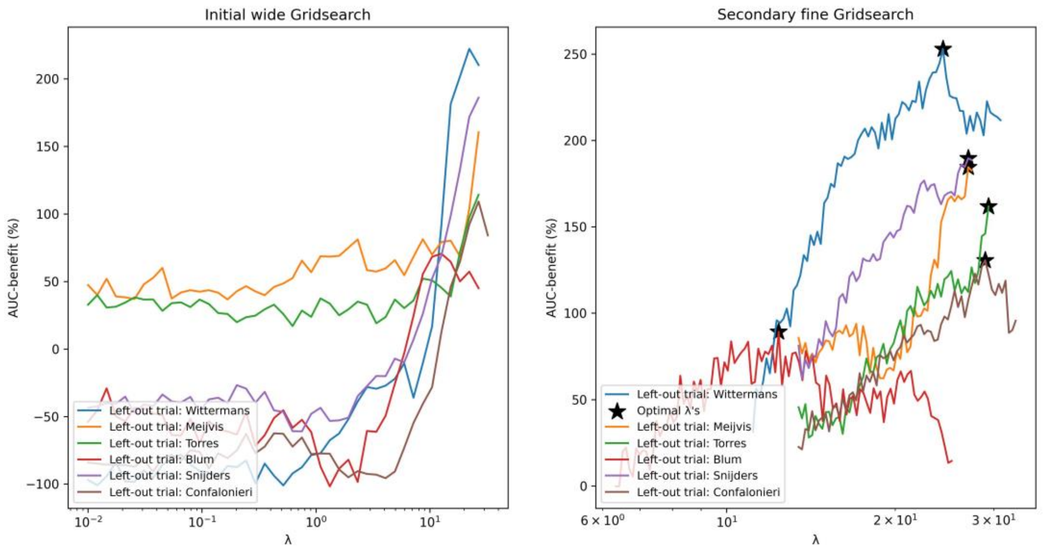

(h) Effect-4, Ridge

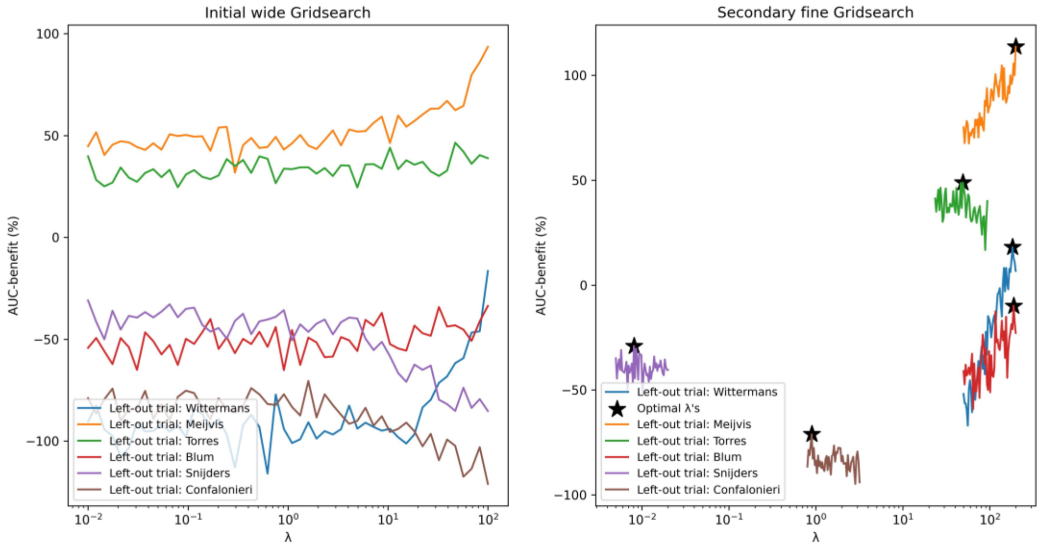

(i) Effect-5, Lasso

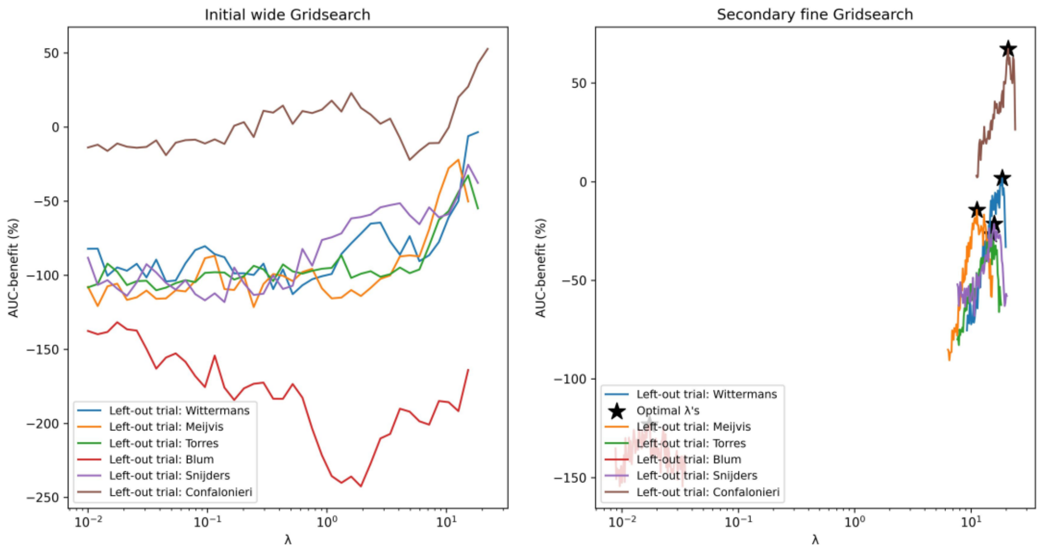

(j) Effect-5, Ridge

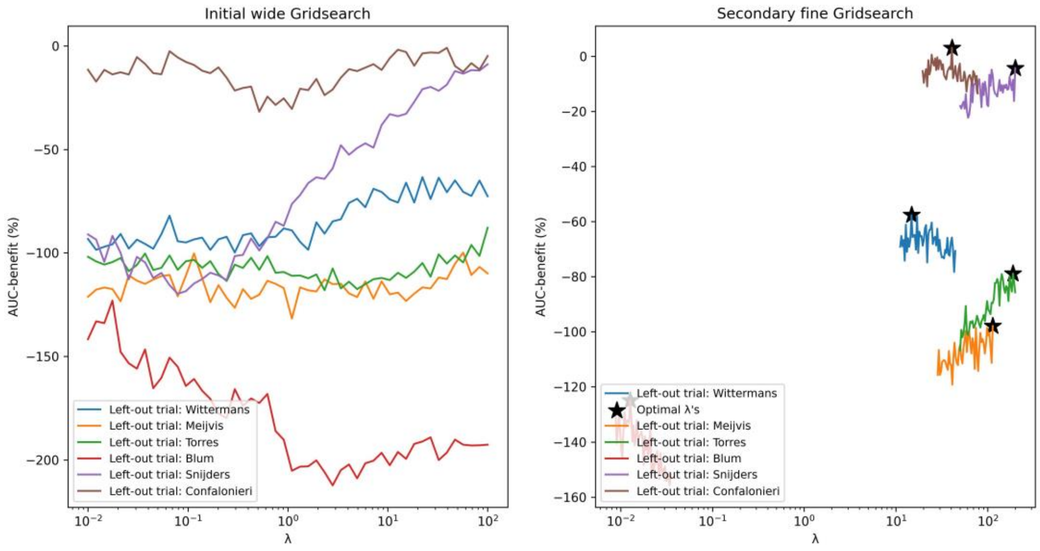

(k) Effect-6, Lasso

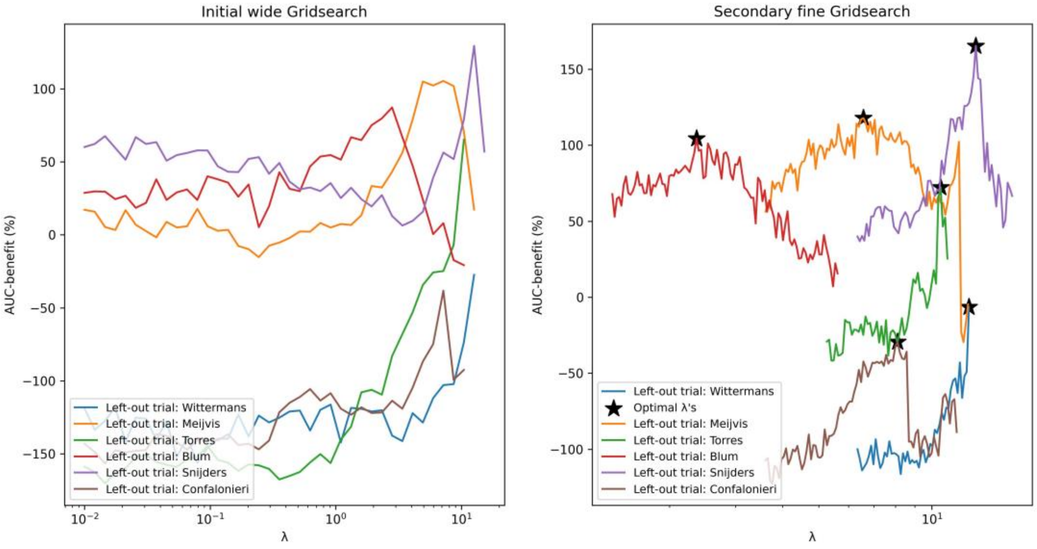

(l) Effect-6, Ridge

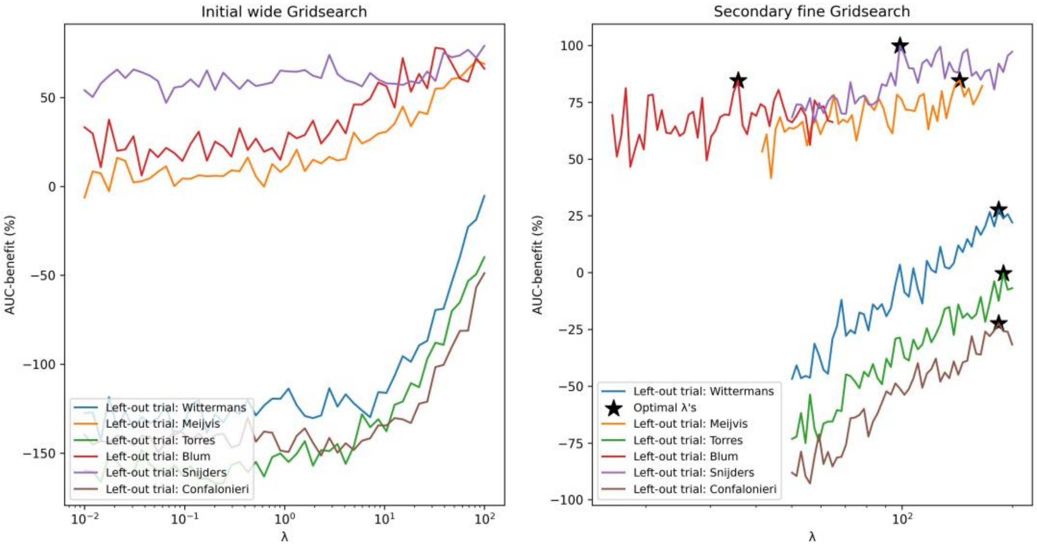

(m) Effect-7, Lasso

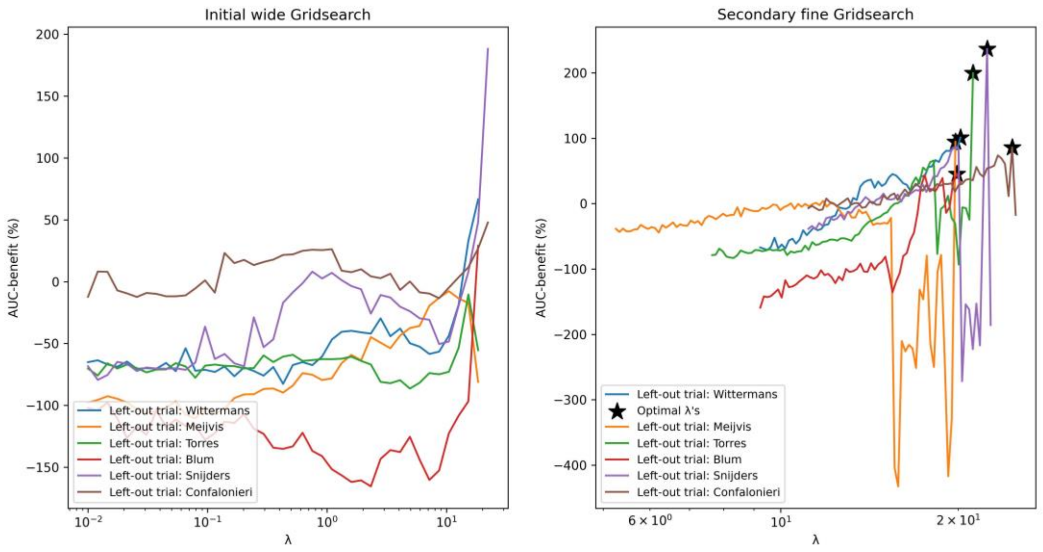

(n) Effect-7, Ridge

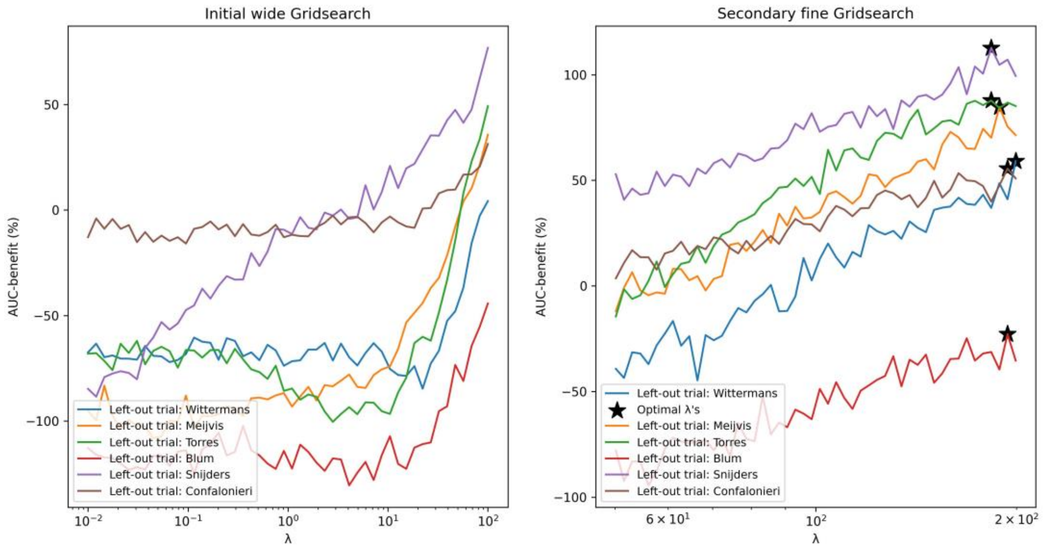

(o) Effect-8, Lasso (‘procedure 2’ in the main text)

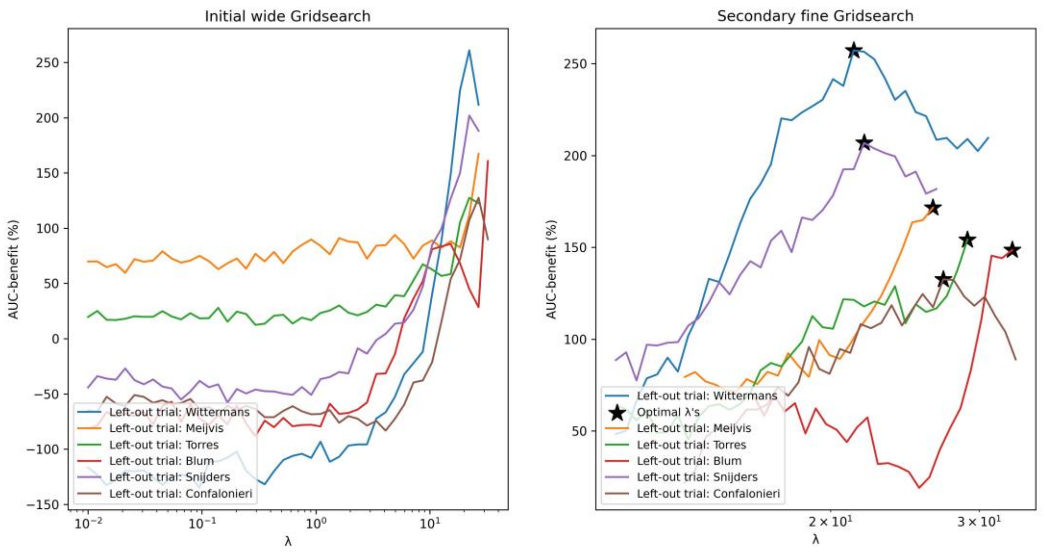

(p) Effect-8, Ridge

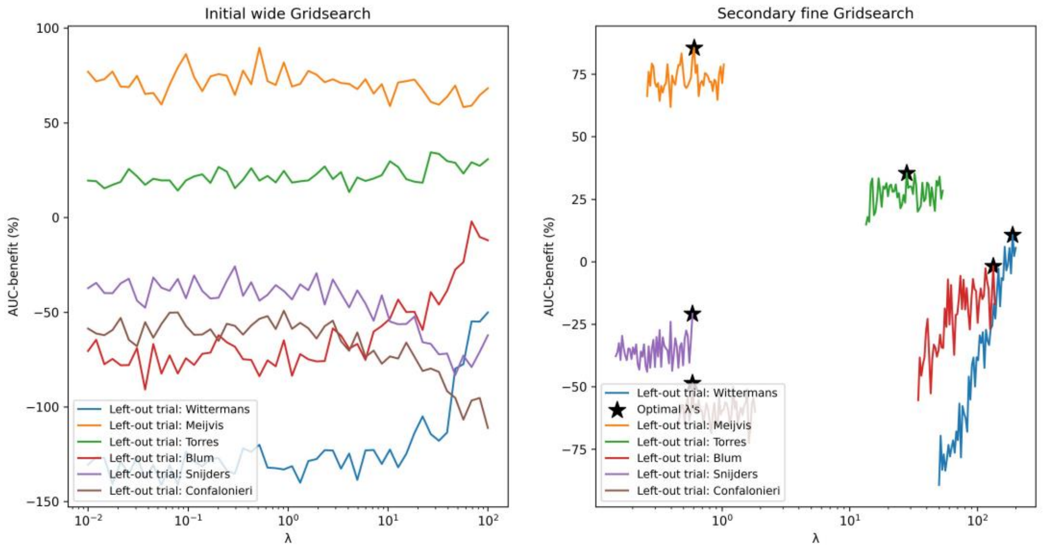

Figure D3: Bar charts of all non-zero weights of the fitted logistic regression models in each LOTO-CV fold, resulting from the different variations of modelling procedures without additional dichotomized variables. RR=respiratory rate, DBP=Diastolic blood pressure, SBP=Systolic blood pressure, Temp.=Body temperature, CRP=C-reactive protein, WBC=White cell count, T=Treatment variable

(a) Risk

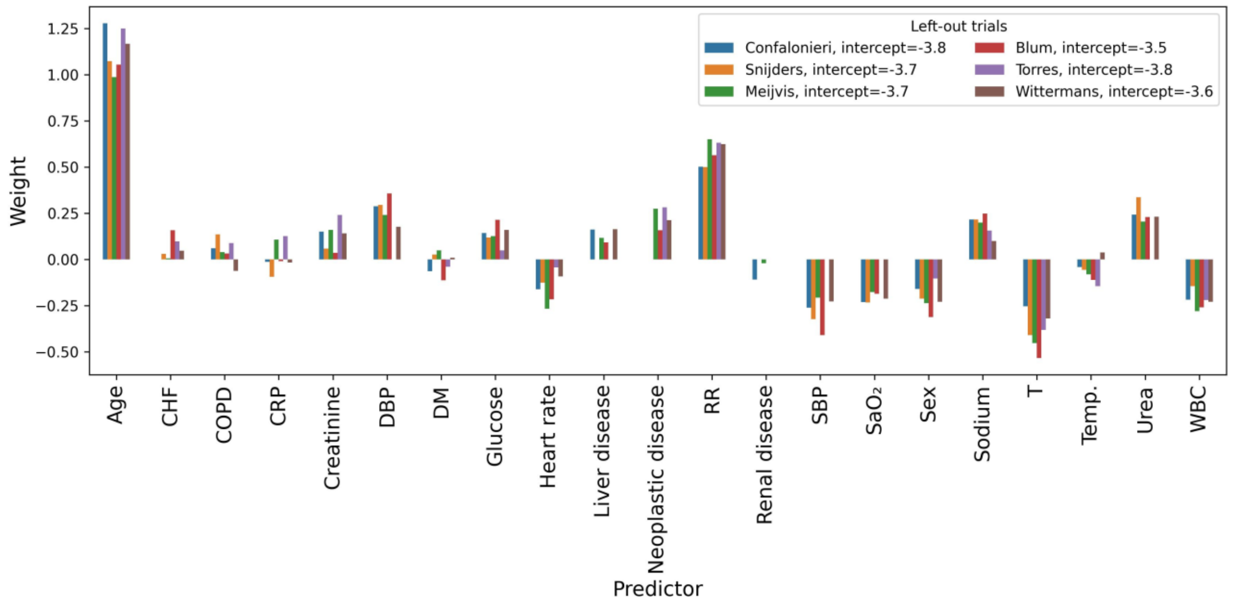

(b) Effect-1, unpenalized

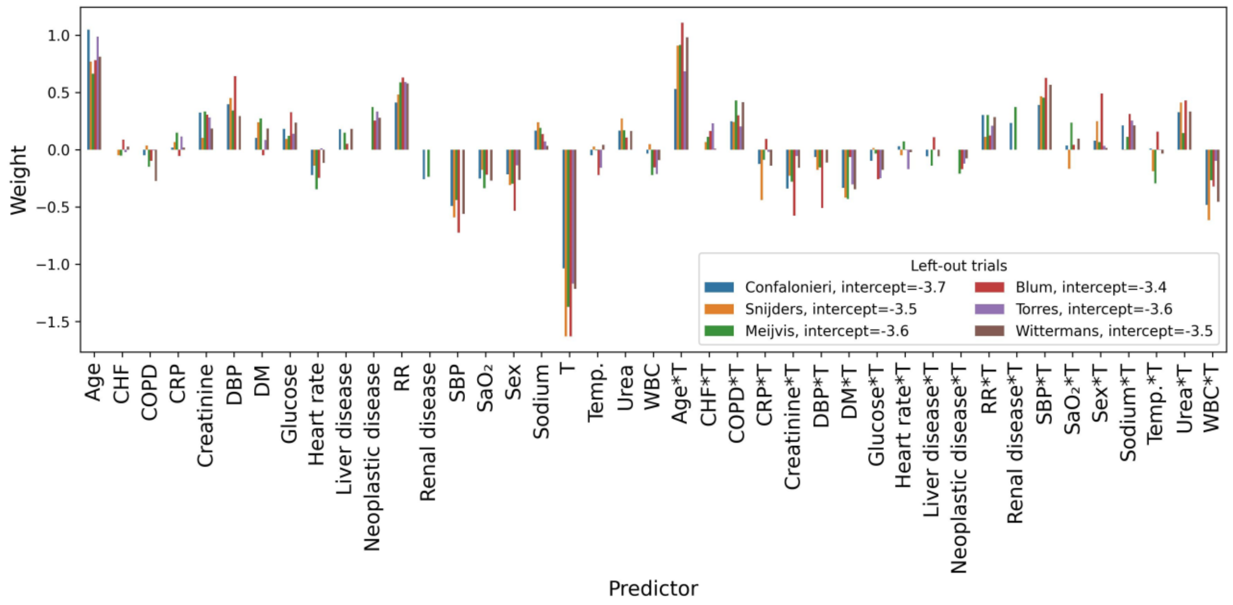

(c) Effect-1, Lasso

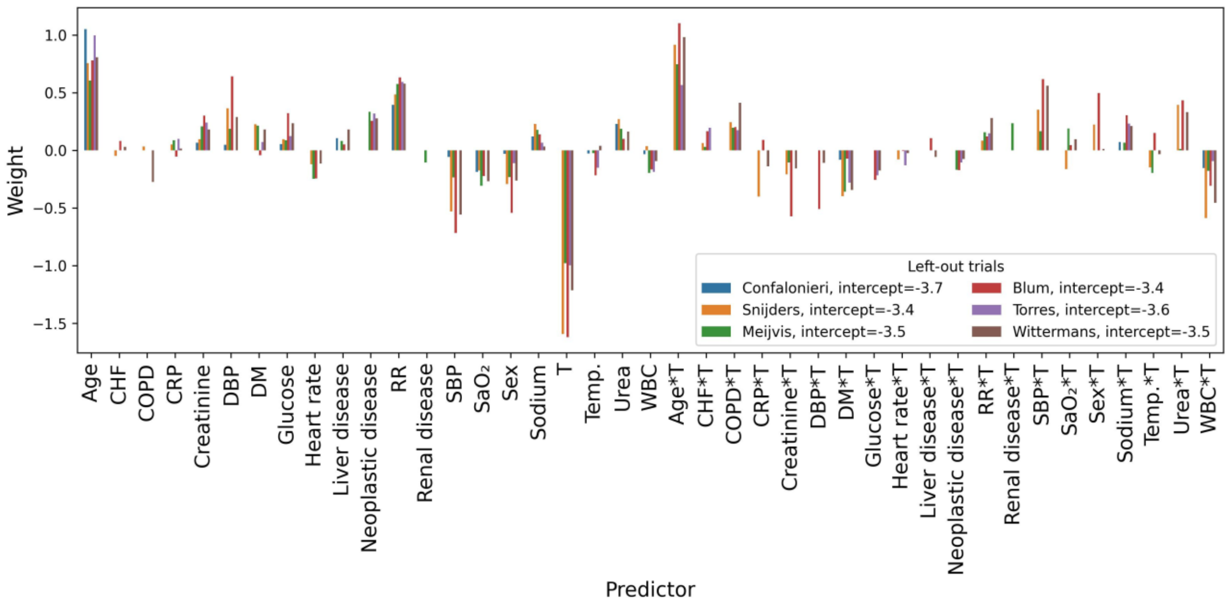

(d) Effect-1, Ridge

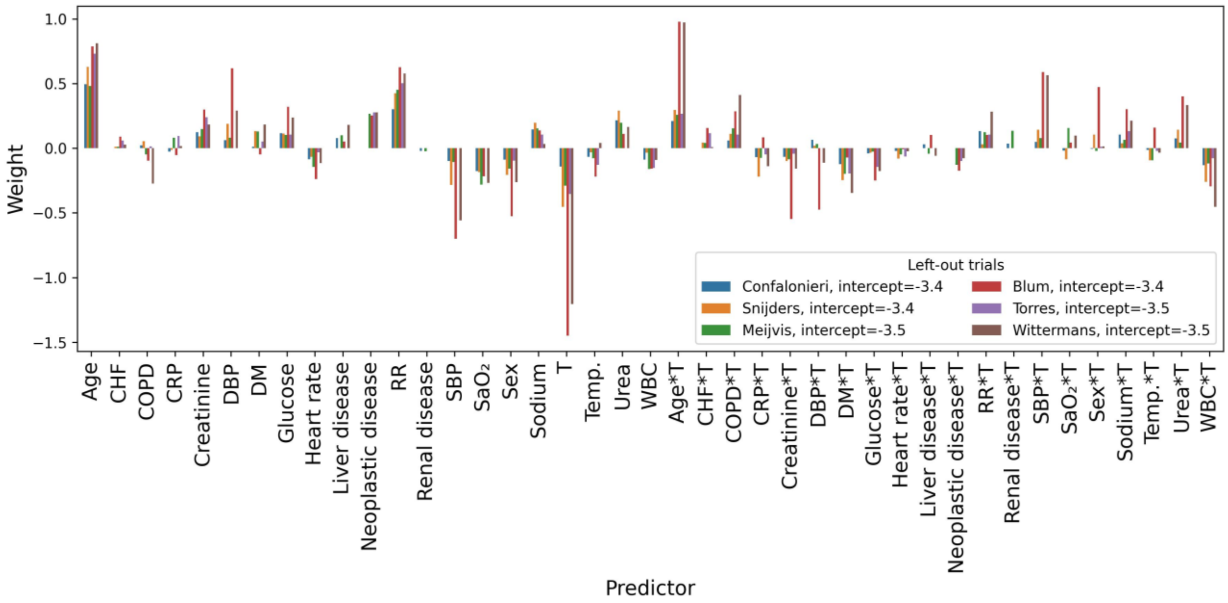

(e) Effect-2, unpenalized

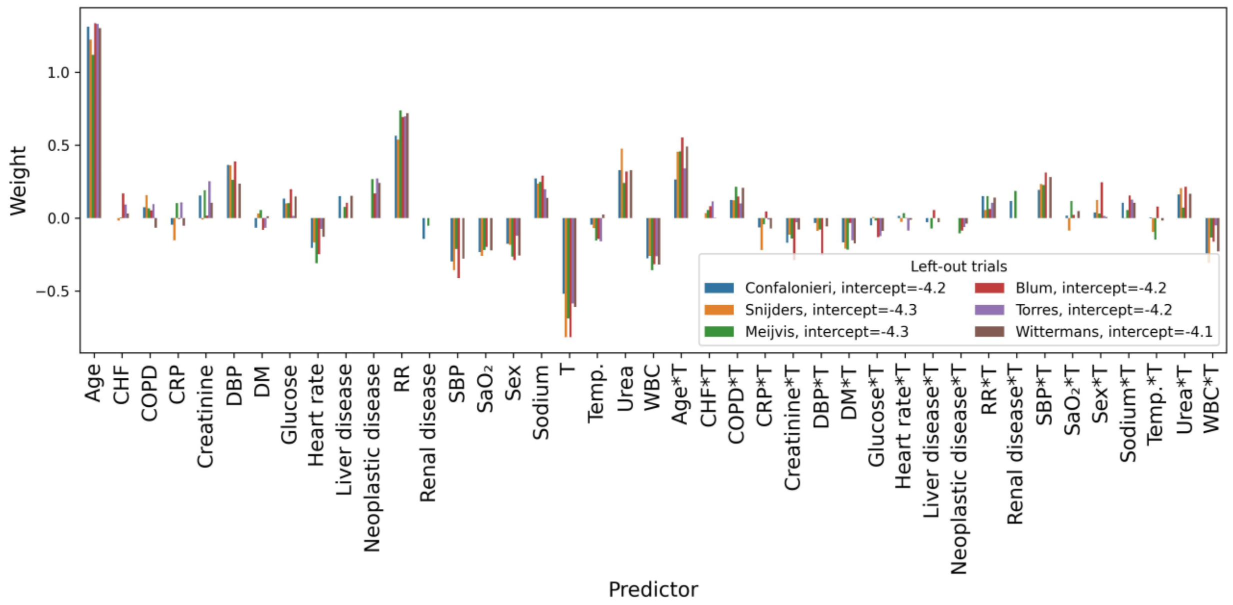

(f) Effect-2, Lasso

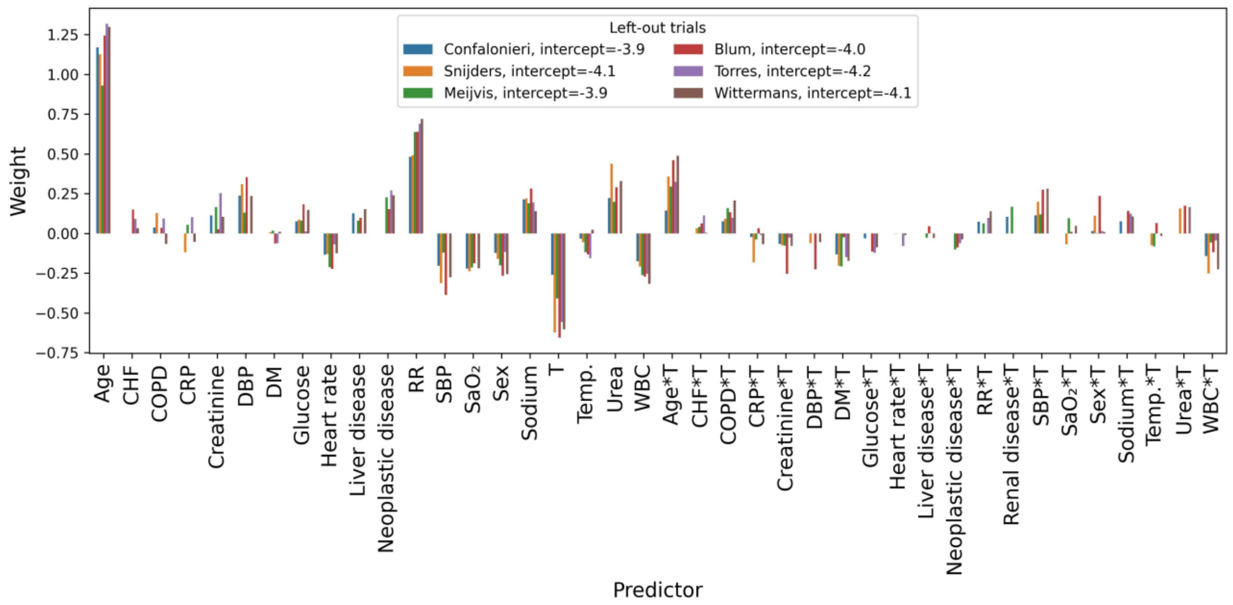

(g) Effect-2, Ridge

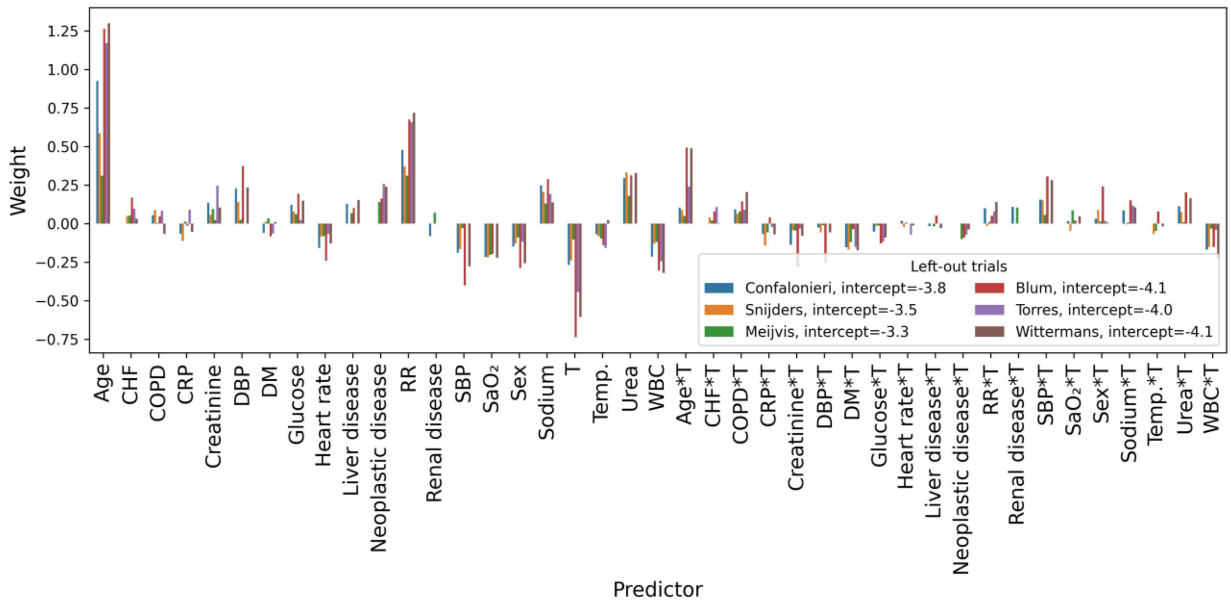

(h) Effect-3, unpenalized

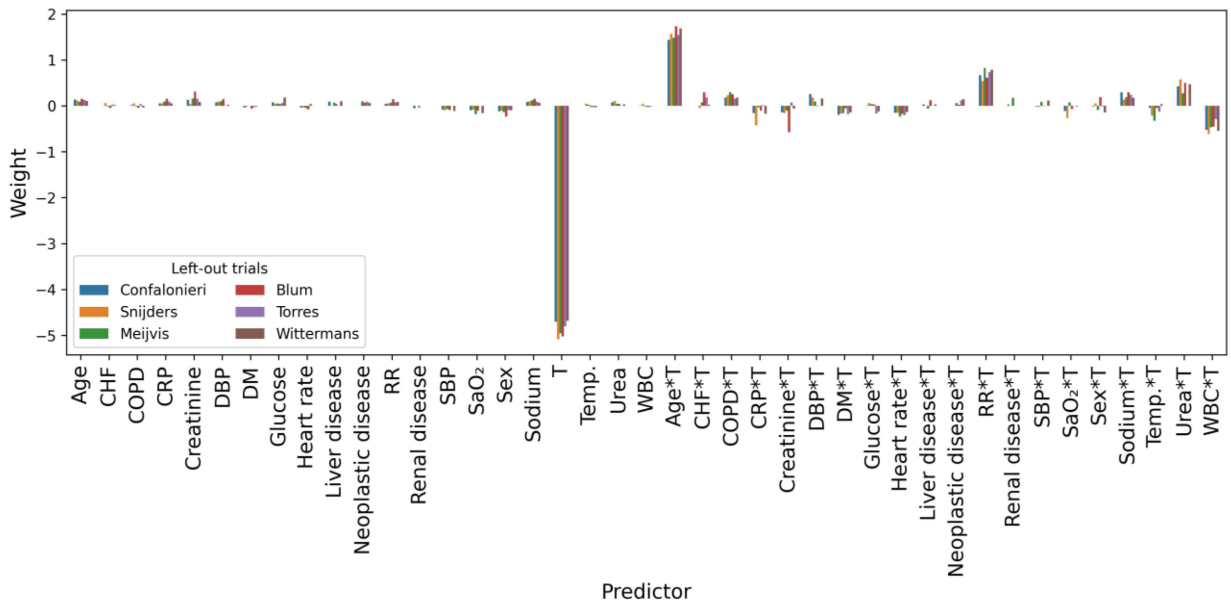

(i) Effect-3, Lasso

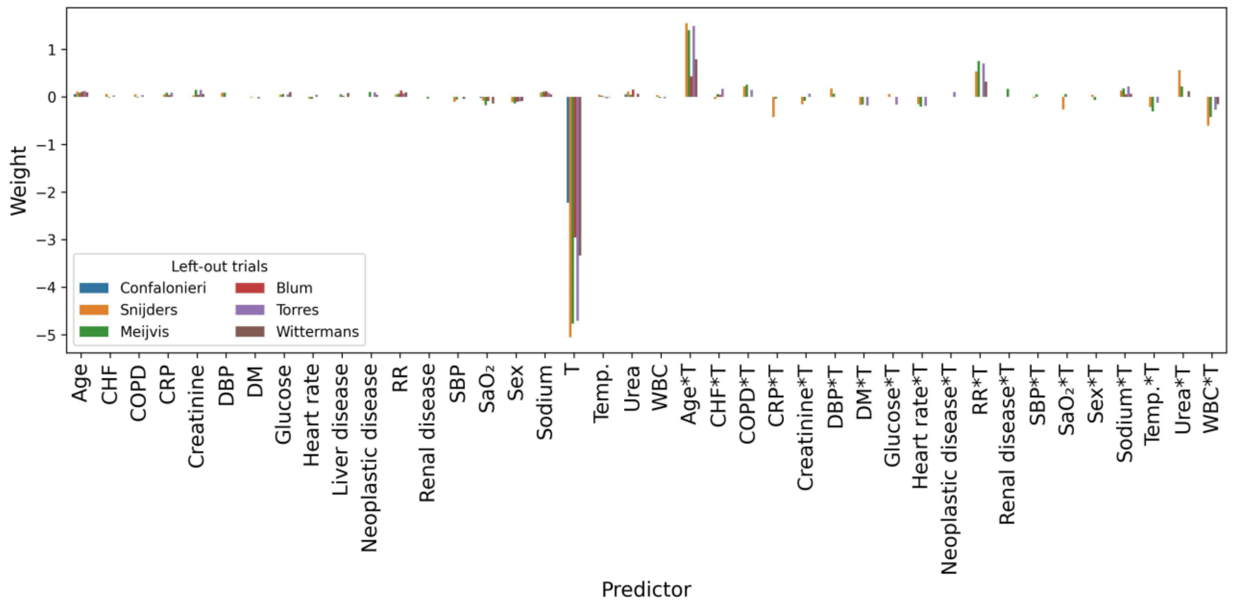

(j) Effect-3, Ridge

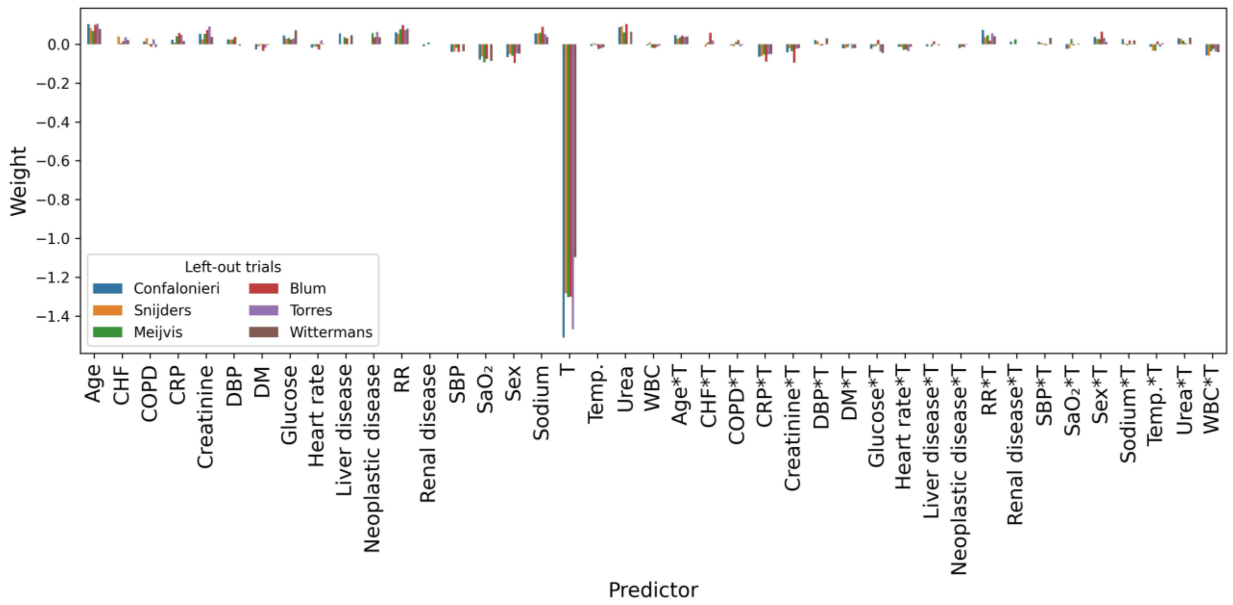

(k) Effect-4, unpenalized

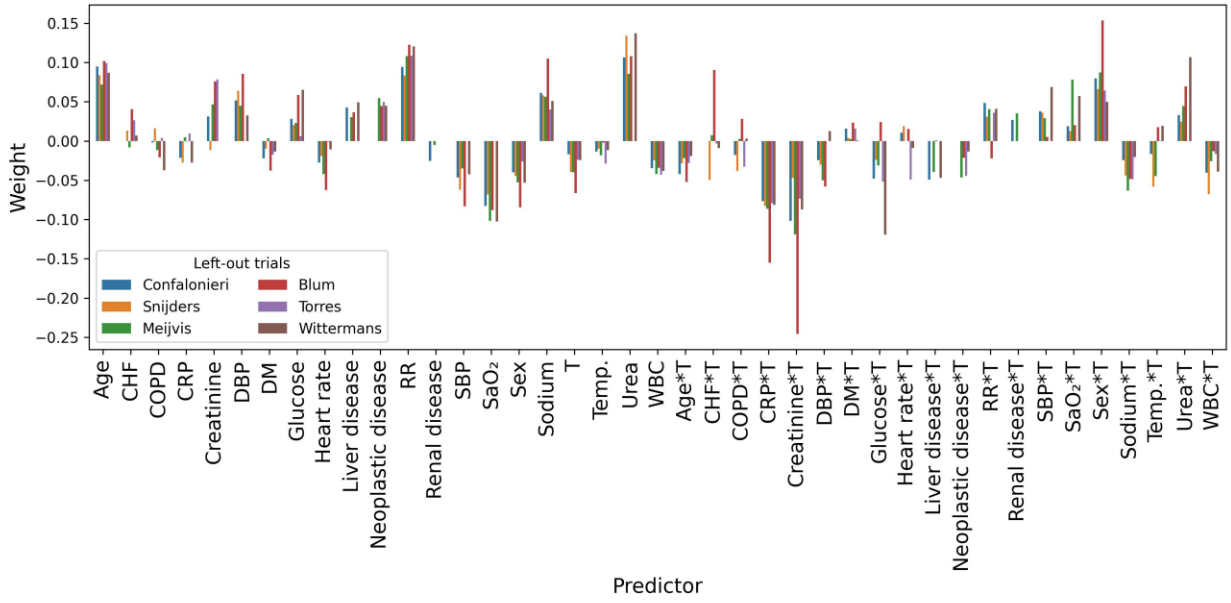

(l) Effect-4, Lasso

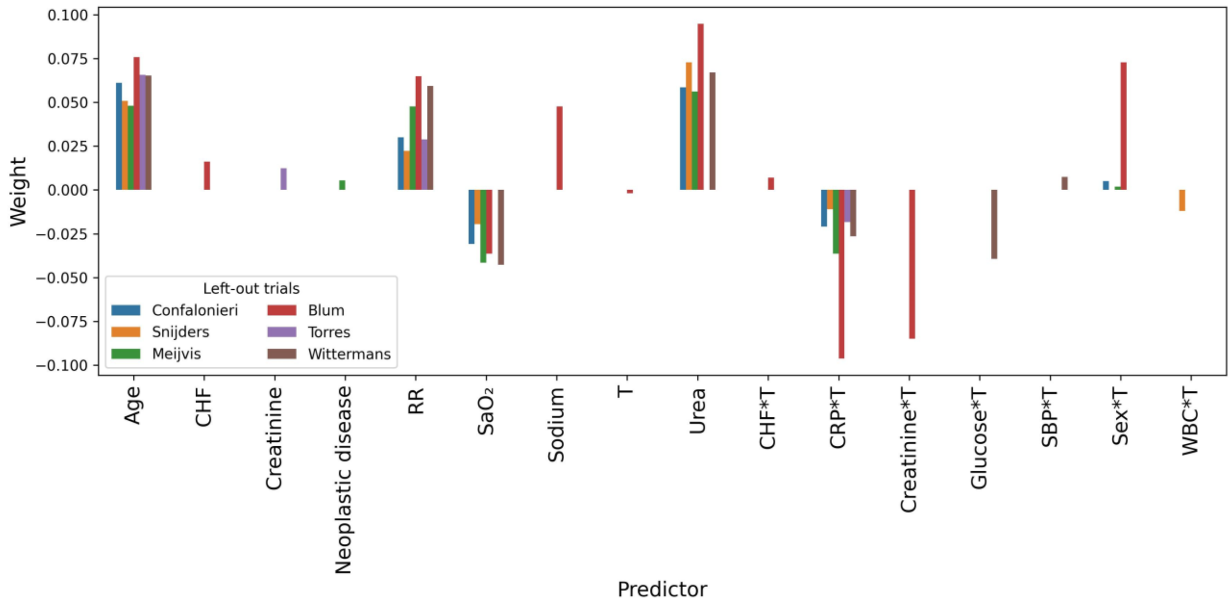

(m) Effect-4, Ridge

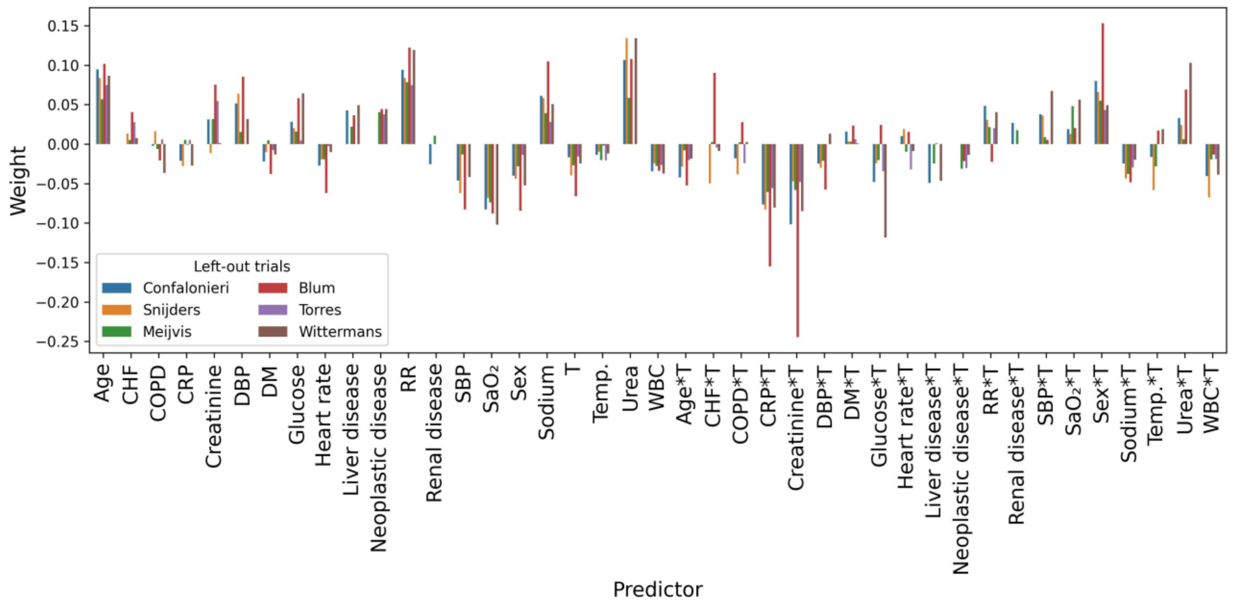

(n) Effect-5, unpenalized

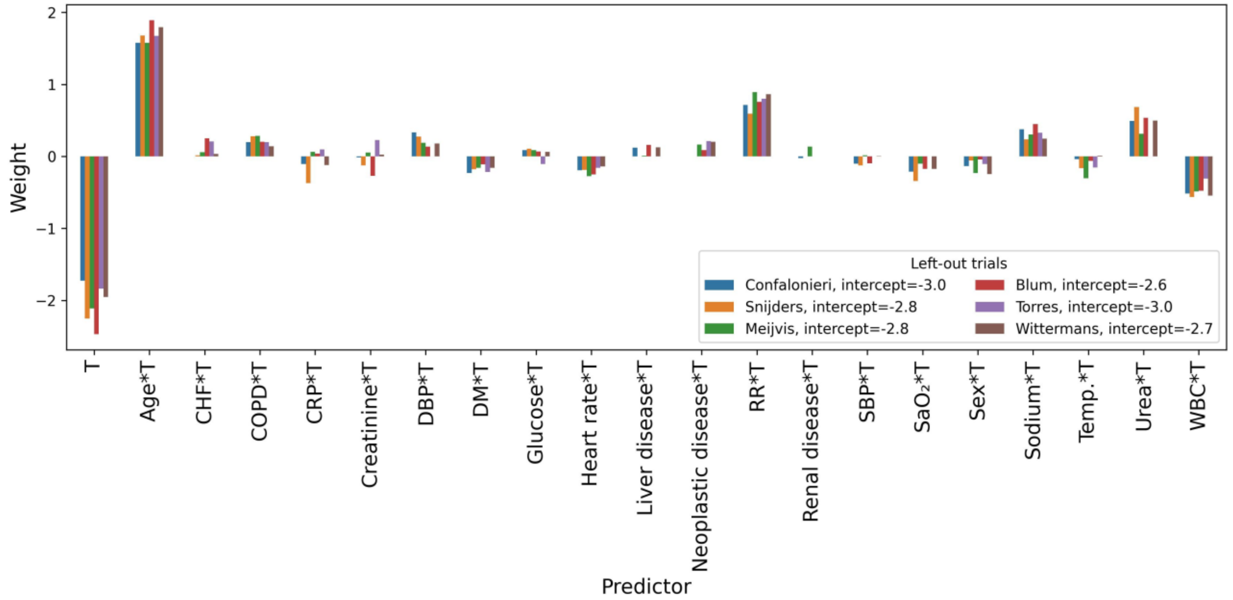

(o) Effect-5, Lasso

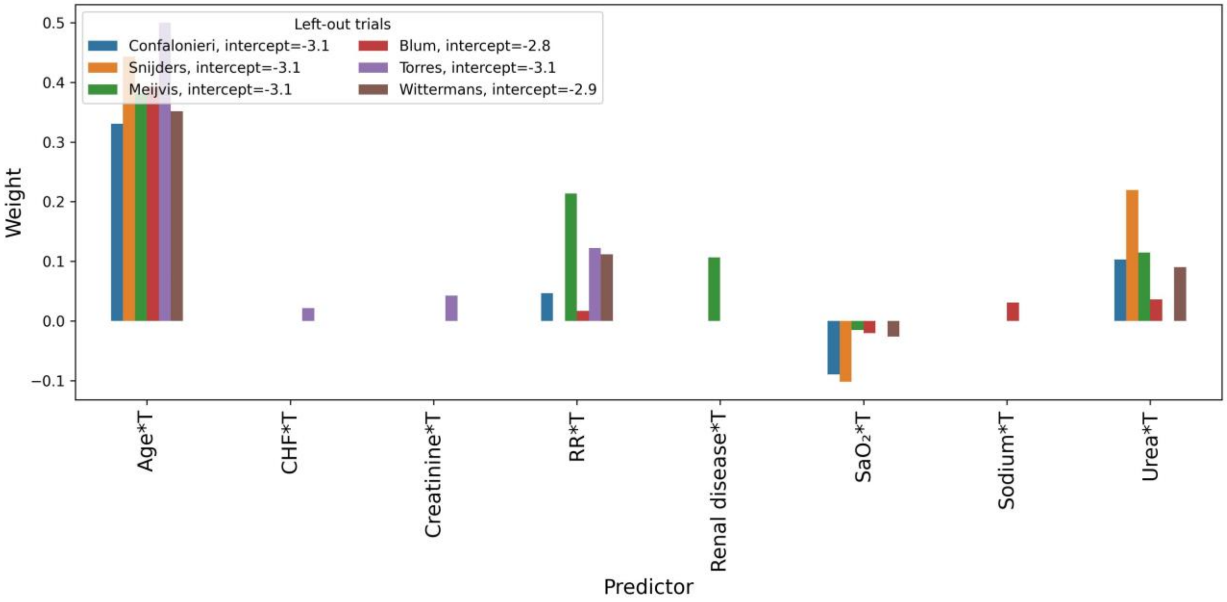

(p) Effect-5, Ridge

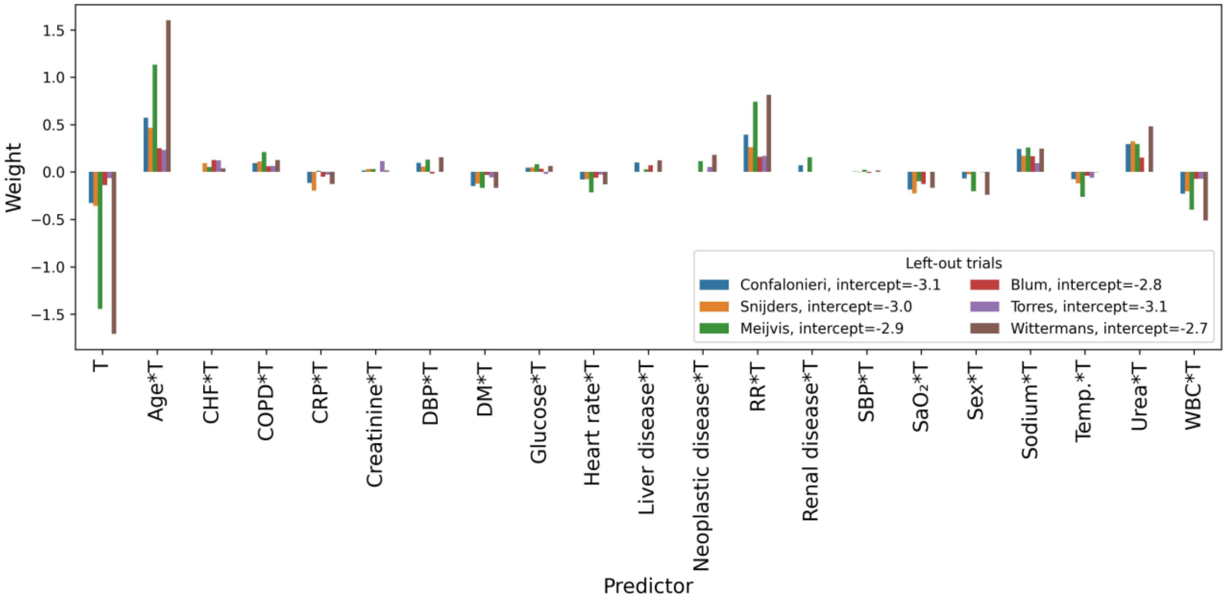

(q) Effect-6, unpenalized

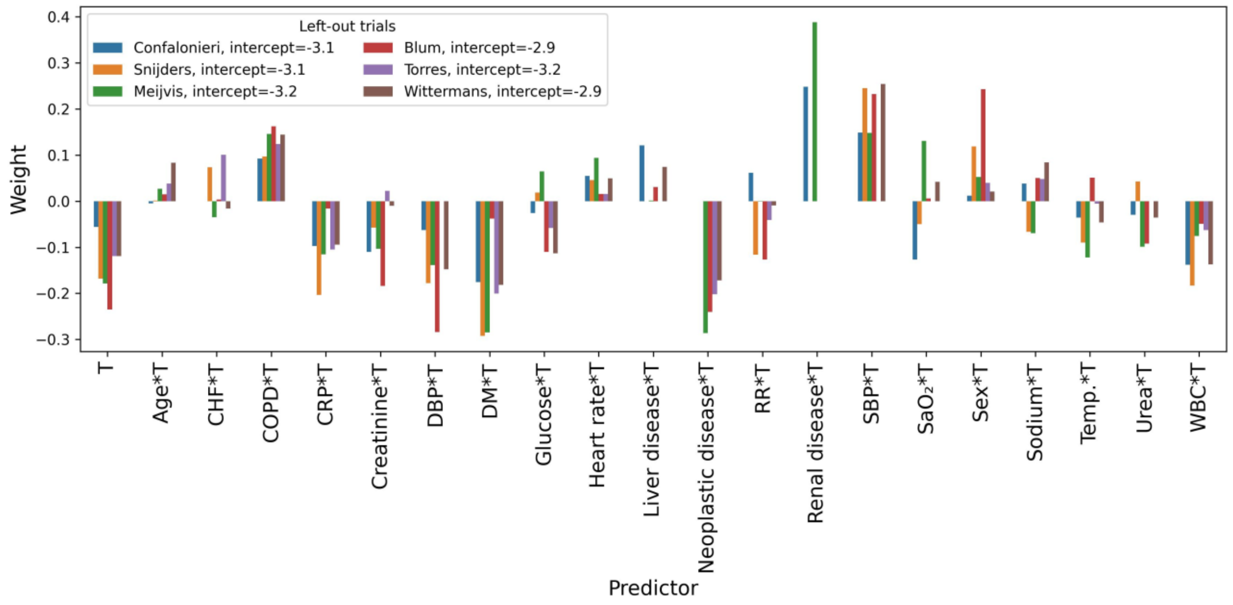

(r) Effect-6, Lasso

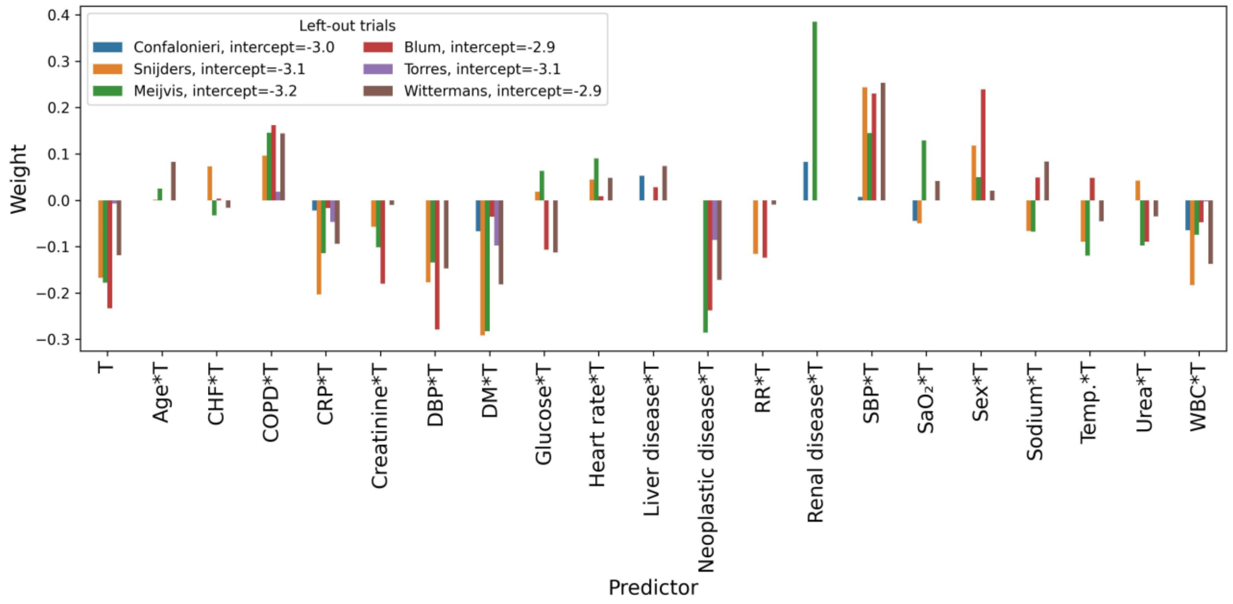

(s) Effect-6, Ridge

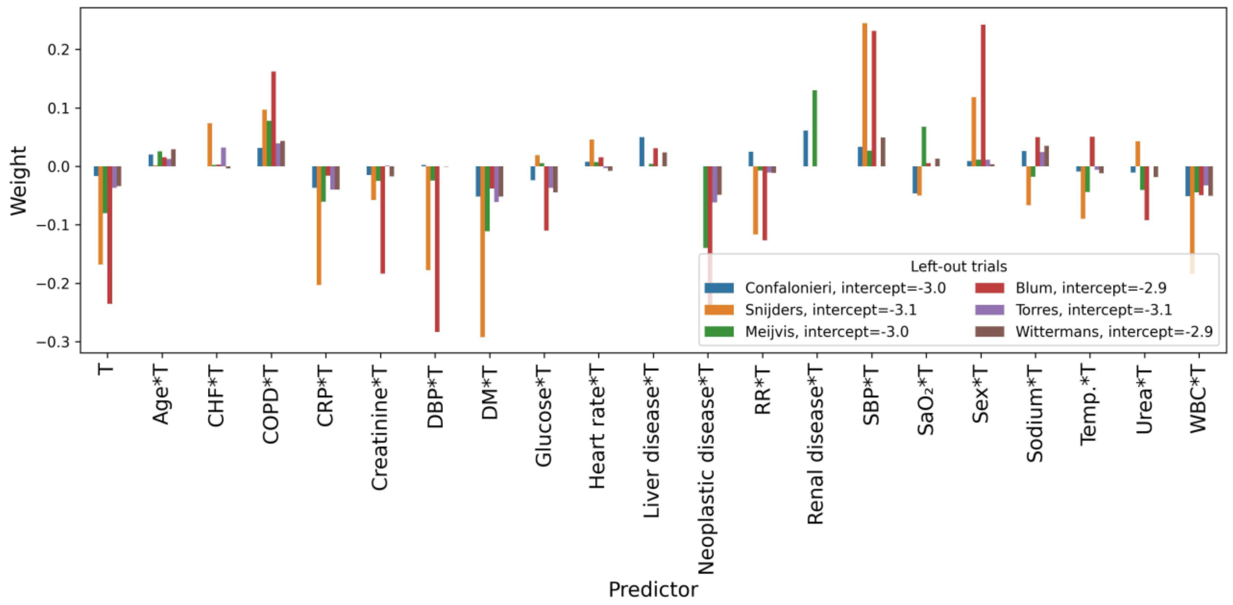

(t) Effect-7, unpenalized

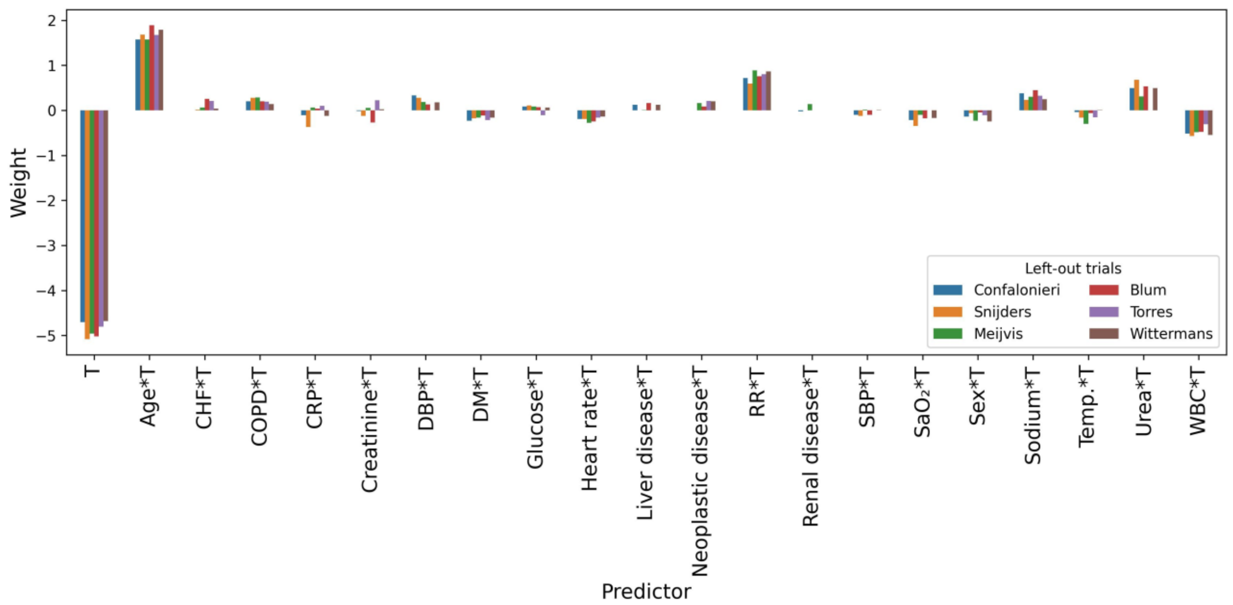

(u) Effect-7, Lasso

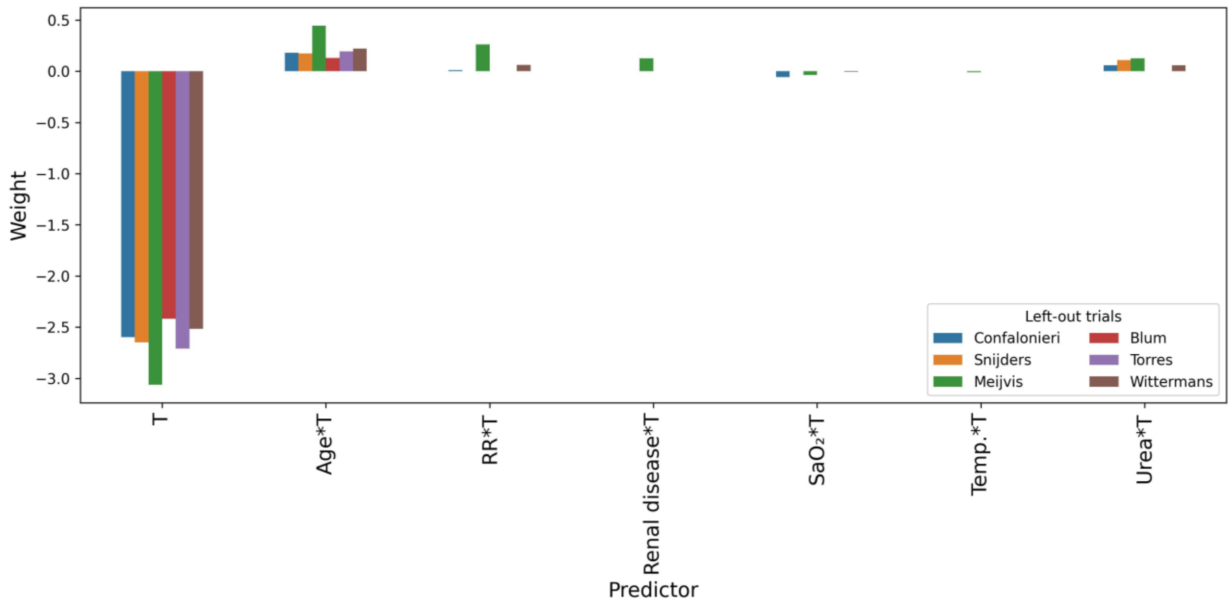

(v) Effect-7, Ridge

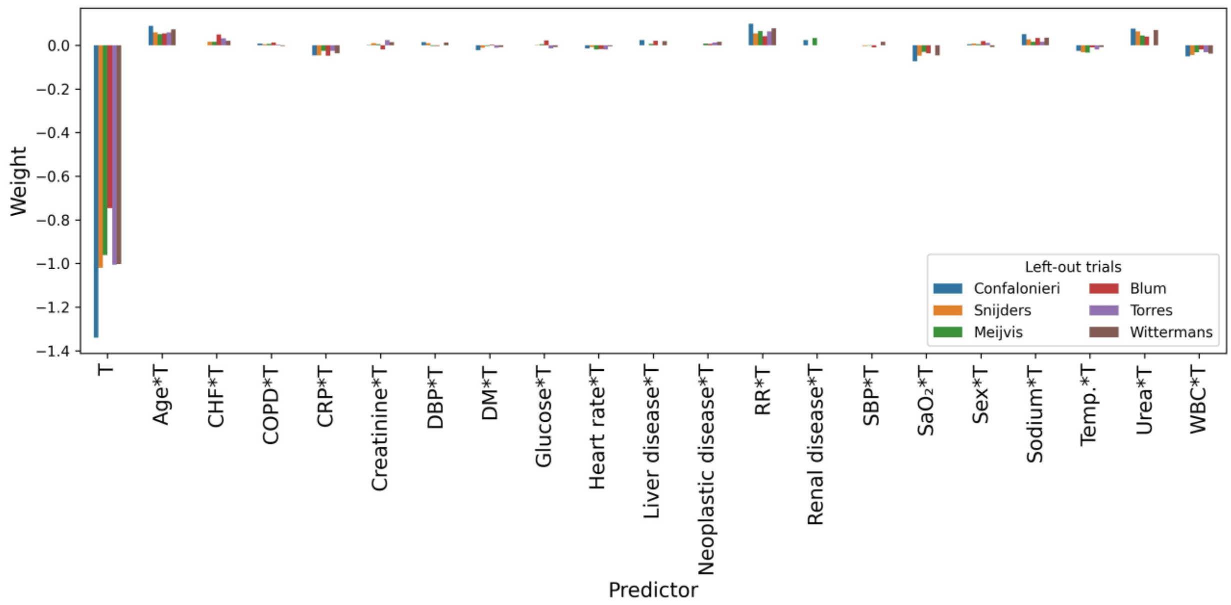

(w) Effect-8, unpenalized

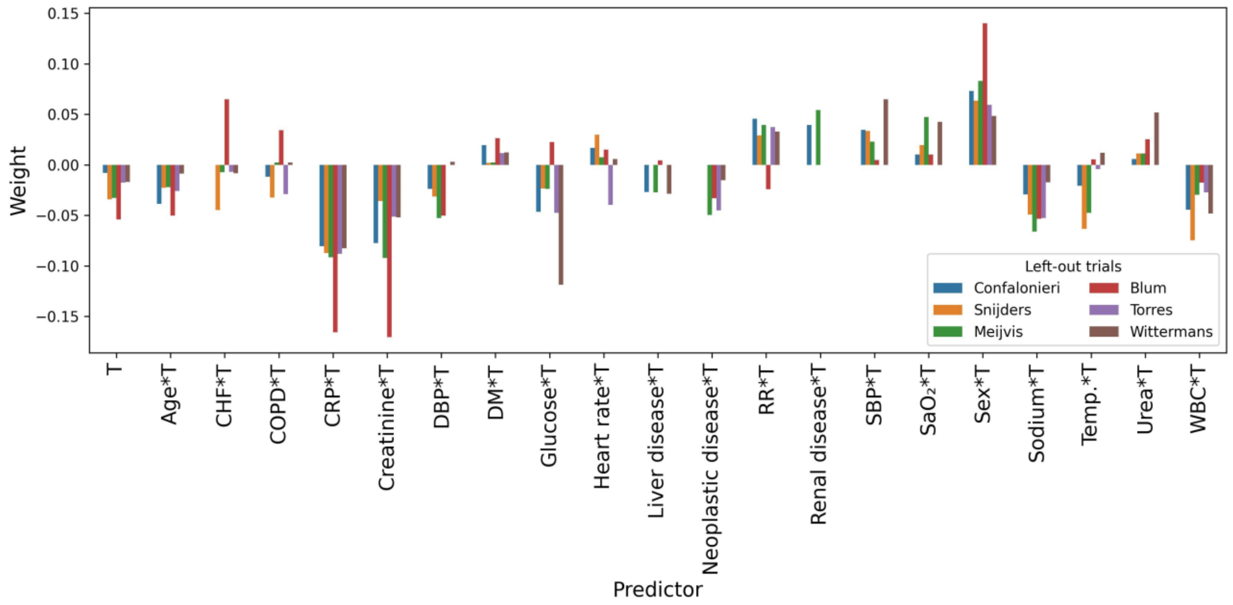

(x) Effect-8, Lasso (‘procedure 1’ in the main text)

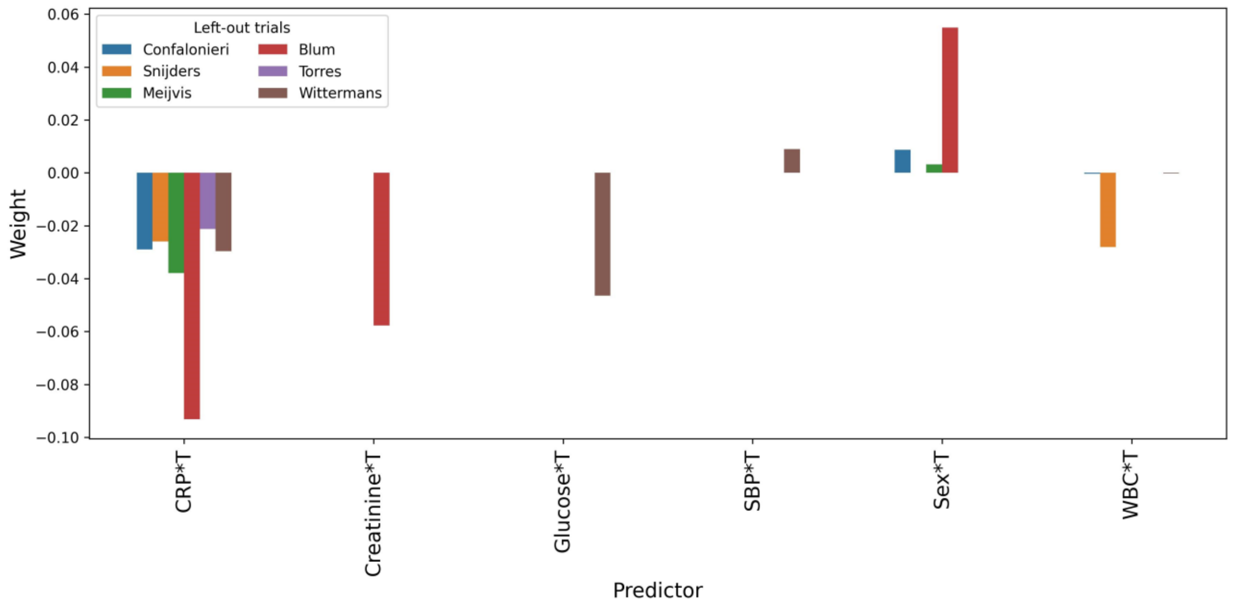

(y) Effect-8, Ridge

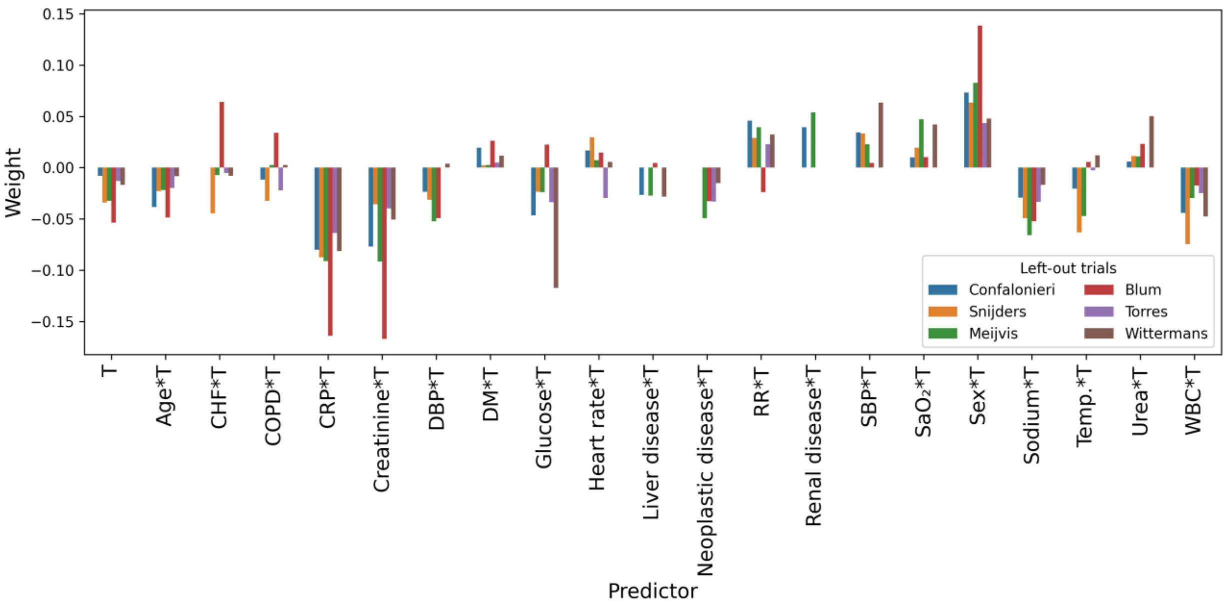

Figure D4: Bar charts of all non-zero weights of the fitted logistic regression models in each LOTO-CV fold, resulting from the different variations of modelling procedures with additional dichotomized variables. RR=respiratory rate, DBP=Diastolic blood pressure, SBP=Systolic blood pressure, Temp.=Body temperature, CRP=C-reactive protein, WBC=White cell count, T=Treatment variable

(a) Risk

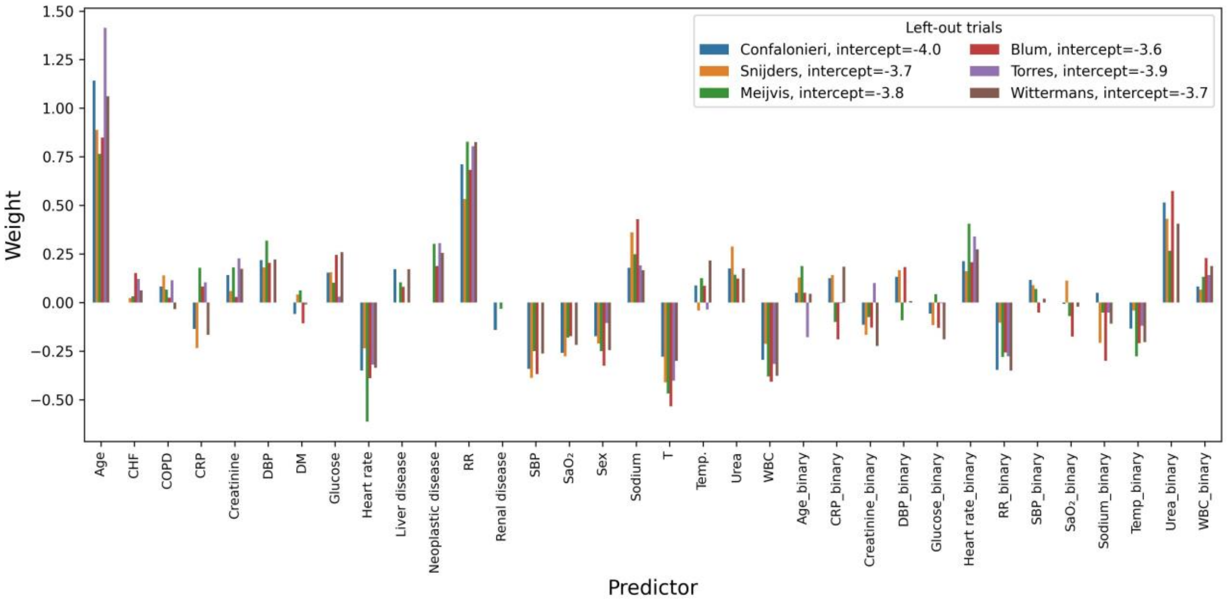

(b) Effect-1, unpenalized

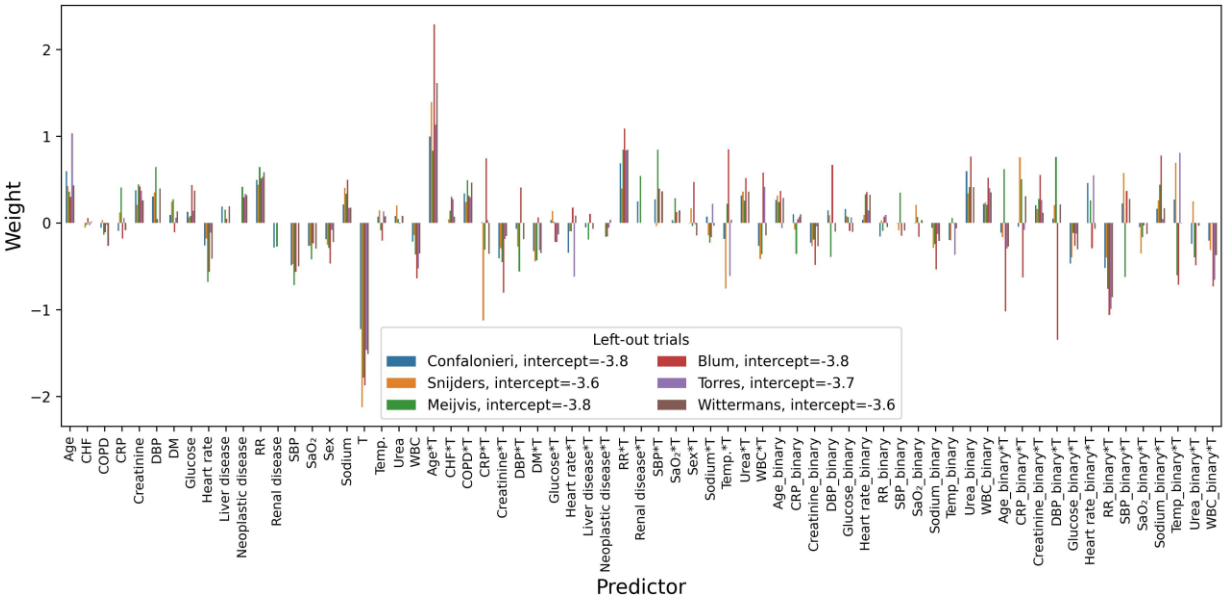

(c) Effect-1, Lasso

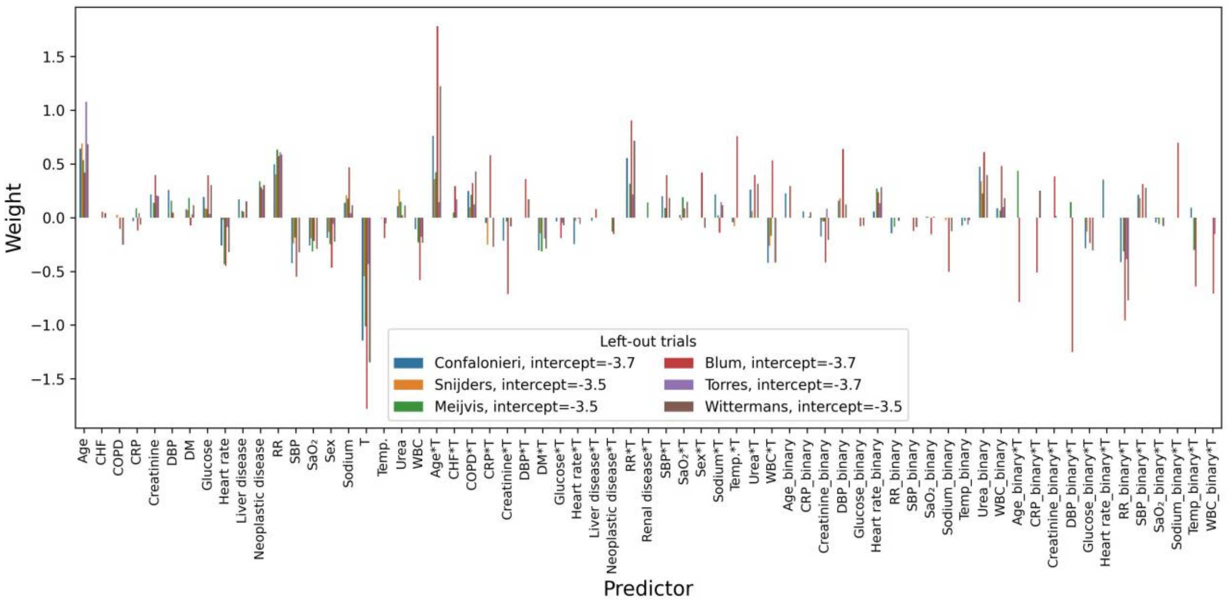

(d) Effect-1, Ridge

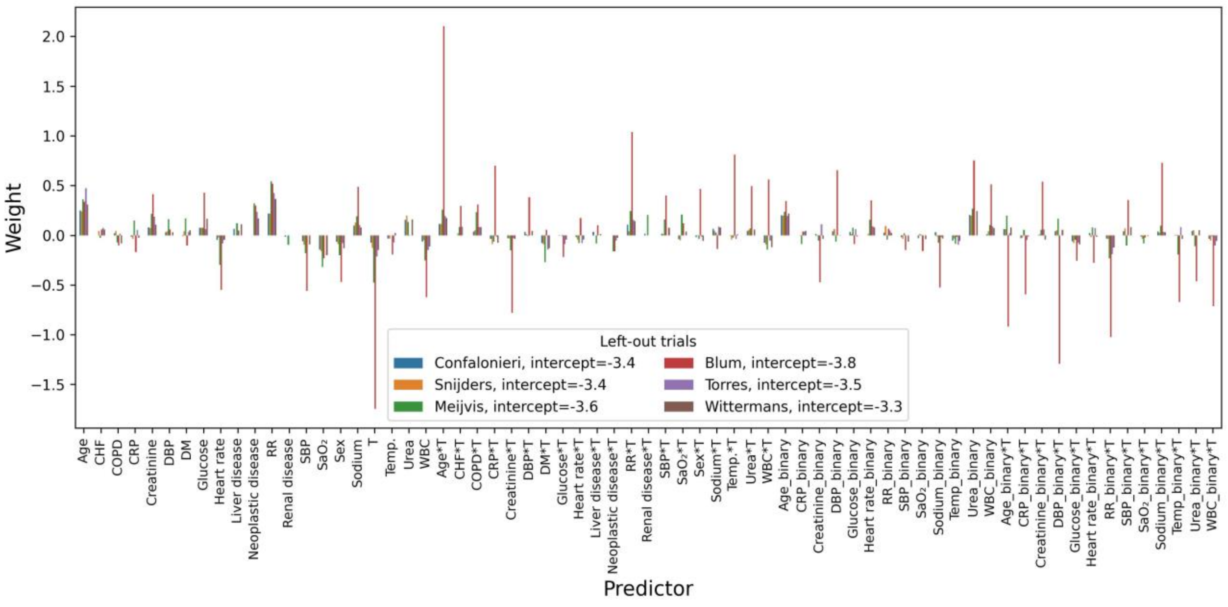

(e) Effect-2, unpenalized

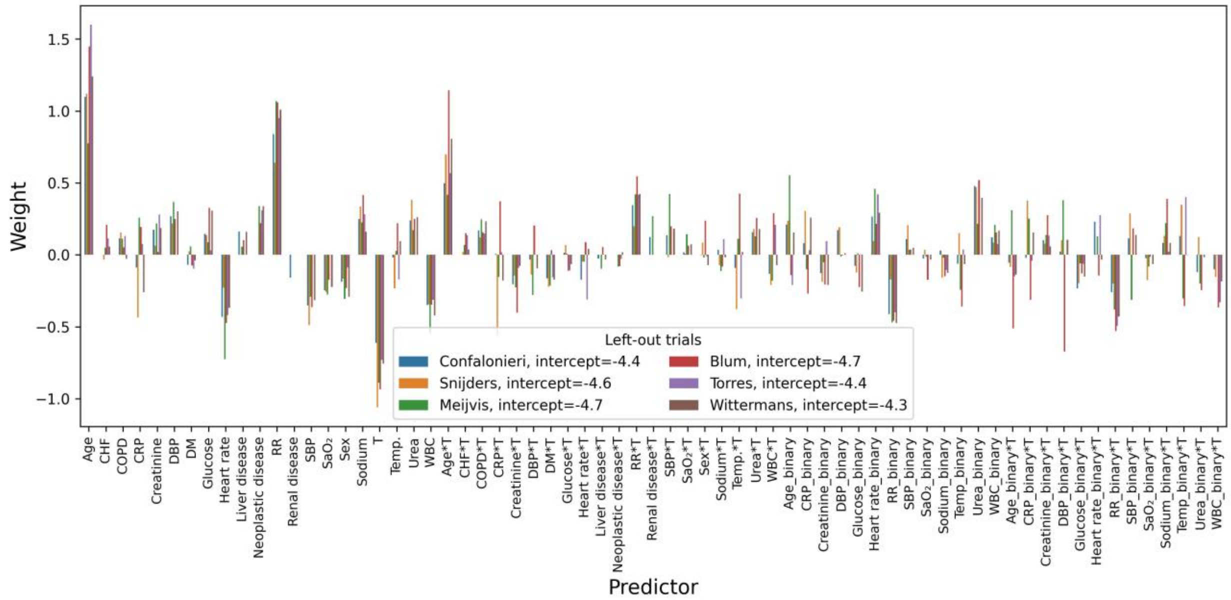

(f) Effect-2, Lasso

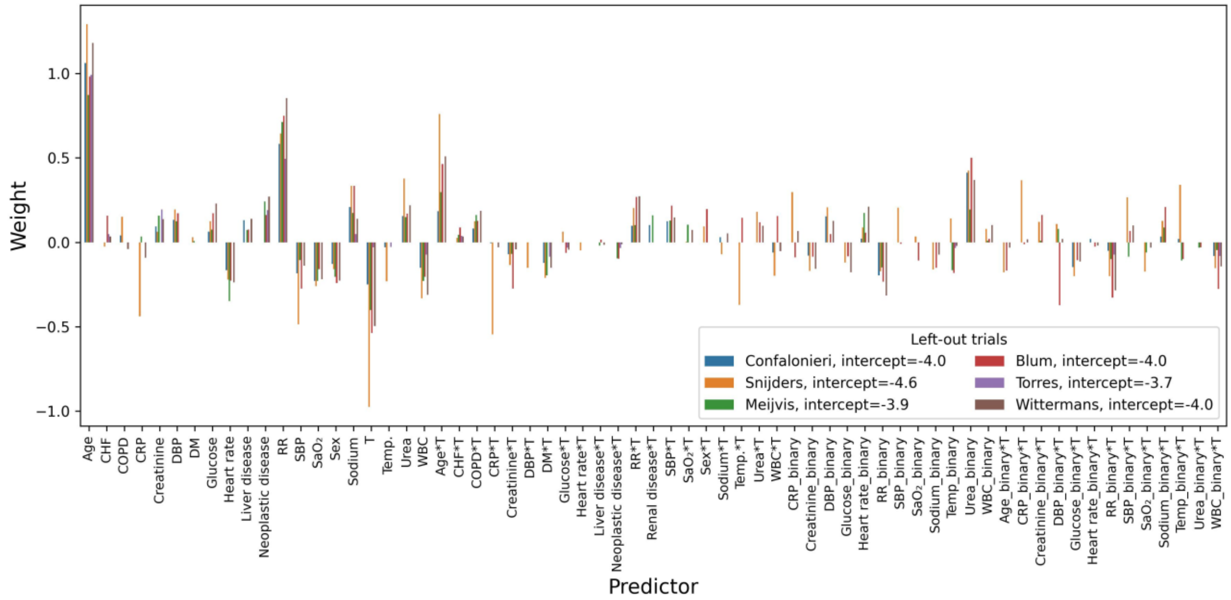

(g) Effect-2, Ridge

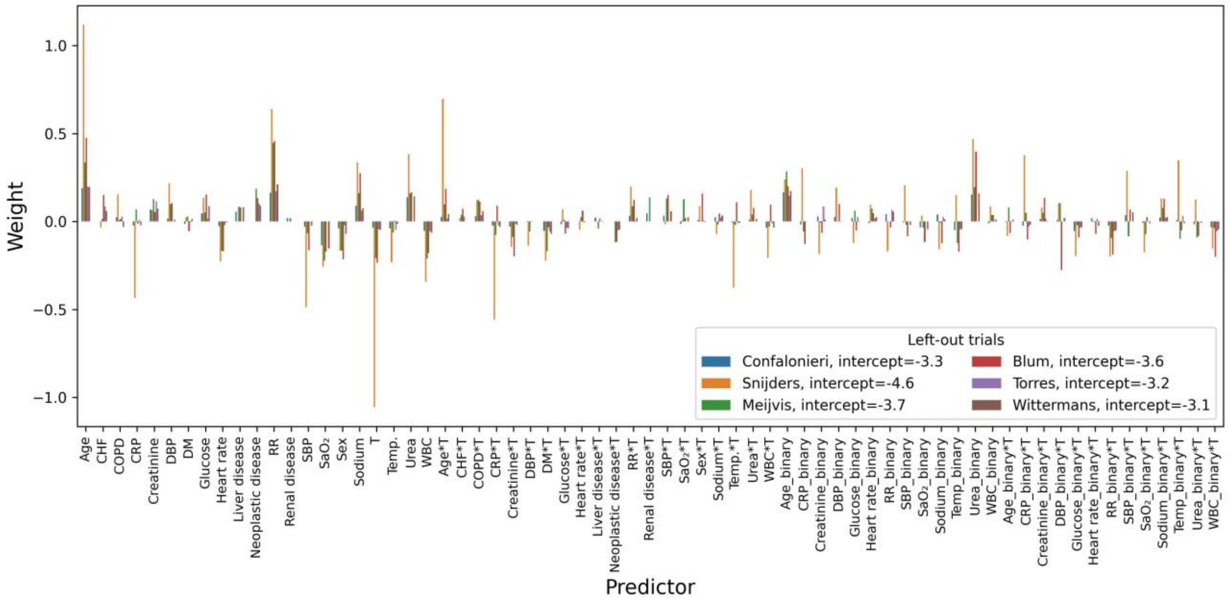

(h) Effect-3, unpenalized

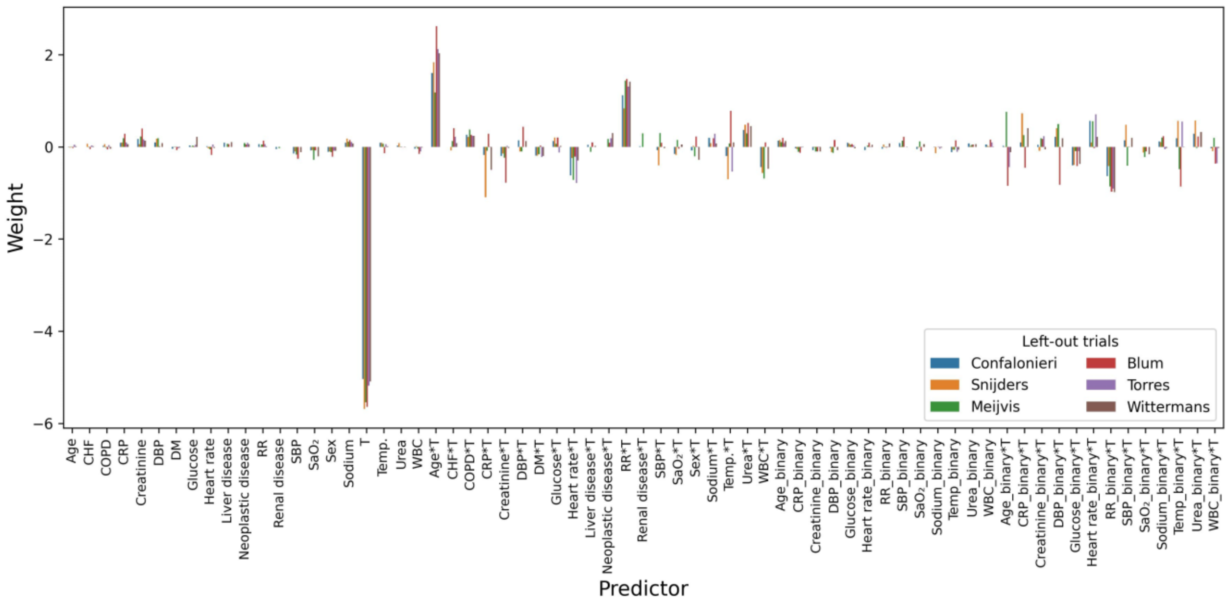

(i) Effect-3, Lasso

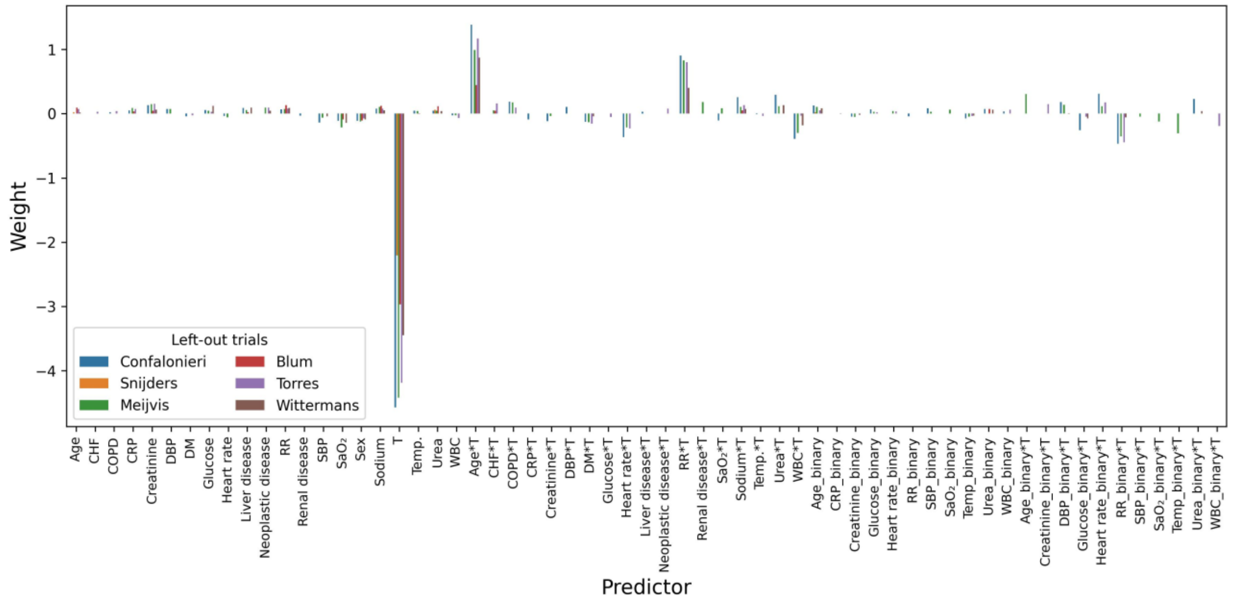

(j) Effect-3, Ridge

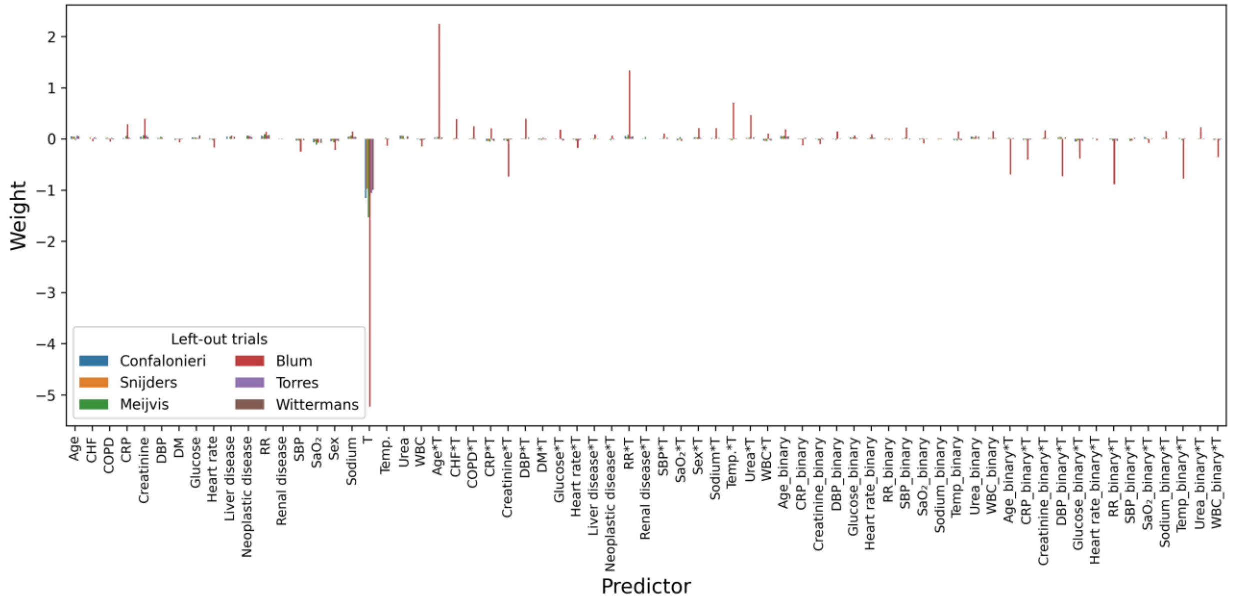

(k) Effect-4, unpenalized

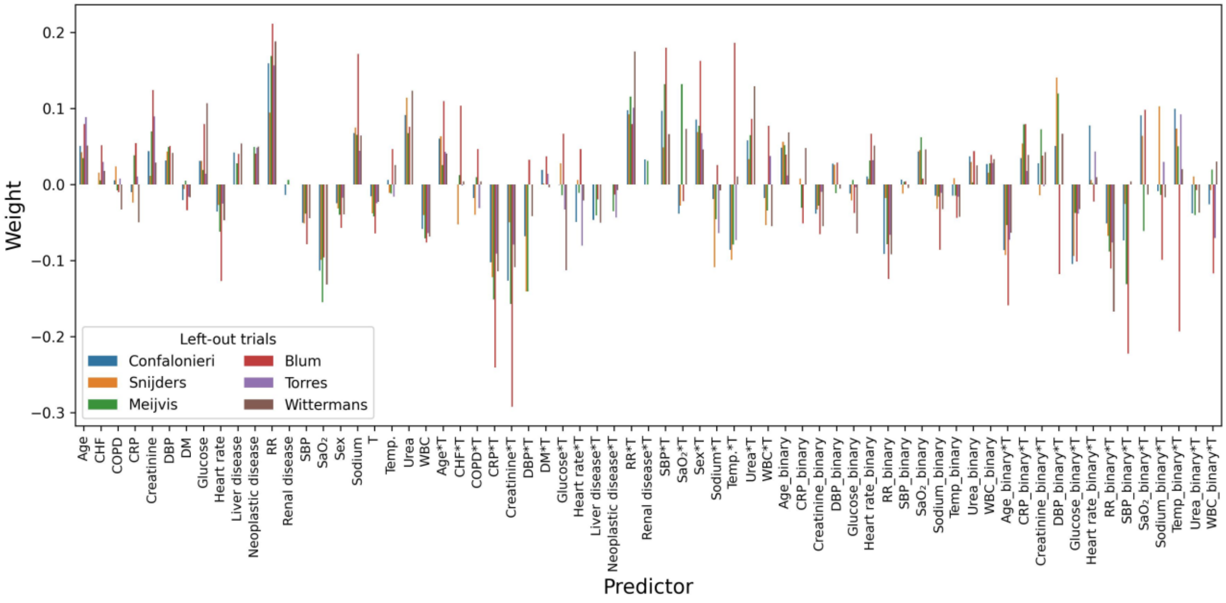

(l) Effect-4, Lasso

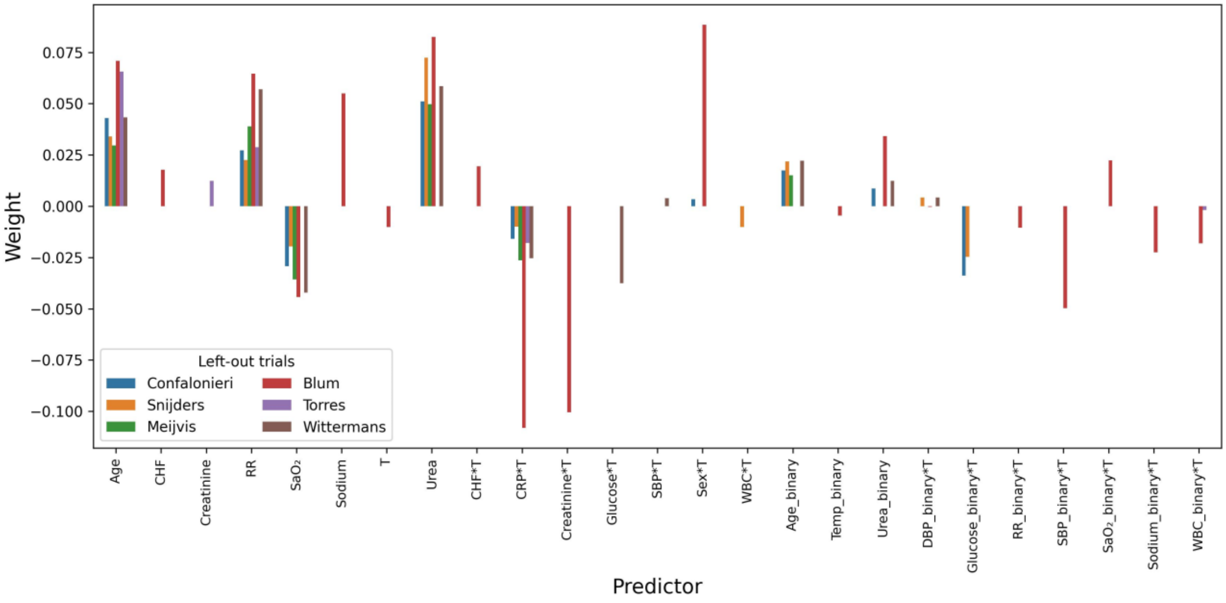

(m) Effect-4, Ridge

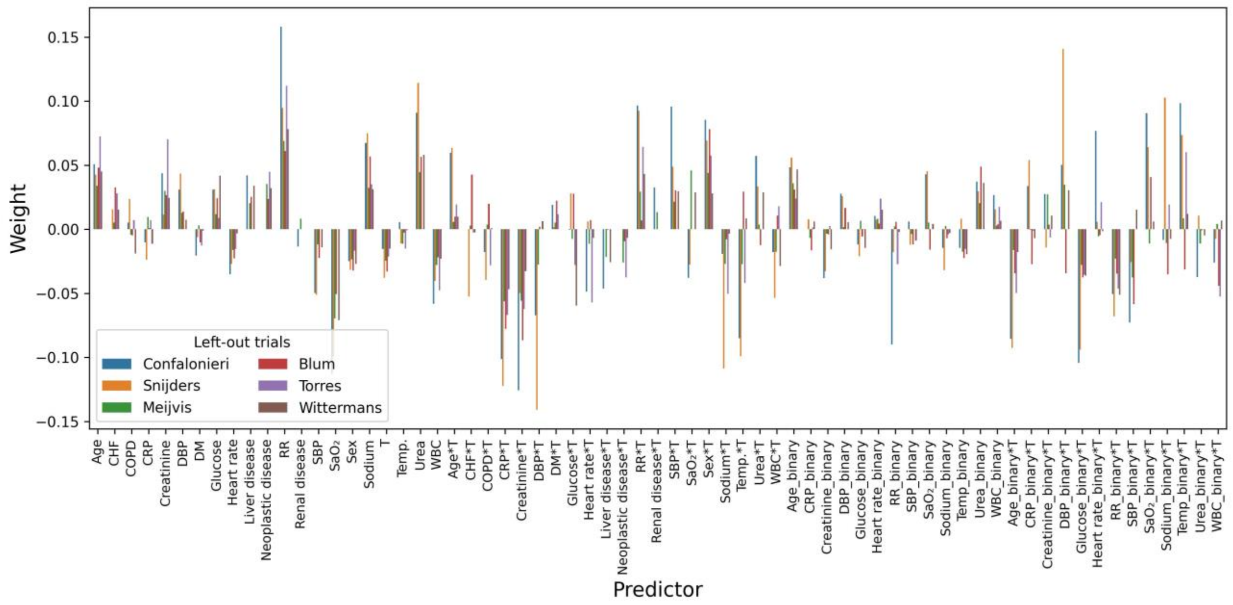

(n) Effect-5, unpenalized

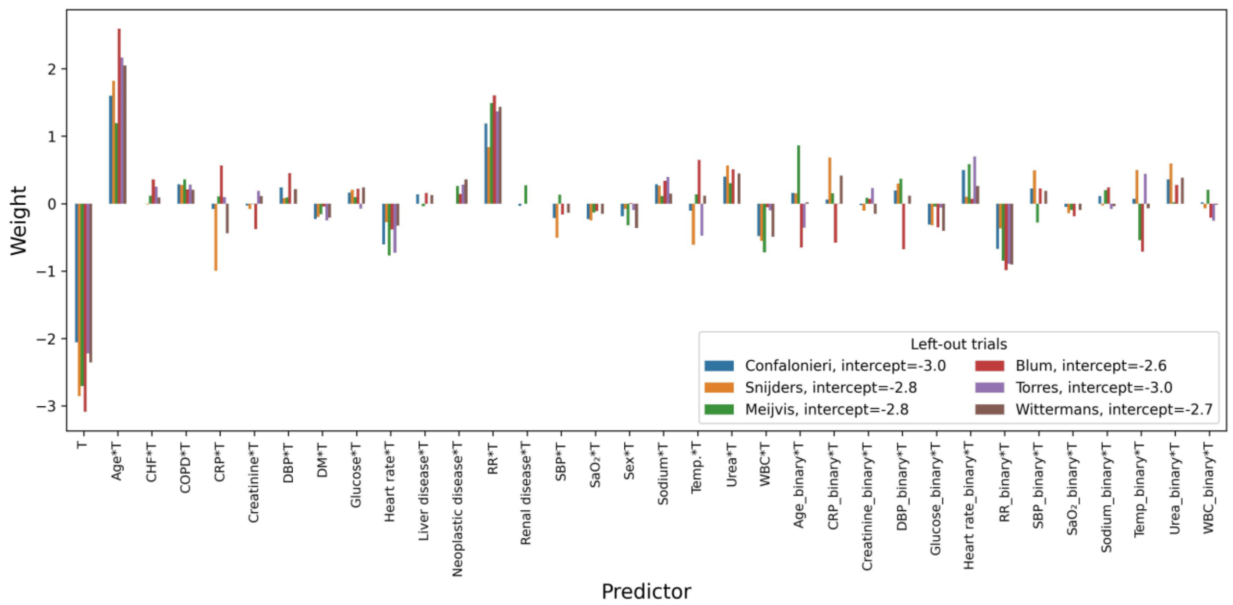

(o) Effect-5, Lasso

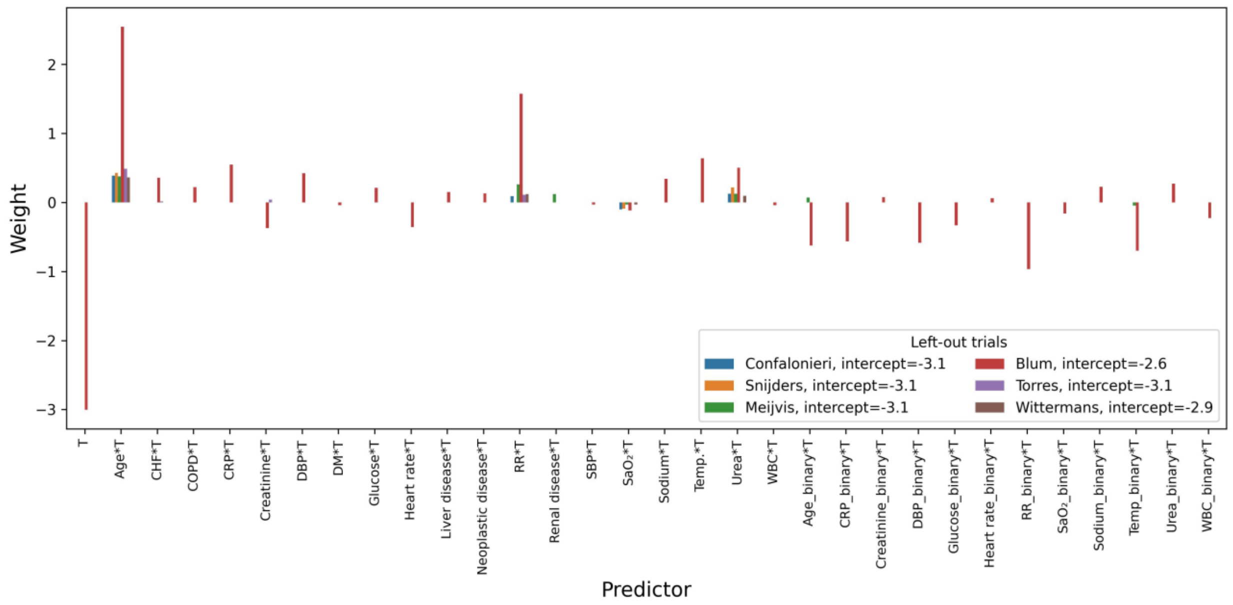

(p) Effect-5, Ridge

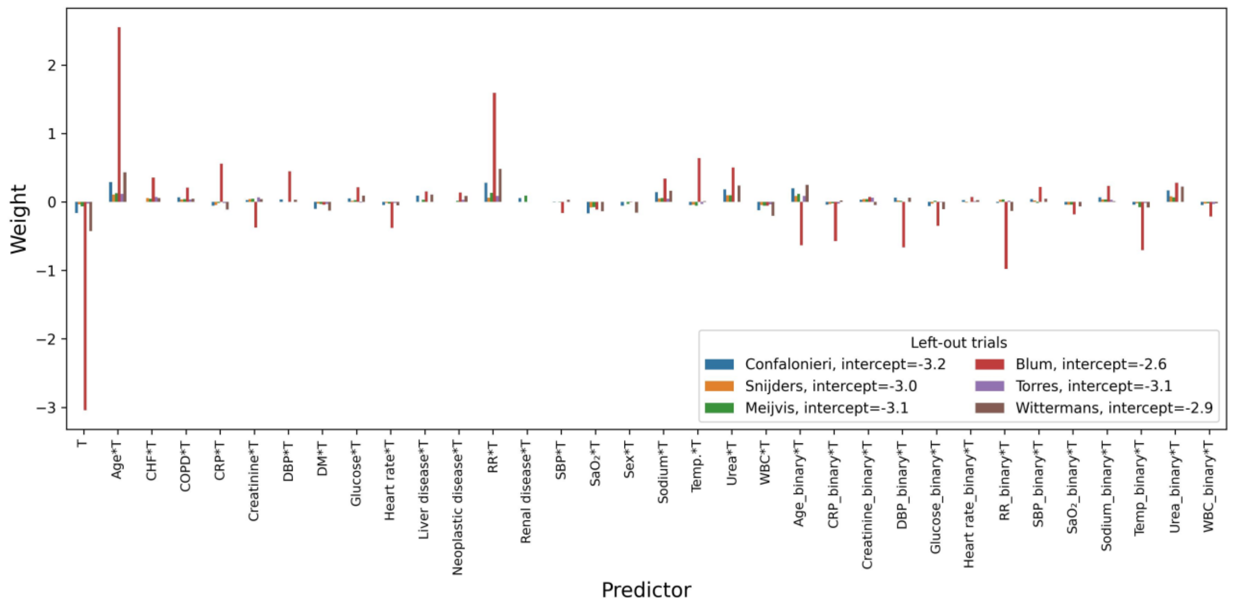

(q) Effect-6, unpenalized

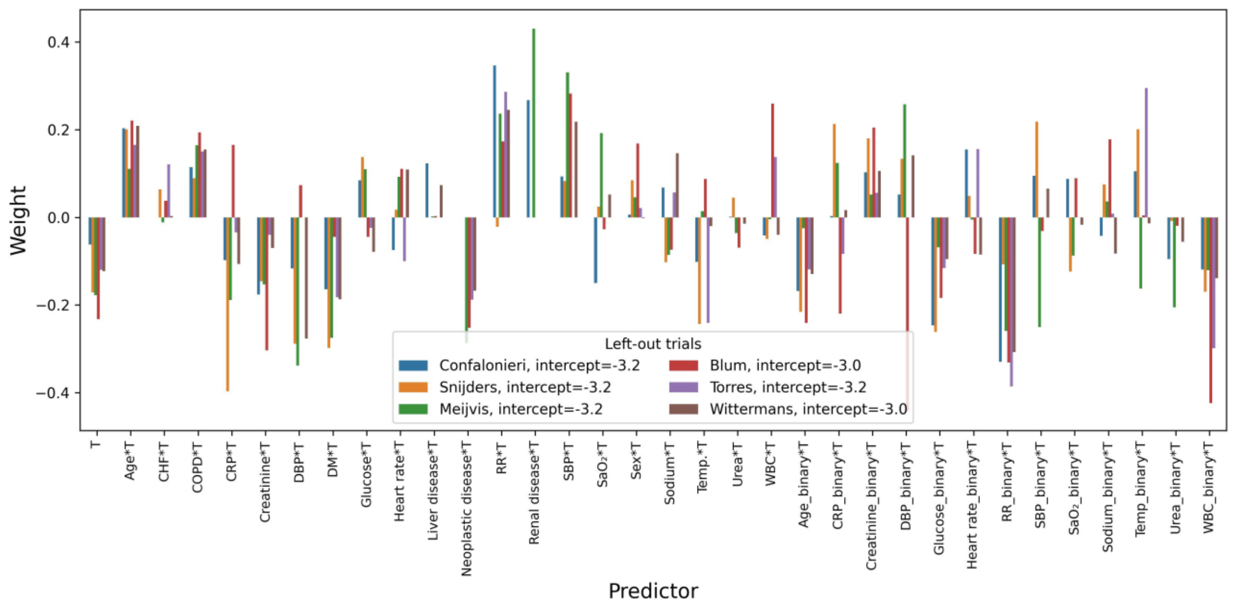

(r) Effect-6, Lasso

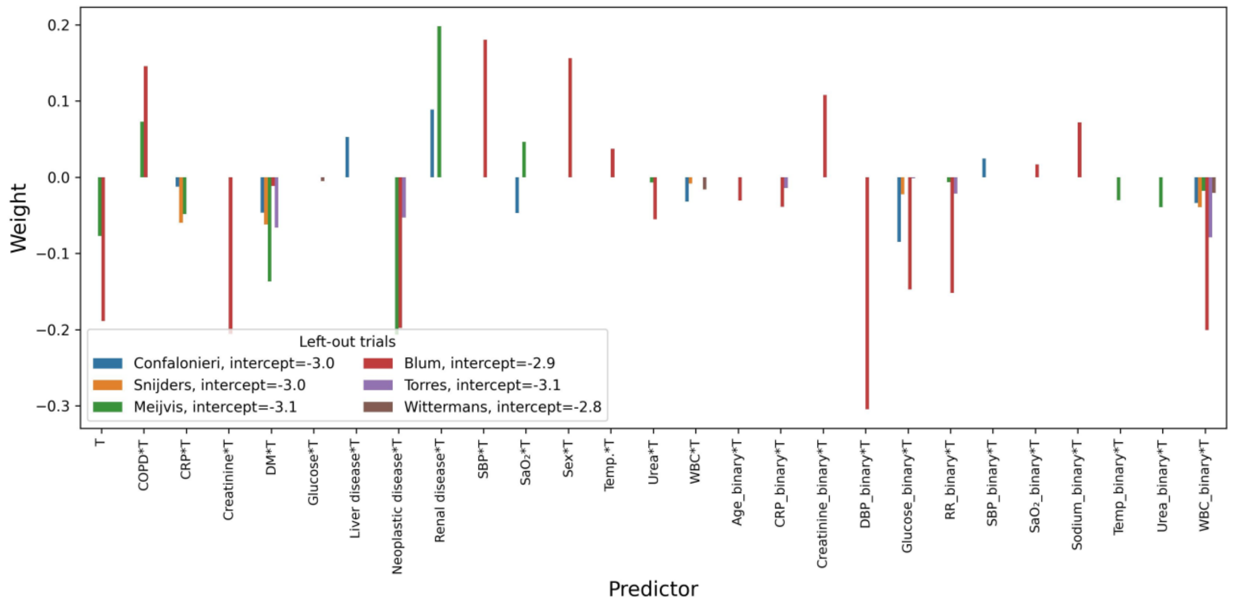

(s) Effect-6, Ridge

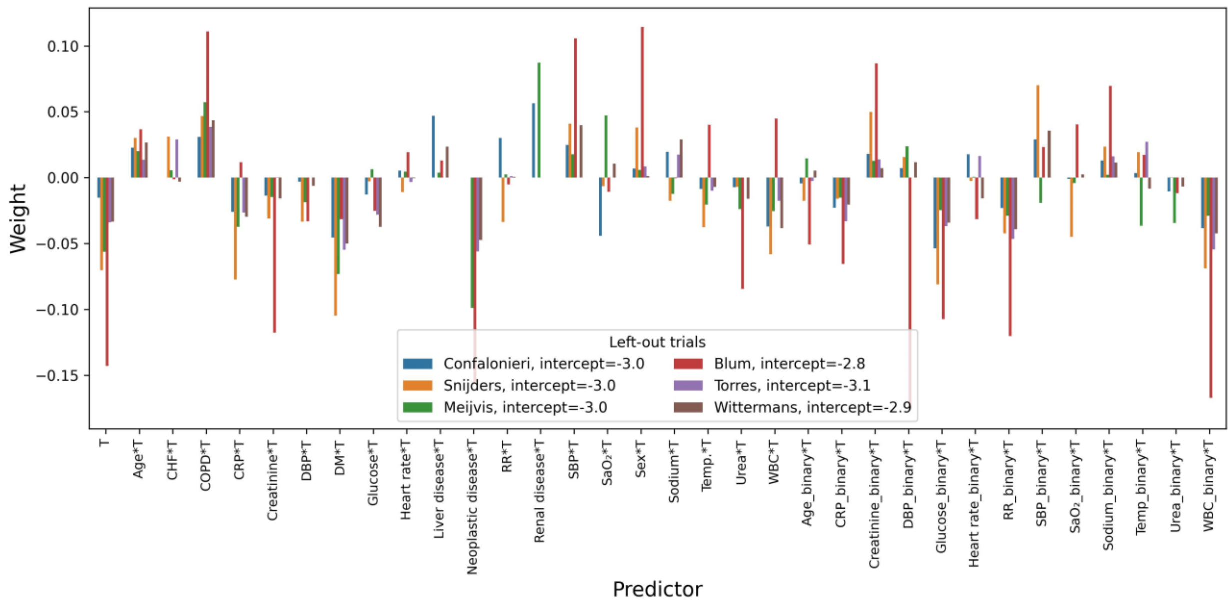

(t) Effect-7, unpenalized

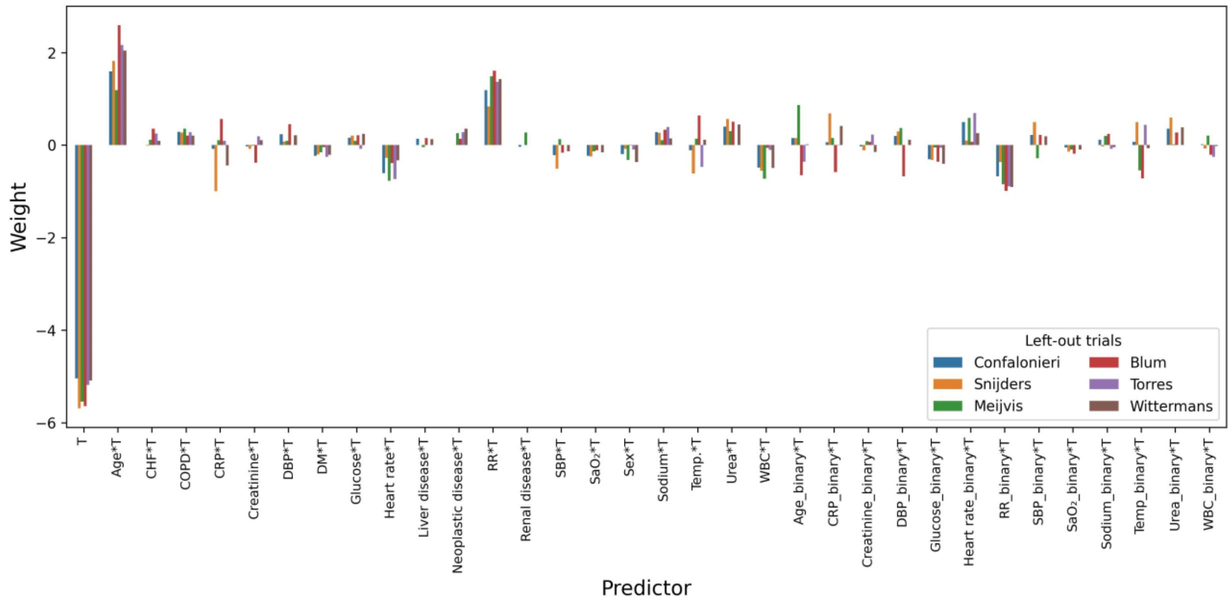

(u) Effect-7, Lasso

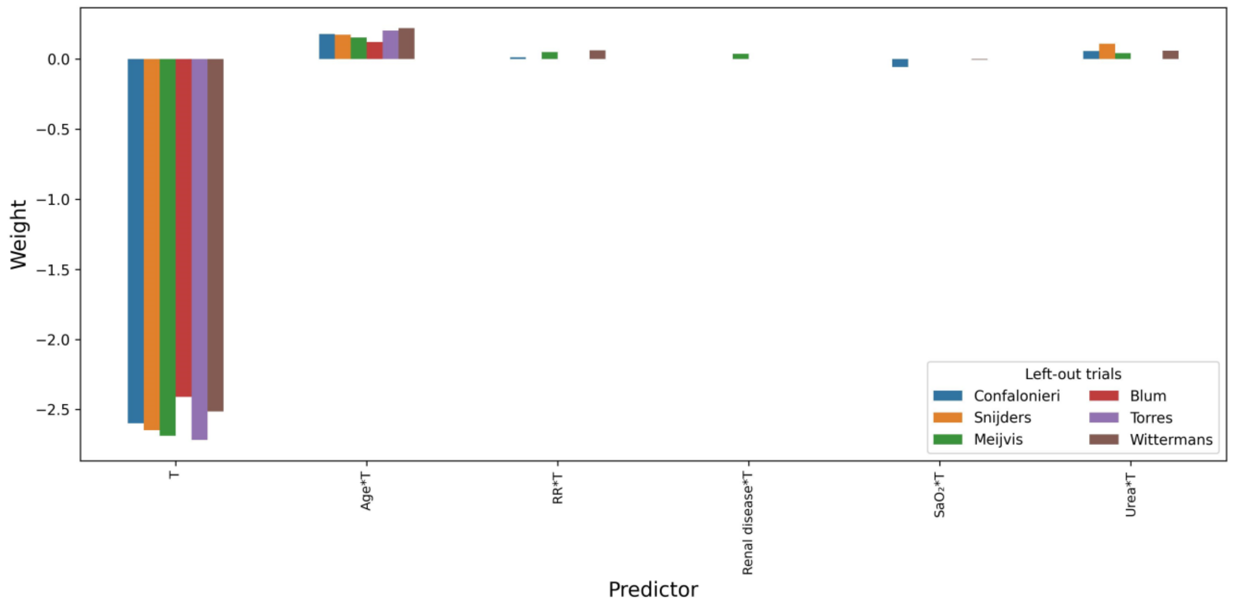

(v) Effect-7, Ridge

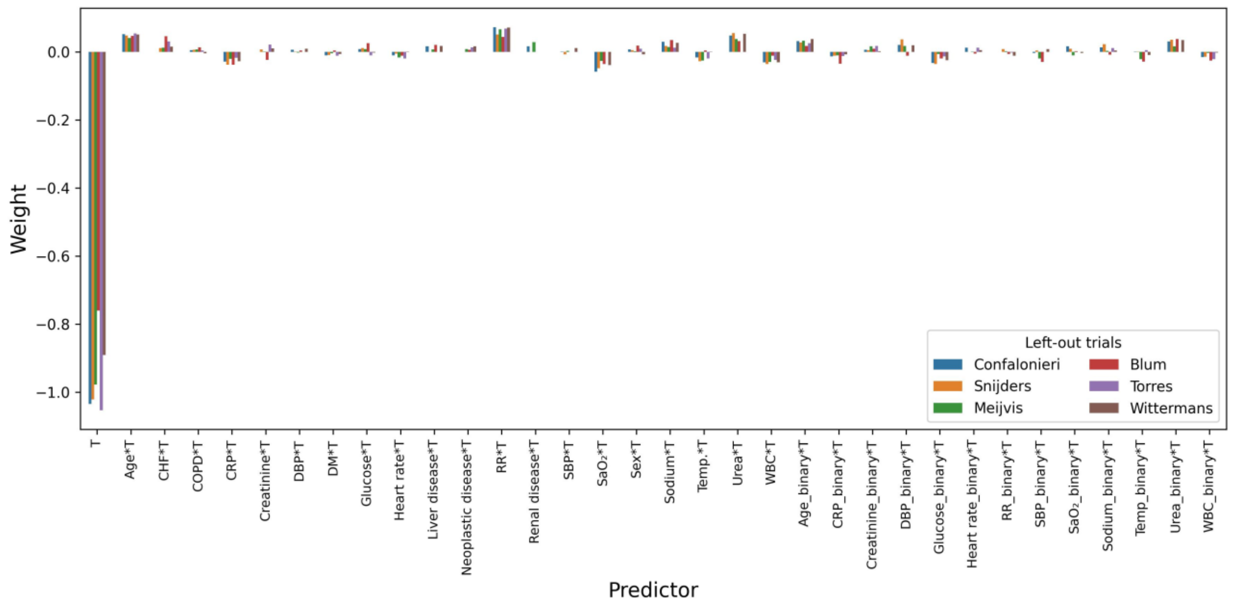

(w) Effect-8, unpenalized

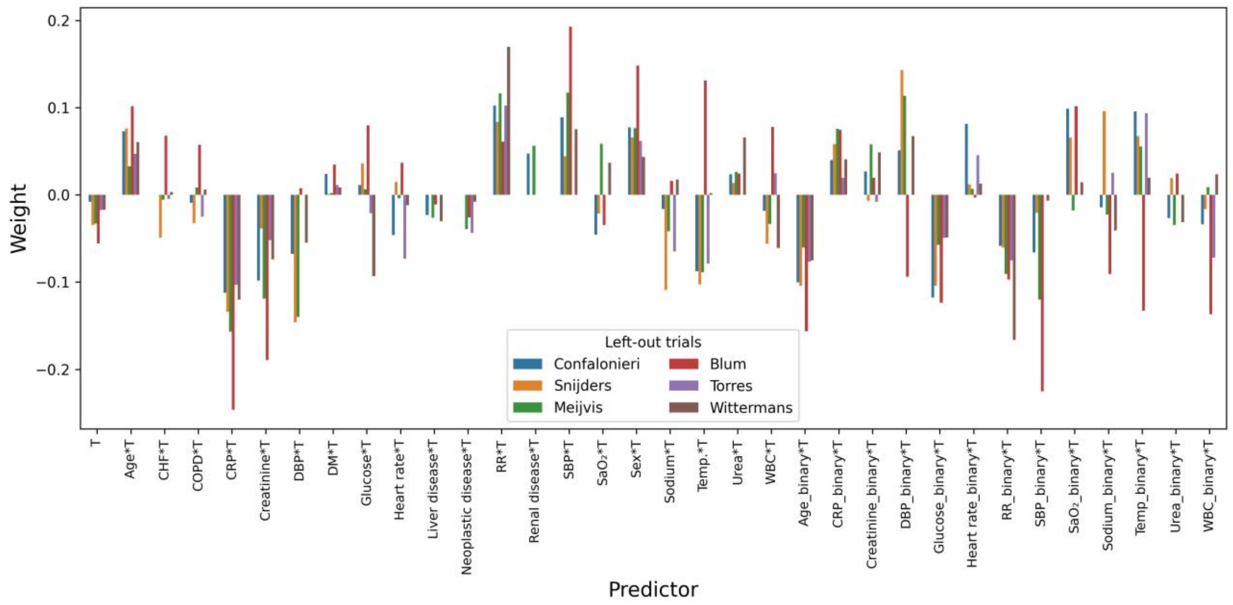

(x) Effect-8, Lasso (‘procedure 2’ in the main text)

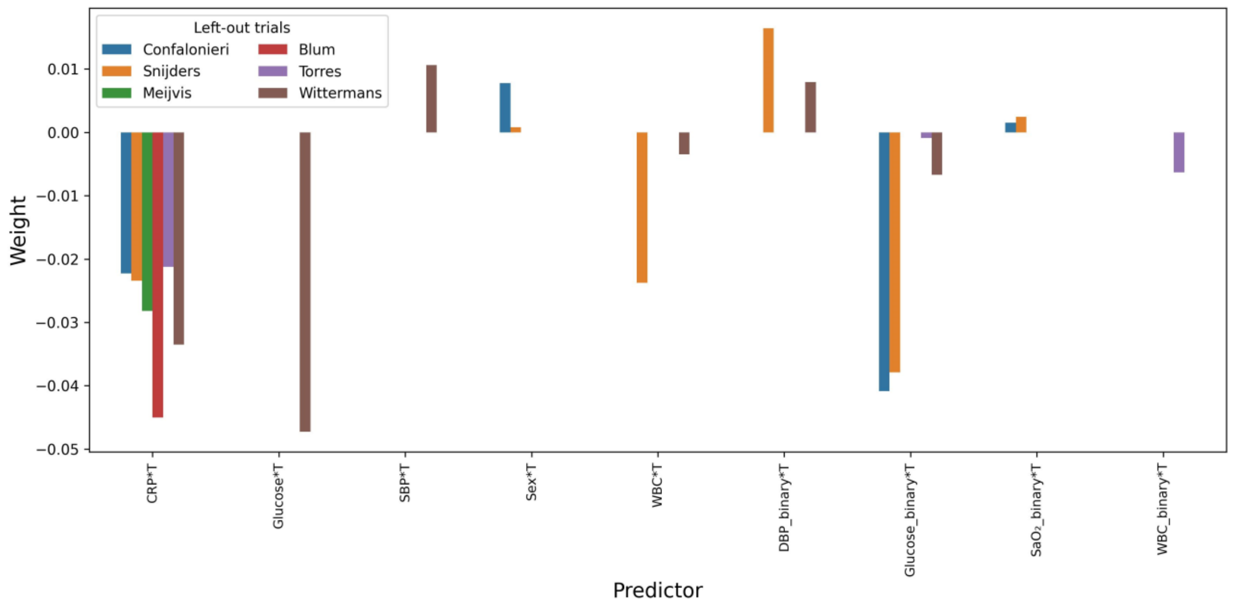

(y) Effect-8, Ridge

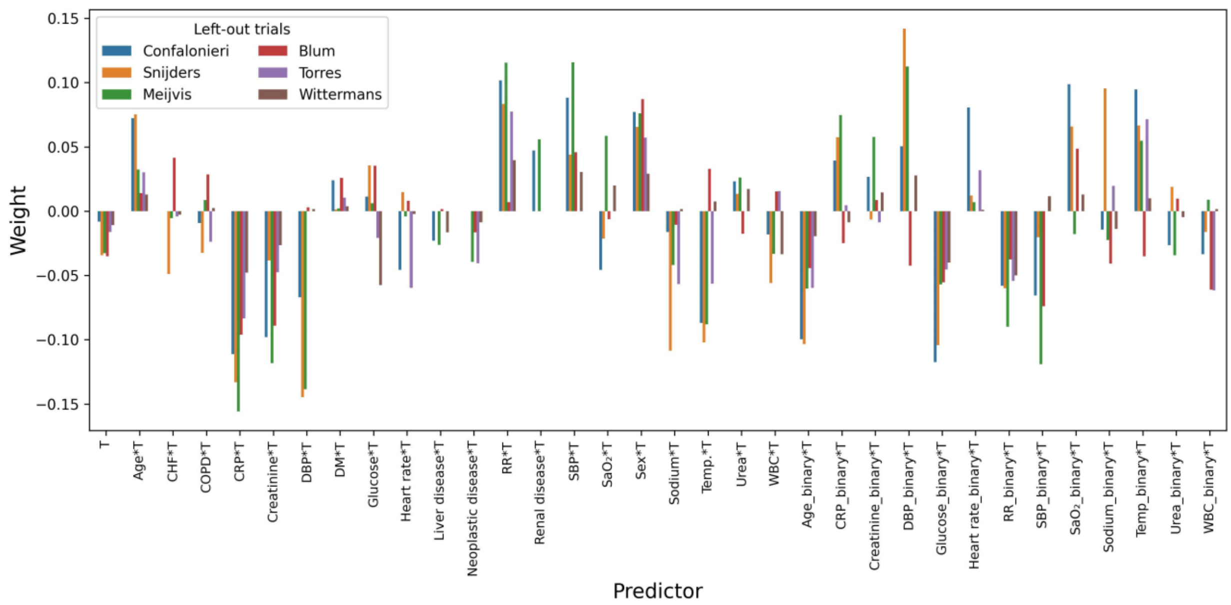

Figure D5: Discriminative performances of the different variations of modelling procedures without additional dichotomized variables, in terms of AUC-benefits (boxplots were created with 2000 bootstrap samples). The Effect-8, LASSO procedure in this figure is described as ‘procedure 1’ in the main text.

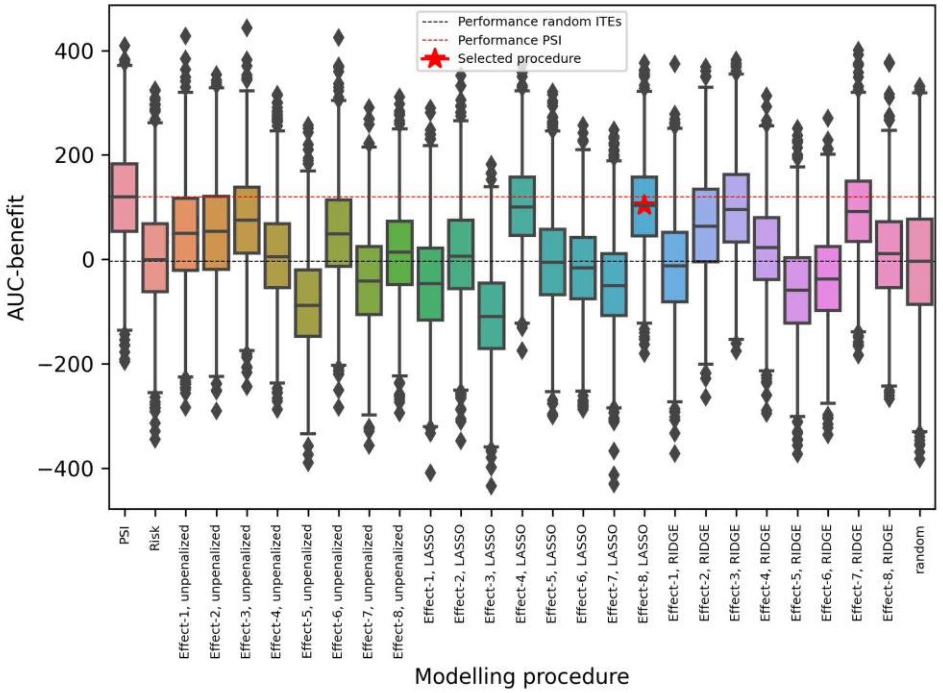

Figure D6: Discriminative performance of the different variations of modelling procedures with additional dichotomized variables, in terms of AUC-benefits (boxplots were created with 2000 bootstrap samples). The Effect-8, LASSO procedure in this figure is described as ‘procedure 2’ in the main text.

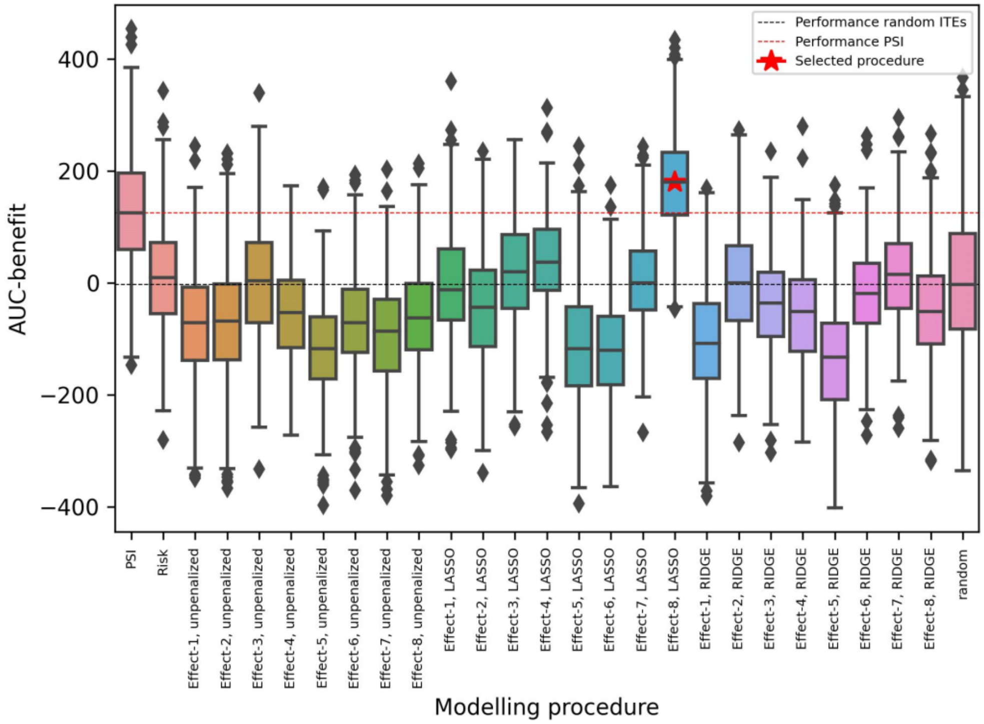

### Appendix E: Models for prospective validation

Training of final models:

To test the performance of the proposed modelling procedures in RCT datasets which did not influence the selection of the modelling procedure (ie, prospective validation), we trained ‘final’ models using all currently available data (ie, the six trials[7,9–13]), and which we will validate using data of the trials by Meduri et al.[5] and Dequin et al.[2], whose datasets were not available to the authors when the modelling procedure was selected and the final models were trained.

To train the final models, we followed modelling procedure 1 and 2 until step 6 (Figure F1), in which data of all six trials formed the train cohort. The λs were optimized in the inner LOTO-CV, in which, for each fold, five of the six trials formed the inner train cohort and the sixth trial the inner test cohort (Figure F2). The results of the wide and fine grid search for λ optimization for procedure 1 and 2 can be found in Figure E1 and E3, respectively. The weights of the trained models resulting from modelling procedure 1 (ie, ‘final model 1’) and 2 (ie, ‘final model 2’) are visualized in Figure E2 and E4, and the exact values of the weights are given in Table E1 and 2, respectively. The weights of final model 3, ie, the extra final model, fitted with the optimal λ which ensured a model with maximally two non-zero weights (see section 5.1 in the main text and Figure E3), are depicted in Figure E5 and Table E3.

The saved Python objects for the trained final models, imputers and scalers (ie, based on the mean and std values for all variables) are available on Github.[14]

Derivation of CRP thresholds:

Assuming a decision threshold (ie, an ITE value above which treating patients is considered worthwhile) of 0, final models 1 and 3 simplify to one and two CRP threshold(s) in the absolute scale (ie, in terms of mg/L), respectively. Final model 1 consists of only one non-zero weight, where the ITE equals 0 for one CRP value. Final model 3 consists of two non-zero weights, ie, CRP and dichotomized glucose, where the ITE equals 0 for two CRP values: one for patients with low (i.e., ≤ 7 mmol/L) and high (i.e., > 7 mmol/L) glucose levels.

Derivation of CRP threshold following from final model 1:

Final model 1 is represented below:

$$Logit\left[P(Y_i = 1|T = t_i, C = c_i)\right] = w_c c_i \underbrace{t_i}_{-1,1}$$

where i indexes the patients, T represents the treatment variable, C the (standardized) CRP value, and $w_c$ the model's weight for the interaction term with CRP (as presented in Table E1).

To find the CRP value that corresponds with an ITE of 0, we equate the models under placebo treatment (ie, $t_i = -1$) and under corticosteroid treatment (ie, $t_i = 1$):

$$Logit\left[P(Y_i = 1|T = -1, C = c_i)\right] = Logit\left[P(Y_i = 1|T = 1, C = c_i)\right]$$

which yields:

$$-w_c c_i = w_c c_i$$
$$c_i = 0$$

Hence, for final model 1, an ITE of 0 corresponds with a standardized CRP value of 0 (ie, the mean), which is 204.1 mg/L.

Derivation of CRP threshold following from final model 3:

Final model 3 is represented below:

$$Logit\left[P(Y_i = 1|T = t_i, C = c_i, G = g_i)\right] = w_c c_i \underbrace{t_i}_{-1,1} + w_g g_i \underbrace{t_i}_{-1,1}$$

Where i indexes the patients, T represents the treatment variable, C the (standardized) CRP value, G the (standardized) dichotomized glucose value, and $w_c$ and $w_g$ the model's weights for the interaction terms with CRP and dichotomized glucose, respectively (as presented in Table E3).

To find the CRP values that corresponds with an ITE of 0, we equate the models under placebo treatment (ie, $t_i$ = -1) and under corticosteroid treatment (ie, $t_i$ = 1):

$$Logit\left[P(Y_i = 1|T = -1, C = c_i, G = g_i)\right] = Logit\left[P(Y_i = 1|T = 1, C = c_i, G = g_i)\right]$$

which yields:

$$-w_c c_i - w_g g_i = w_c c_i + w_g g_i$$

$$-2w_g g_i = 2w_c c_i$$

$$c_i = -\frac{w_g}{w_c} g_i$$

Hence, for each value of $g_i$ (ie, the standardized value for the dichotomized glucose variable), the value for $c_i$ can be calculated using the values for $w_c$ and $w_g$ (as given in Table E3).

For patients with low (i.e., ≤ 7 mmol/L) glucose, $g_i$ equals -0.981446, which yields:

$$c_i = -\frac{-0.032551}{-0.030991} * -0.981446$$

$$c_i = 1.03085$$

Given a mean value of 204.1 mg/L and a standard deviation of 132.98 mg/L for CRP, this corresponds to a CRP threshold of **341.2 mg/L**.

For patients with high (i.e., > 7 mmol/L) glucose, $g_i$ is 1.018905, which yields:

$$c_i = -\frac{-0.032551}{-0.030991} * 1.018905$$

$$c_i = -1.0702$$

which, given a mean value of 204.1 mg/L and a standard deviation of 132.98 mg/L for CRP, this corresponds to a CRP threshold of **61.8 mg/L**.

Figure E1: Results of the initial wide and second fine grid search for λ optimization for final model 1 (trained using all six trials).

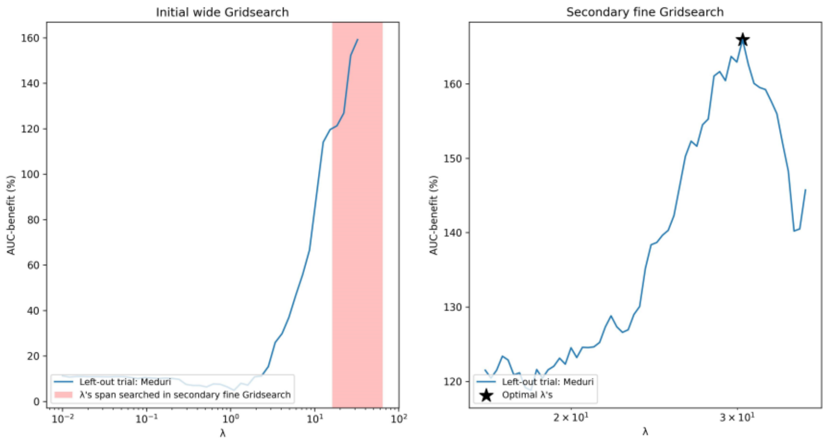

Figure E2: Bar charts of all non-zero weights of final model 1. CRP=C-reactive protein, T=treatment variable.

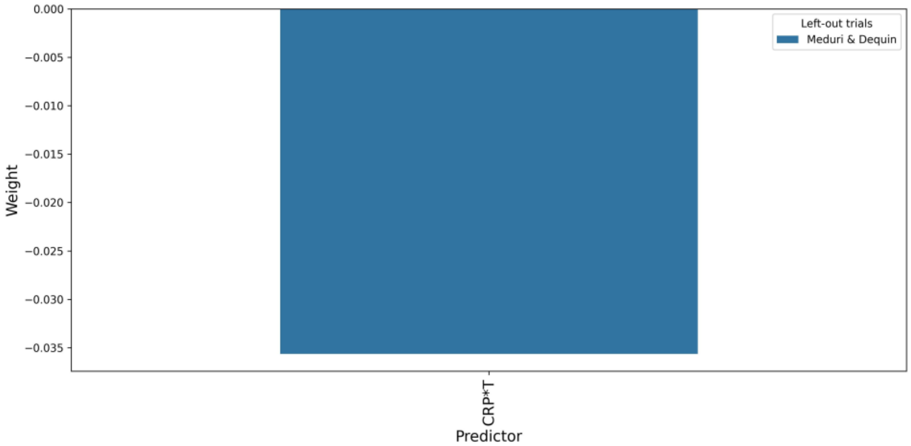

Figure E3: Results of the initial wide and second fine grid search for λ optimization for final models 2 (which used the optimal λ) and 3 (which used the optimal λ among λs that yield maximally two non-zero weights).

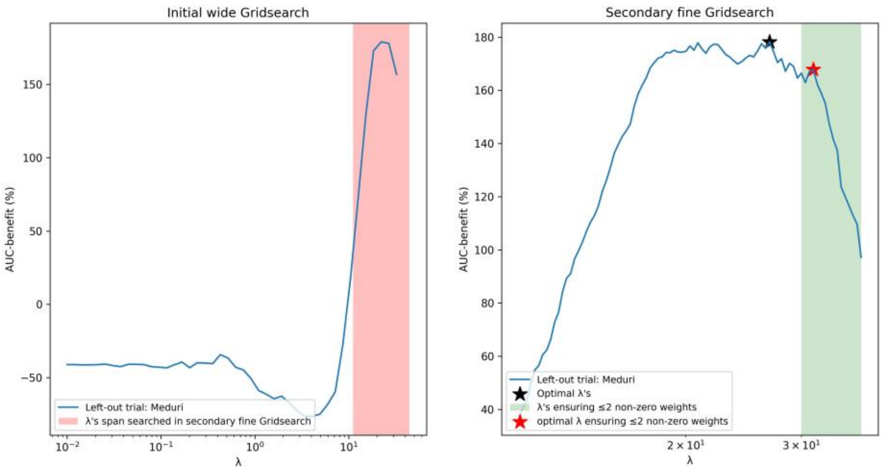

Figure E4: Bar charts of all non-zero weights of final model 2. CRP=C-reactive protein, T=treatment variable.

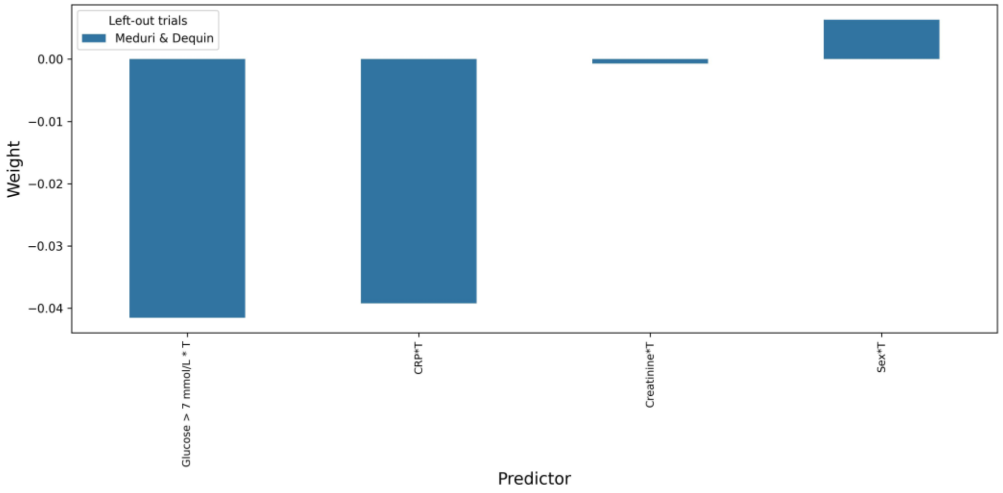

Figure E5: Bar charts of all non-zero weights of final model 3. CRP=C-reactive protein, T=treatment variable.

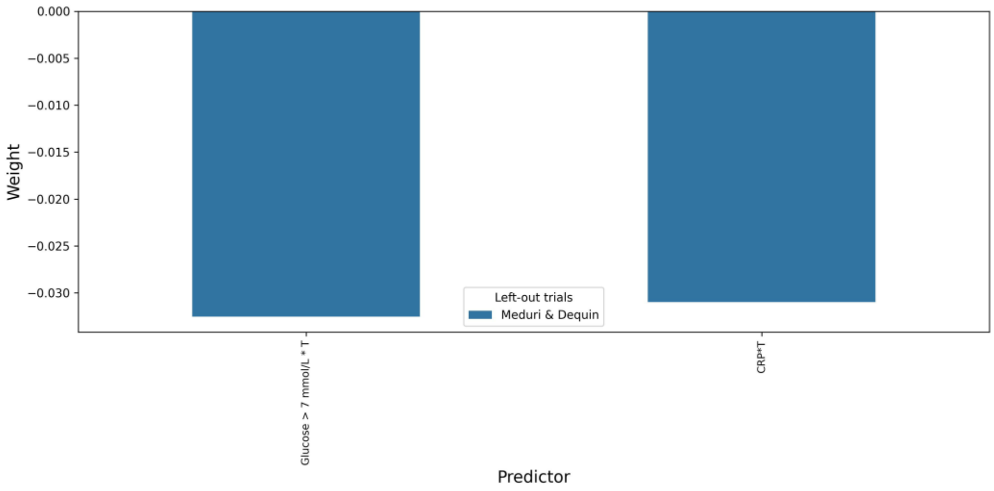

Table E1: Values of all non-zero weights of  final model 1. CRP=C-reactive protein, T=treatment variable.

| *Variable* | weight |
|---|---|
| *CRP*T* | -0,035644731369696 |

Table E2: Values of all non-zero weights of  final model 2. CRP=C-reactive protein, T=treatment variable.

| *Variable* | weight |
|---|---|
| *Sex*T* | 0,0063385987642517 |
| *Creatinine*T* | -0,0007262401947579 |
| *CRP*T* | -0,0392263630352131 |
| *Glucose > 7 mmol/L *T* | -0,041542017923151 |

Table E3: Values of all non-zero weights of  final model 3. CRP=C-reactive protein, T=treatment variable.

| *Variable* | weight |
|---|---|
| *CRP*T* | -0,0309912129137976 |
| *Glucose > 7 mmol/L *T* | -0,0325513665681267 |

**Appendix F: Supplementary figures and tables**

Figure F1: Visualization of the outer loop of the full modelling procedures.
* For procedure 2 (ie, with extra dichotomized variables), in step 2, after imputation of missing values, dichotomized variables for each continuous variable are added.
** Step 5, ie, the optimization of the λs, is separately visualized in Figure F2.

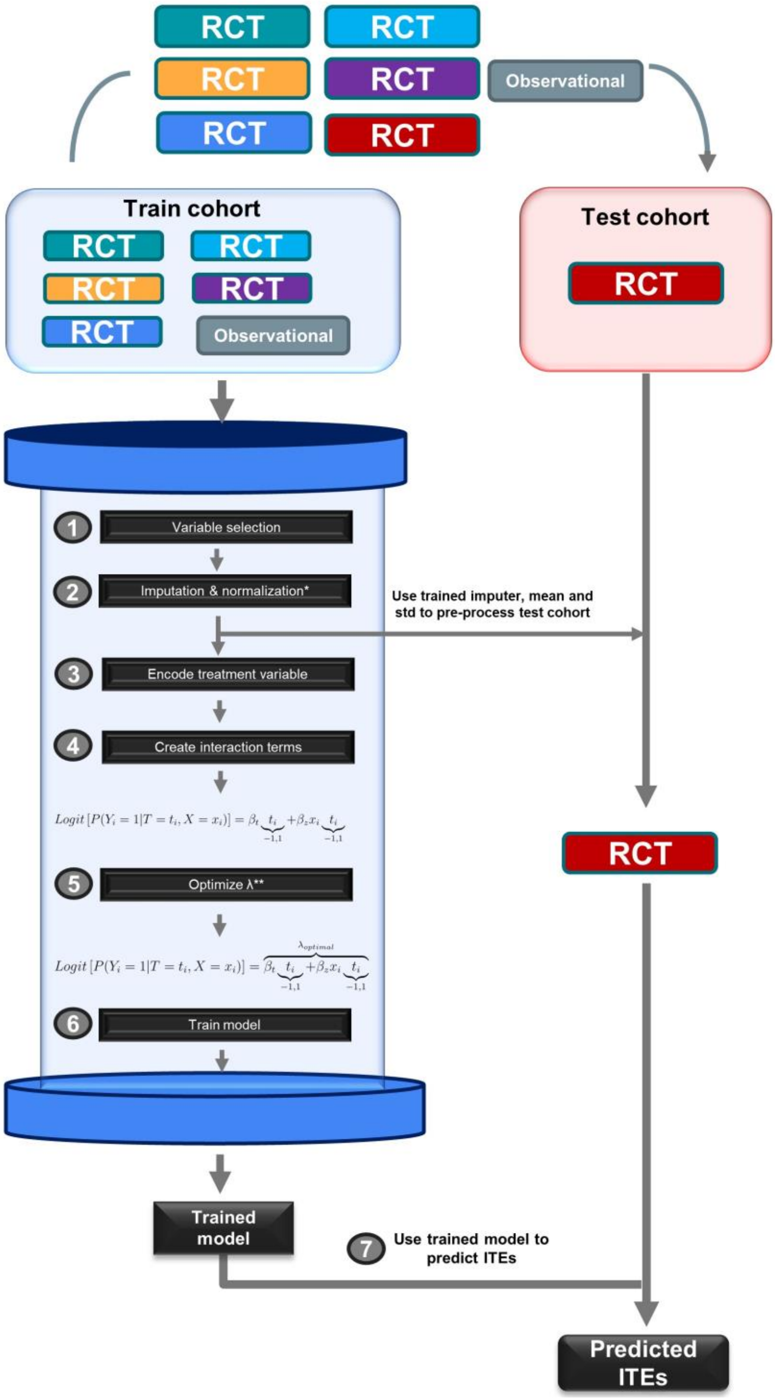

Figure F2: Visualization of the inner loop of the modelling procedures, used to optimize the λs (this represents step 5 of the full modelling procedure, Figure F1).
* For procedure 2 (ie, with extra dichotomized variables), in step 2, after imputation of missing values, dichotomized variables for each continuous variable are added.

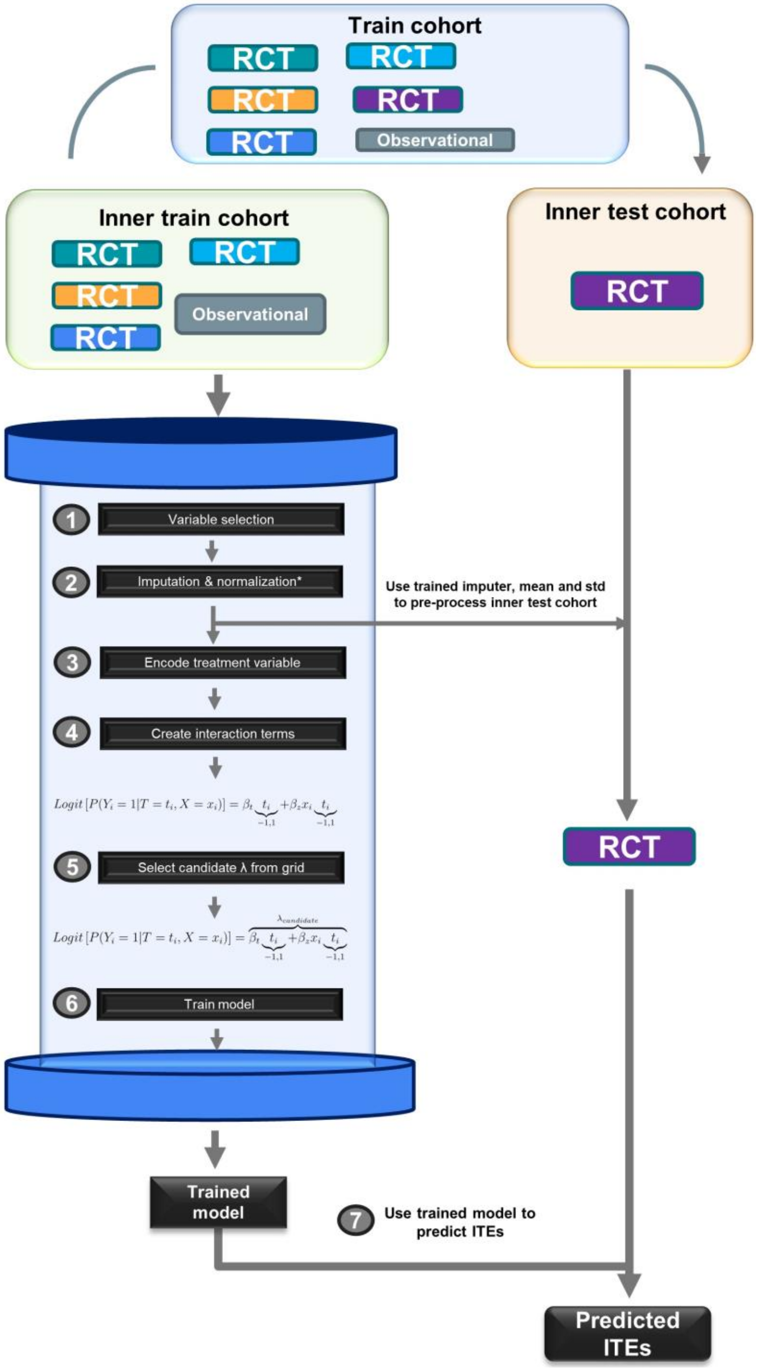

Figure F3: Cumulative dose of corticosteroids for each study. All doses were transformed into equivalent quantities of hydrocortisone (in mg), using Clincalc's Corticosteroid Conversion Calculator.[15] To calculate the cumulative dose in the treatment regime of *Torres et al*[12]., which assigned patients in the treatment arm to 0.5 mg/kg per 12 hours of methylprednisolone, we assumed an average weight of 84 kg for male patients and 65.9 kg for female patients.[16]

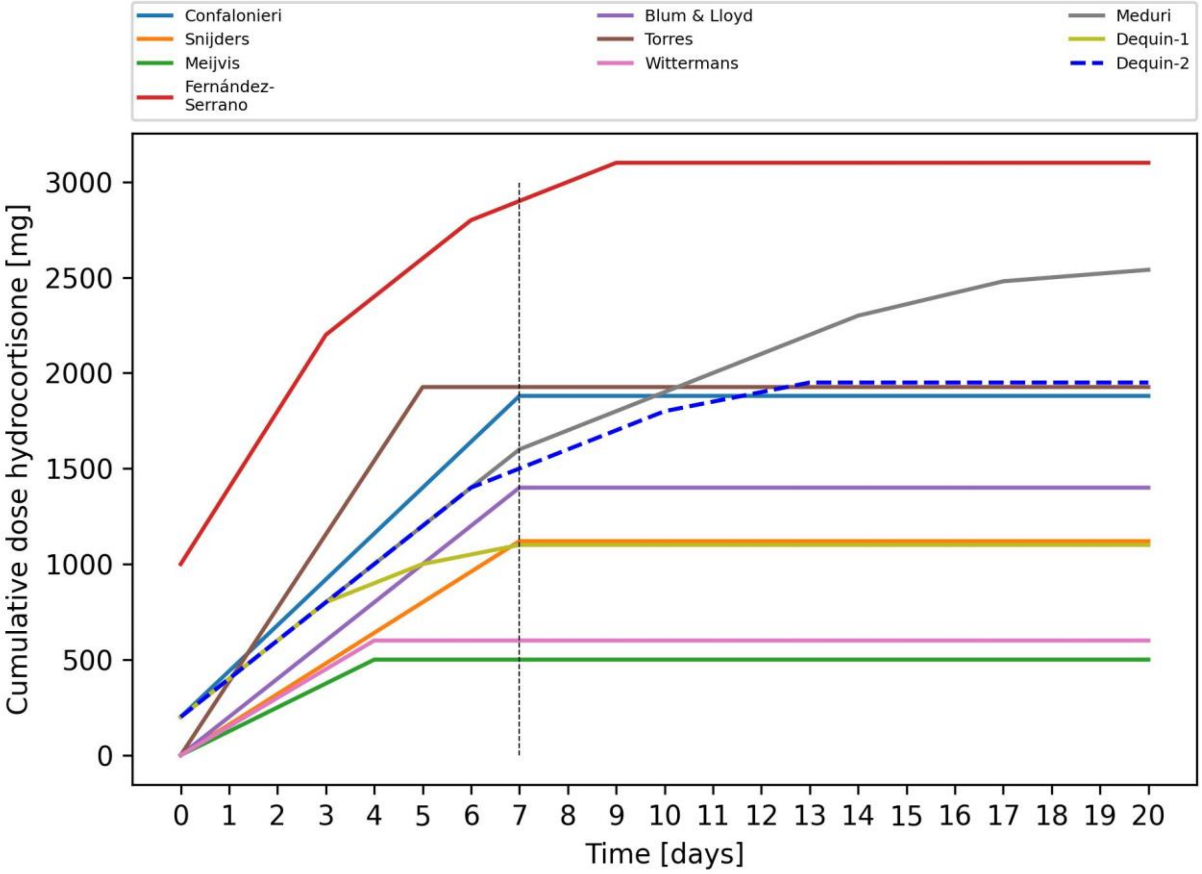

Figure F4: Violin plots representing the distributions of included variables among the patients from the different included trials and the observational study[17] (left panel) and the distributions split for treatment arms for all included trials (right panel). The x-axis specifies the number of patients per distribution (which could be smaller than the study size due to missingness). In some trials, a variable was completely missing and therefore no distribution is plotted. Distributions of the placebo and corticosteroid arms were compared using a Fisher exact test for categorical variables and a two-sample t test for continuous variables. Significant differences between the distributions (ie, P<0.05) are marked with an asterisk (*).

(a) Sex

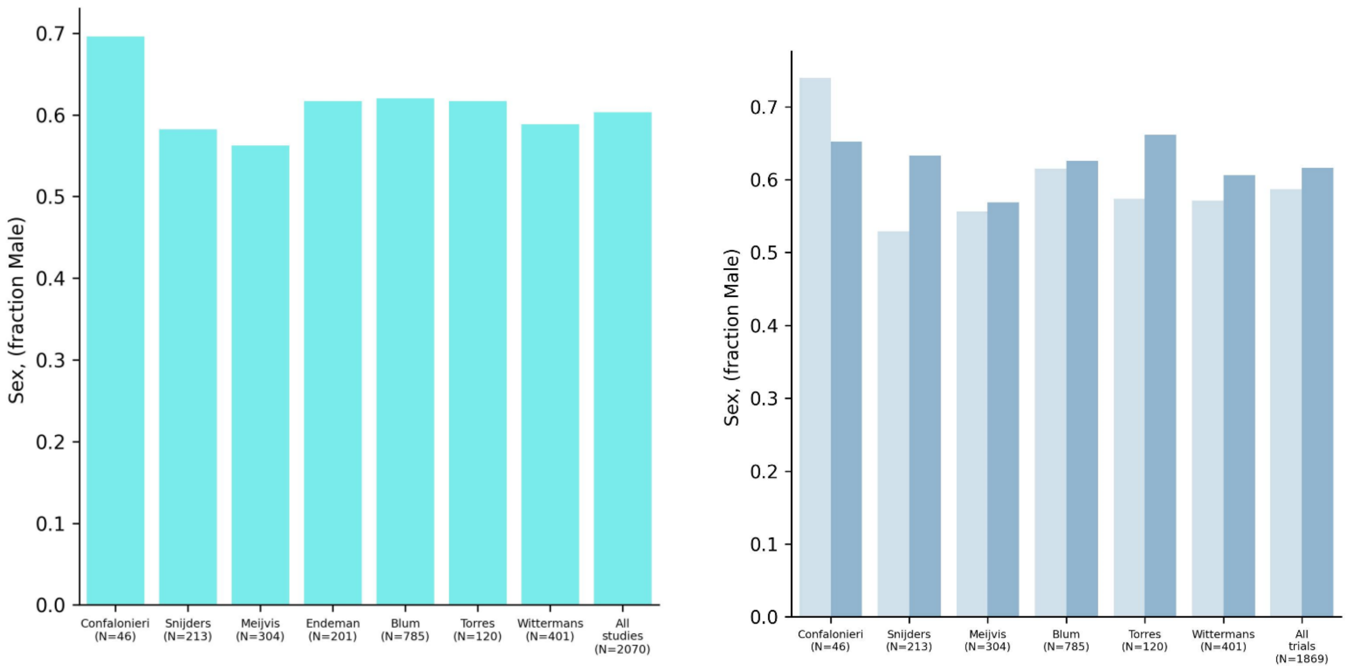

(b) Age

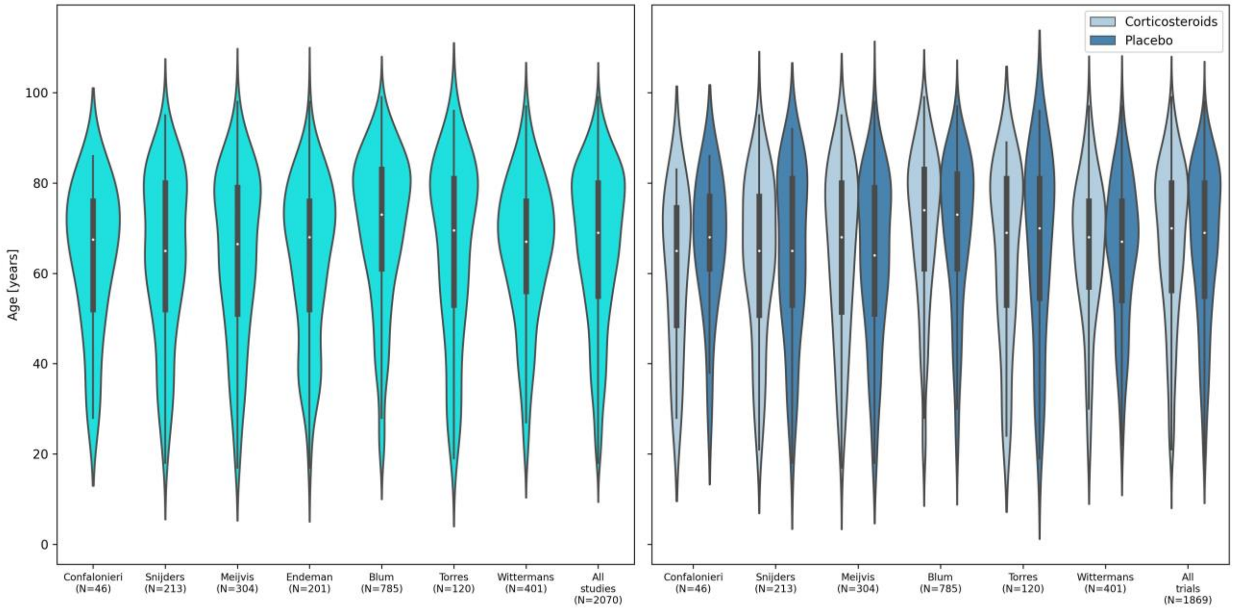

(c) Respiratory rate

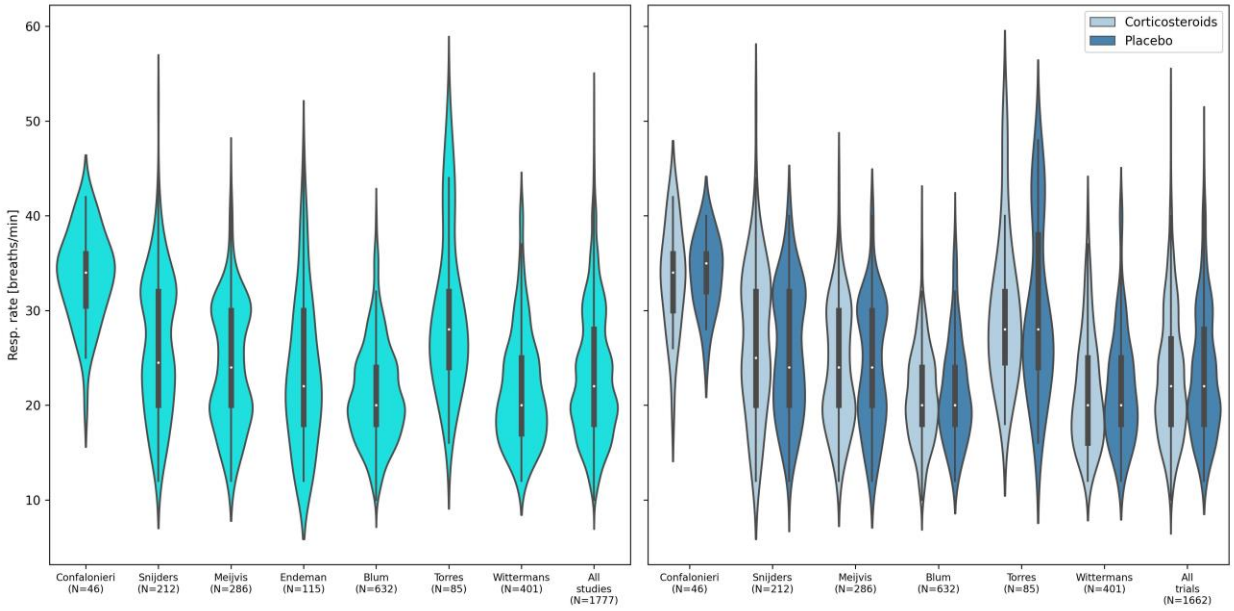

(d) Diastolic blood pressure

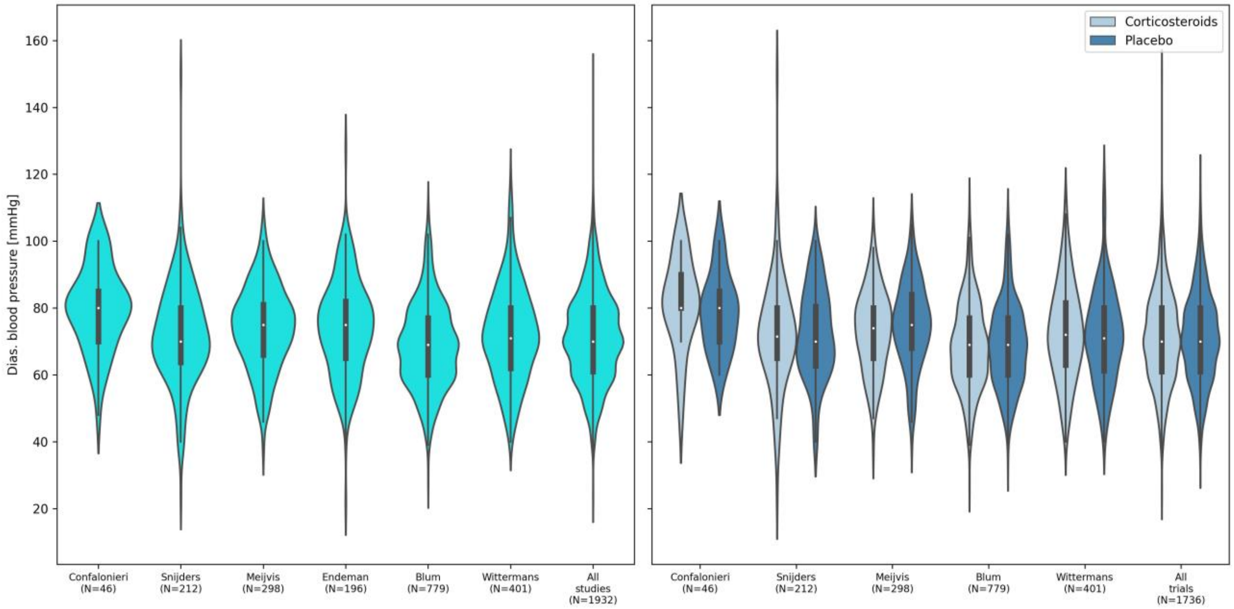

(e) Systolic blood pressure

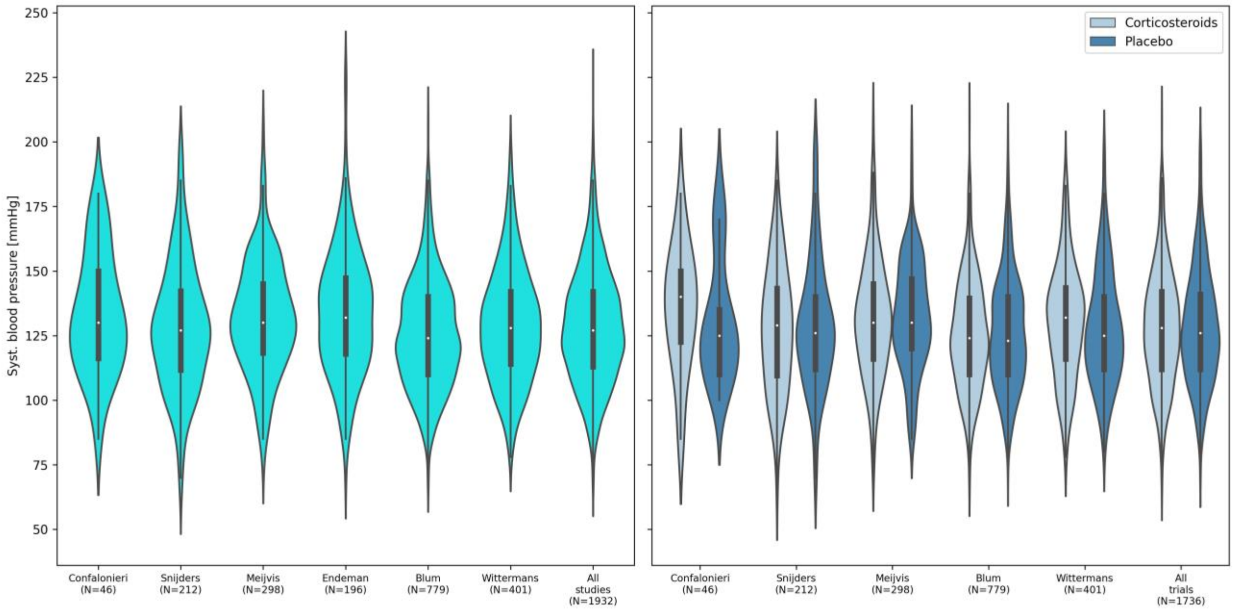

(f) Temperature

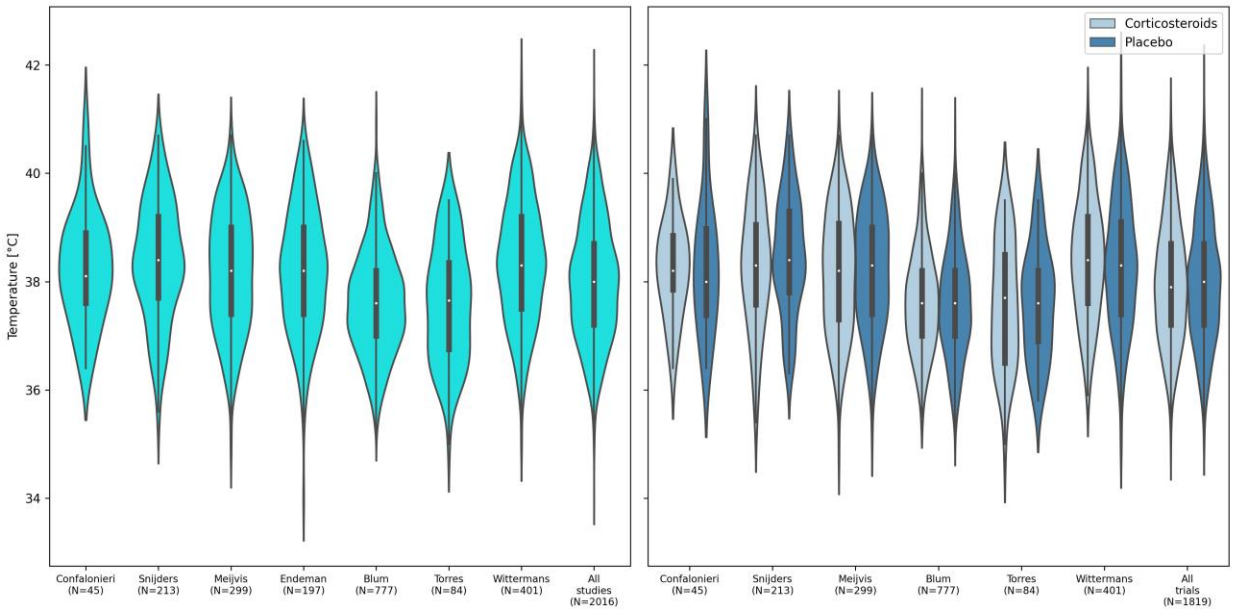

(g) Heart rate

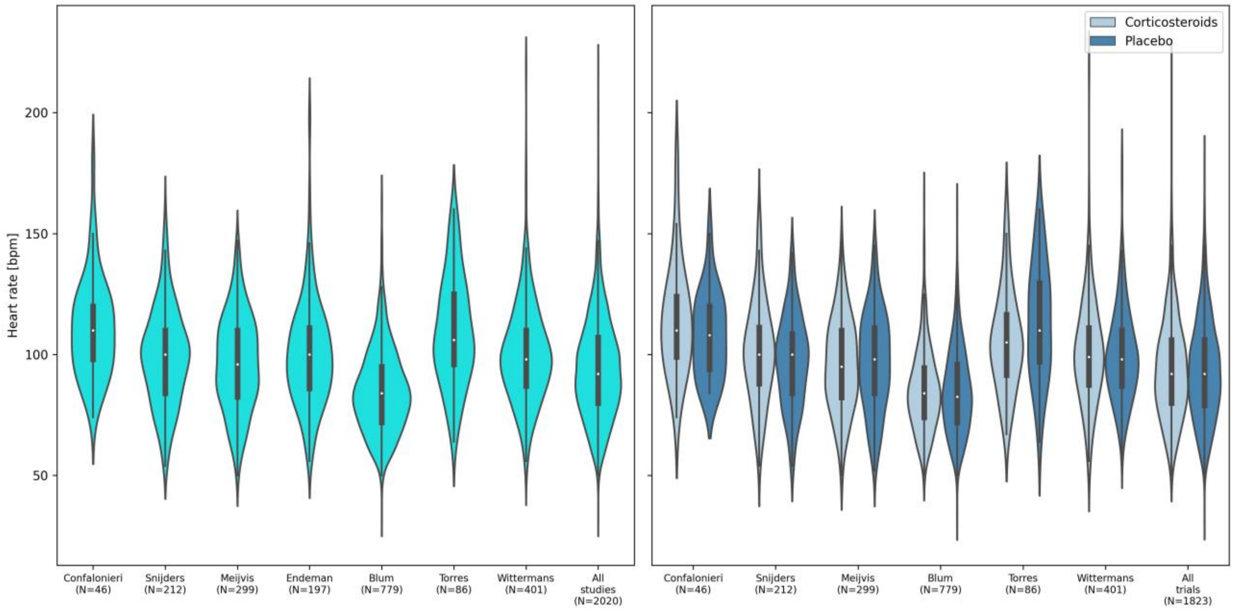

(h) Oxygen saturation ($S_aO_2$)

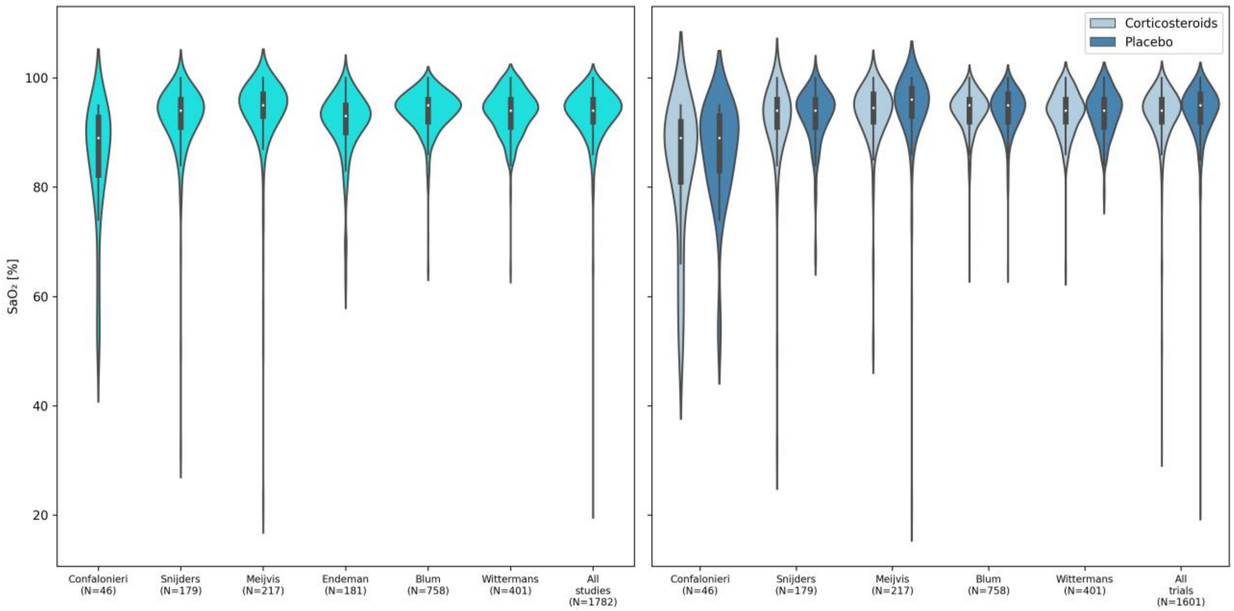

(i) Creatinine

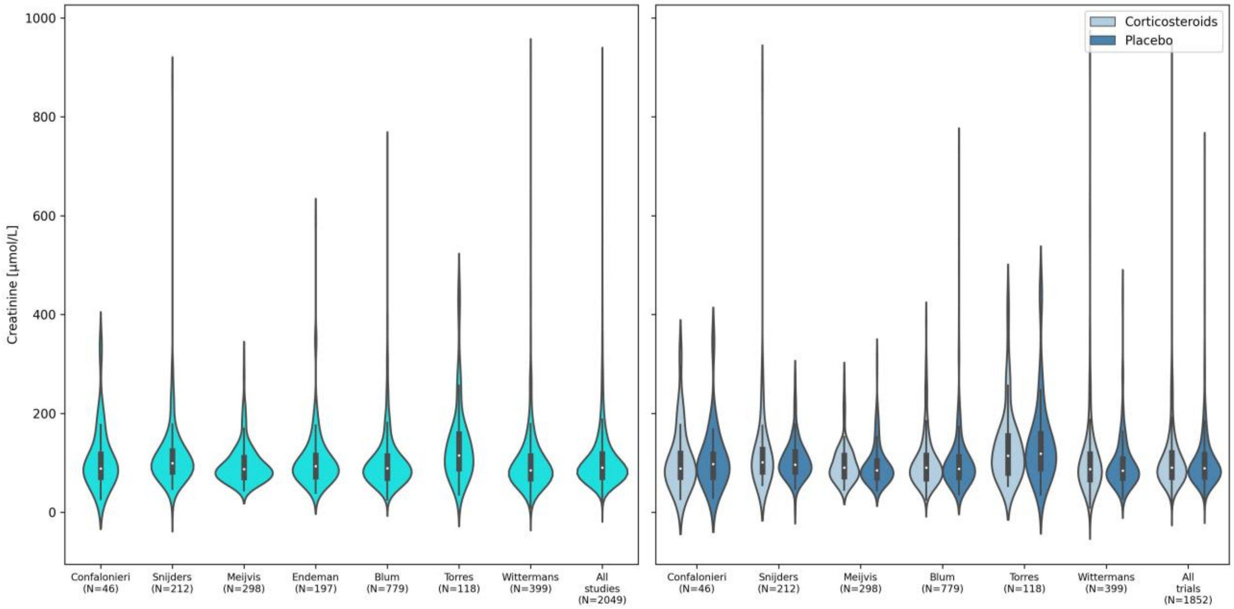

(j) Sodium

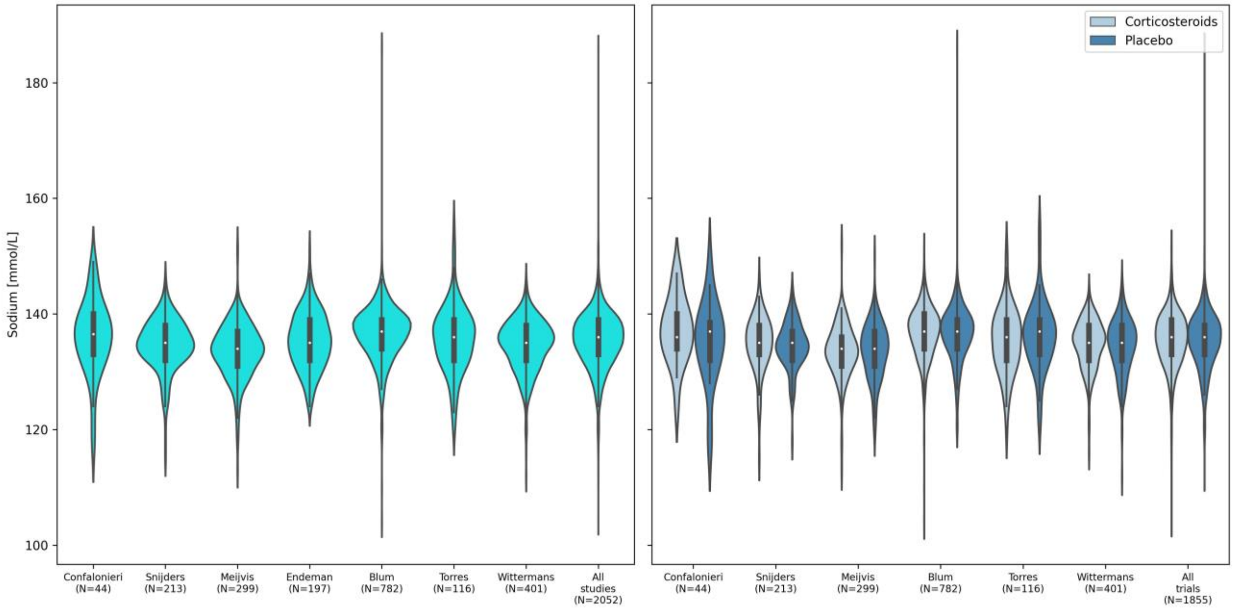

(k) Urea

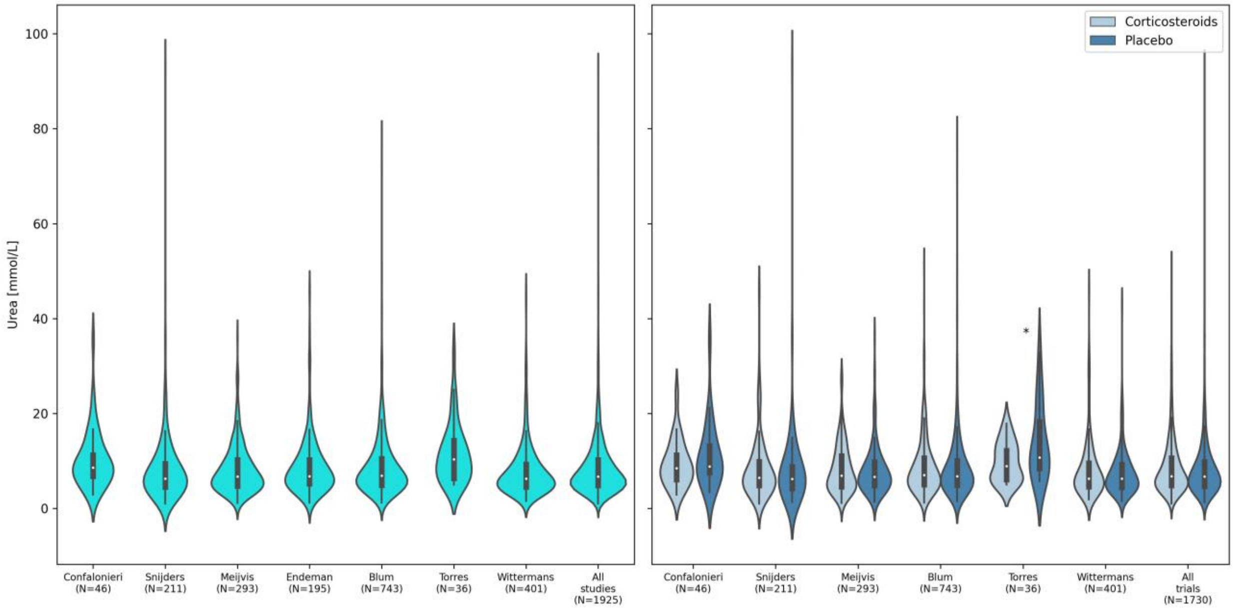

(l) C-reactive protein

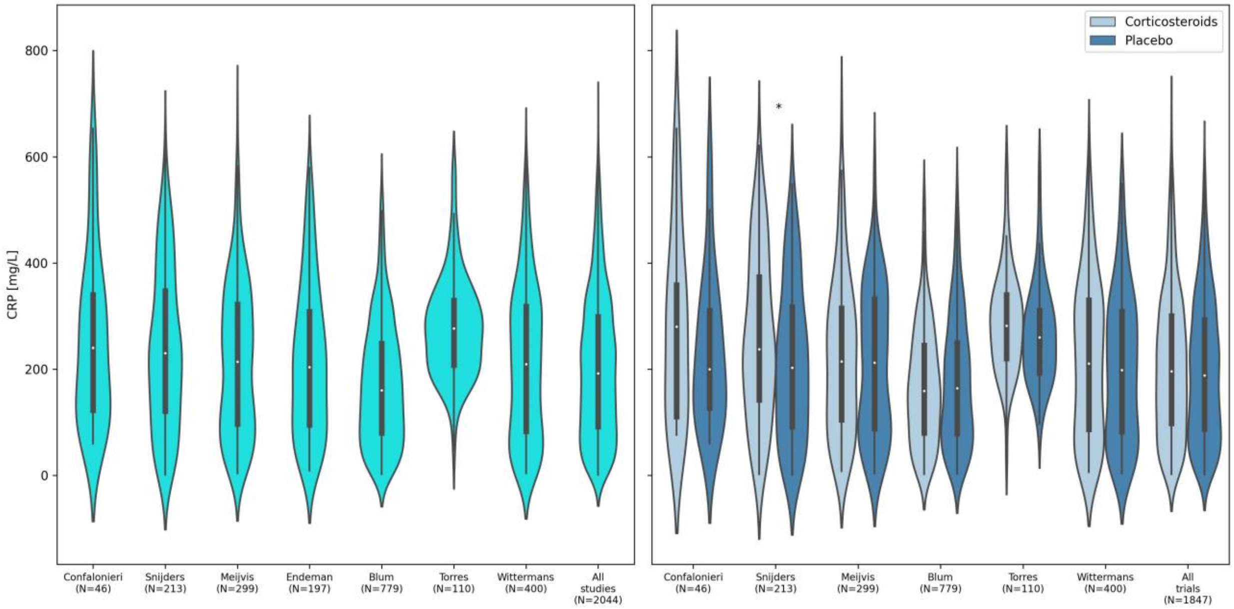

(m) Glucose

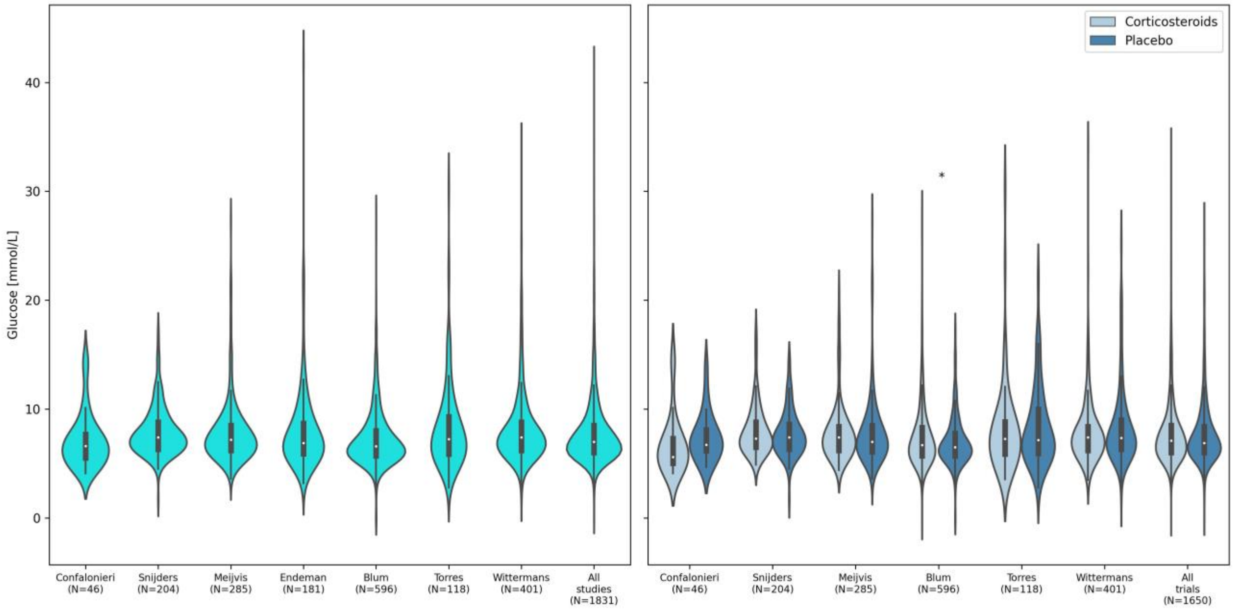

(n) White cell count

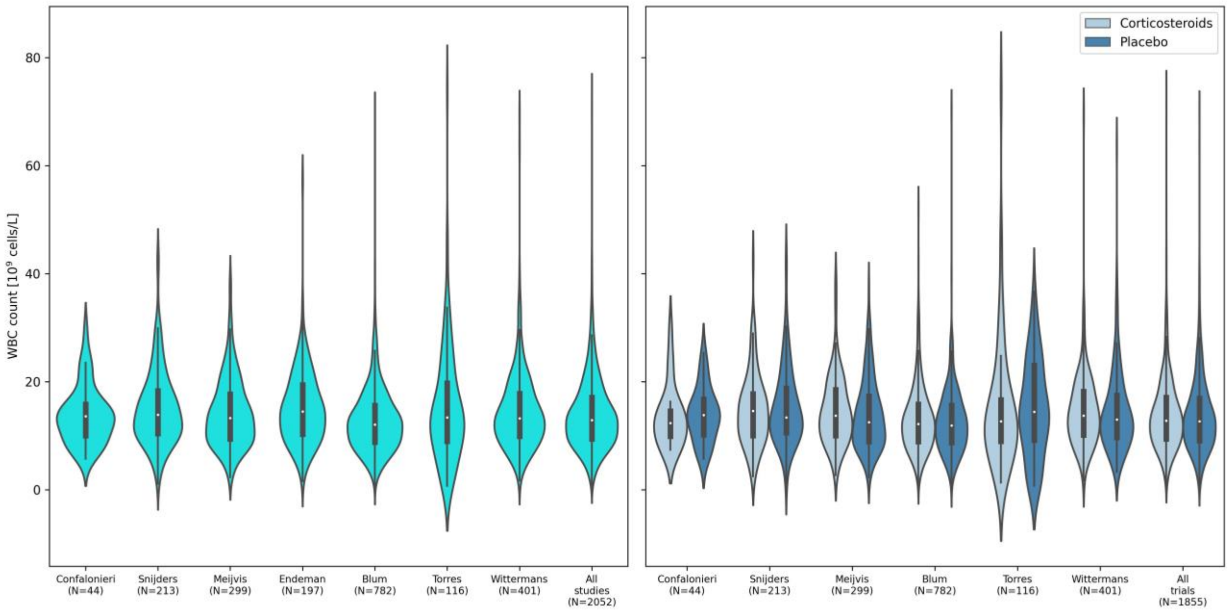

(o) Neoplastic disease

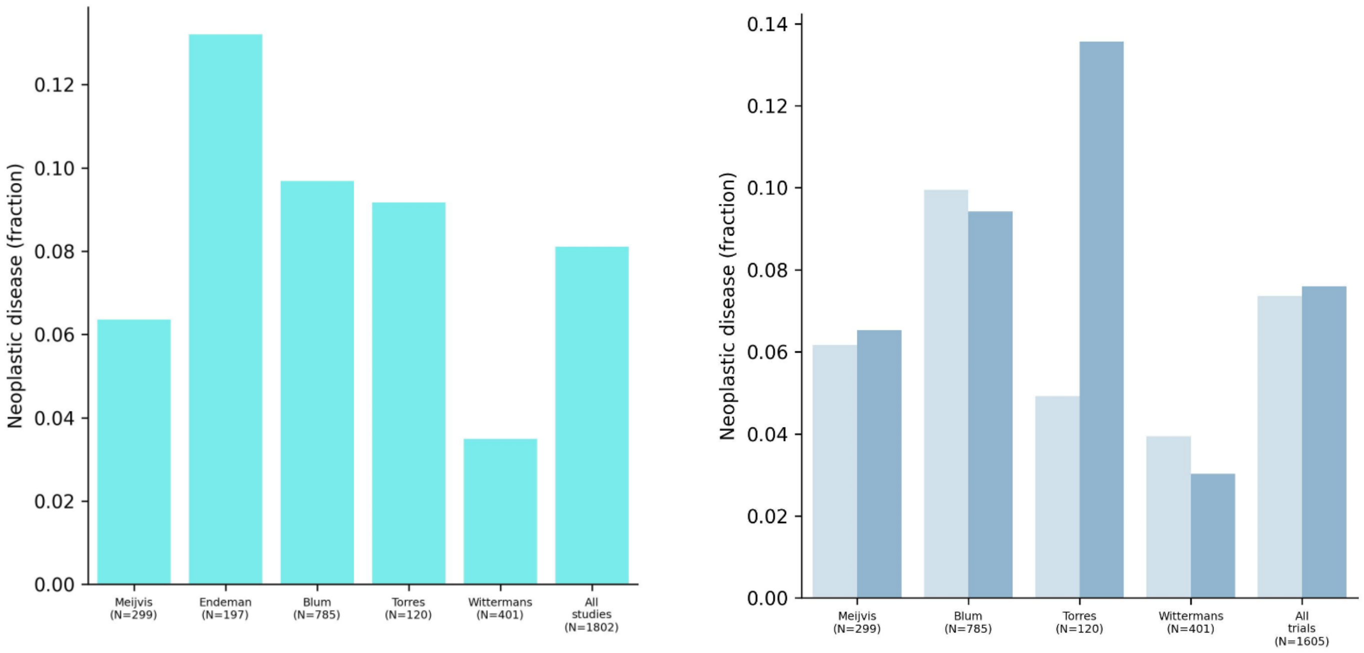

(p) Liver disease

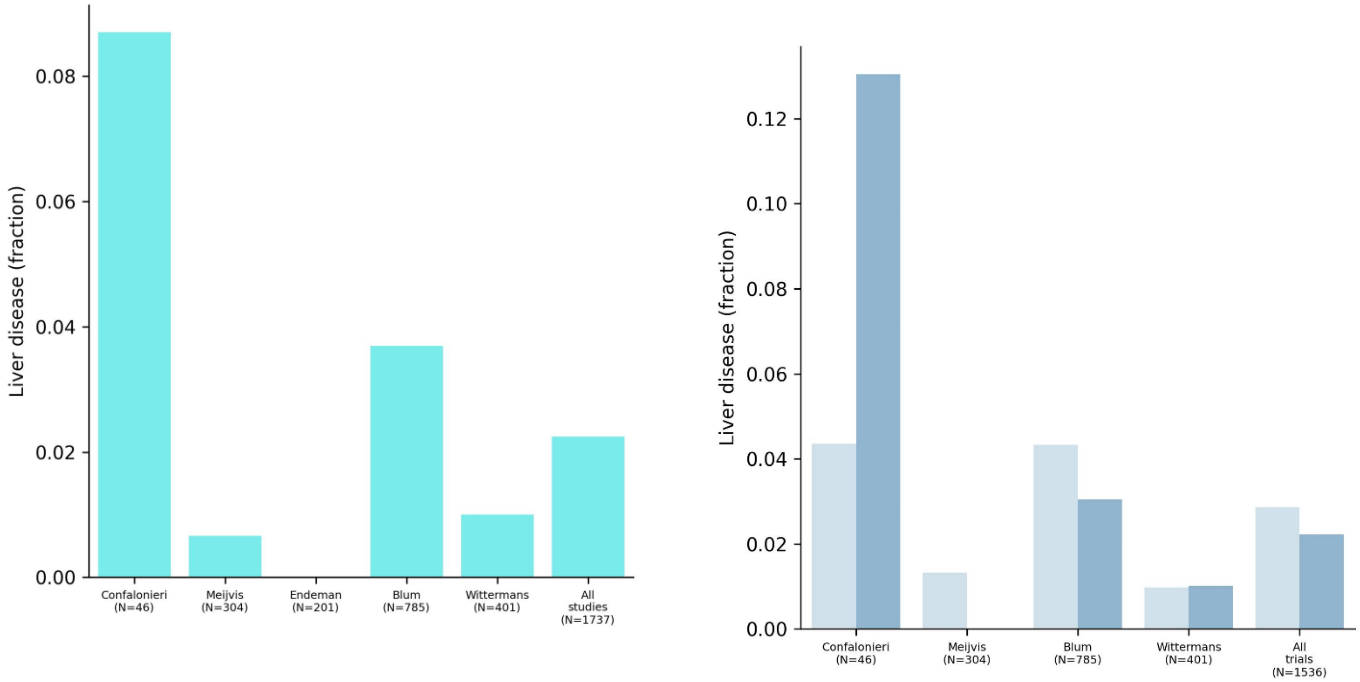

(q) Congestive heart failure

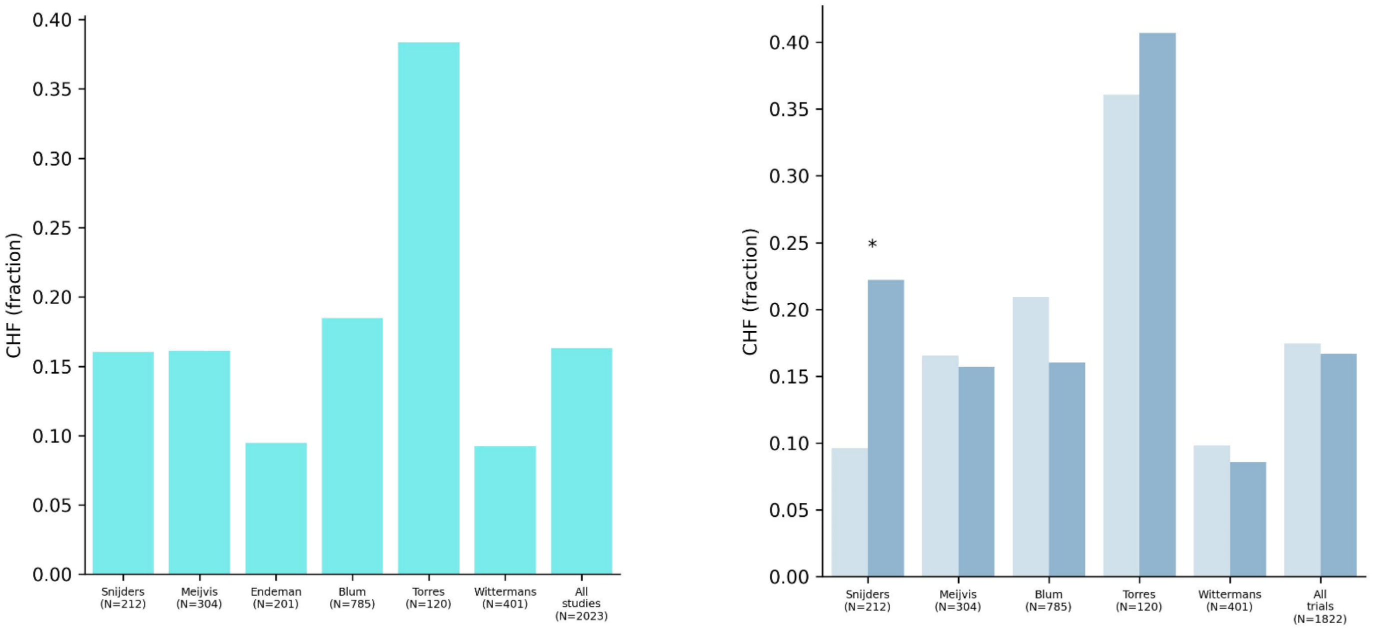

(r) Renal disease

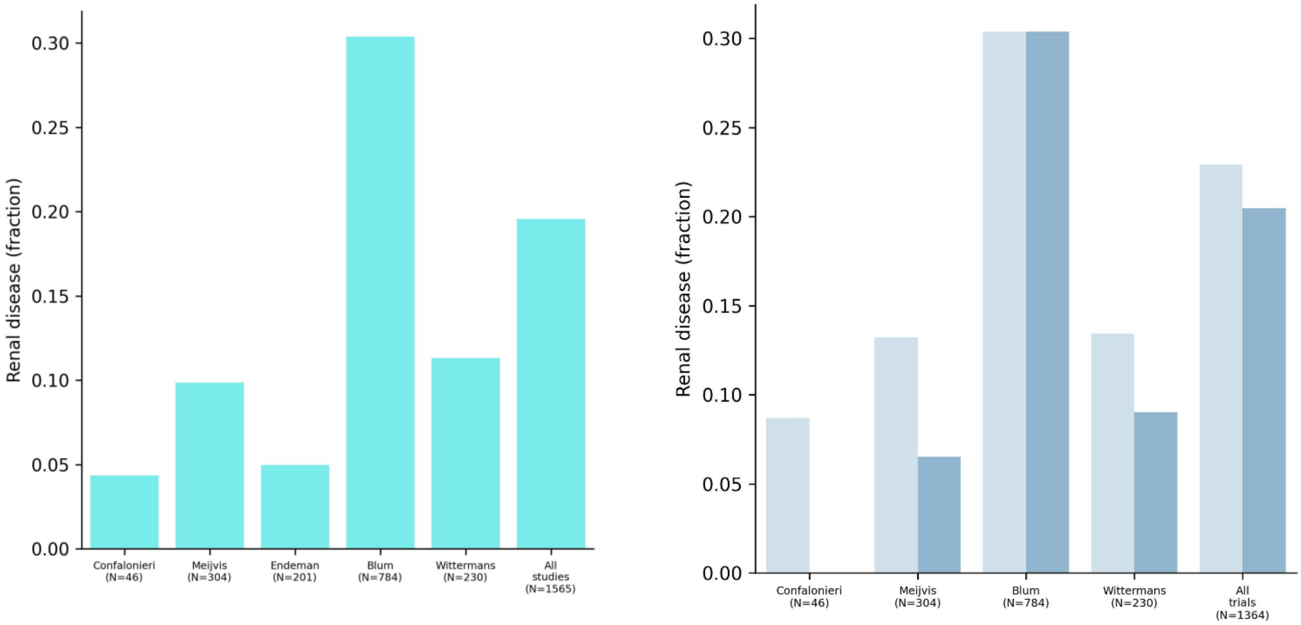

(s) Diabetes Mellitus

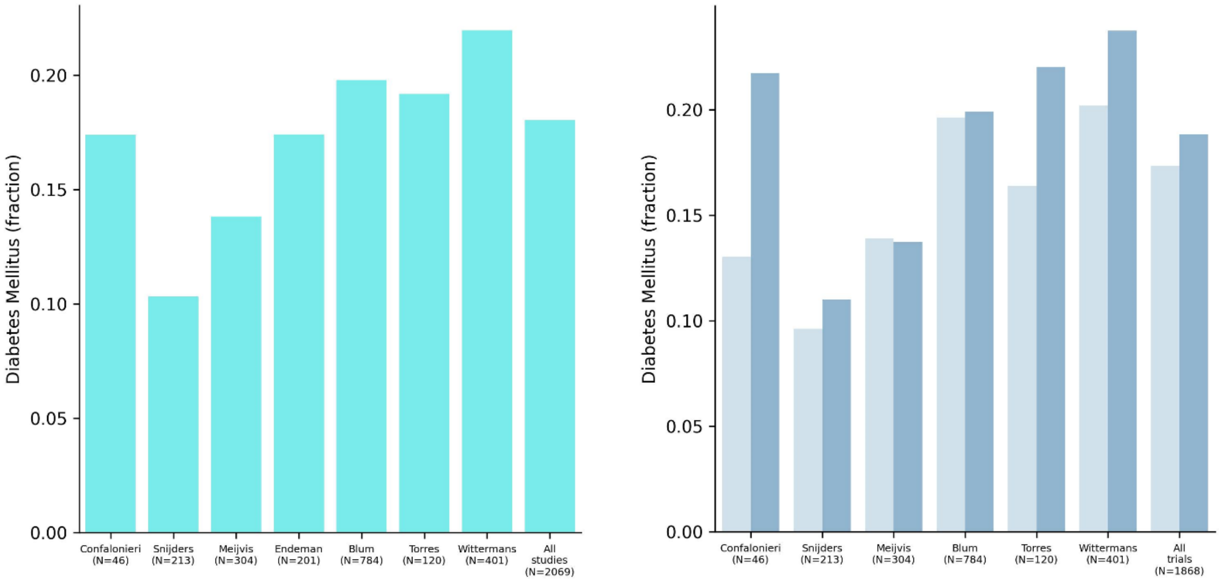

(t) Chronic obstructive pulmonary disease

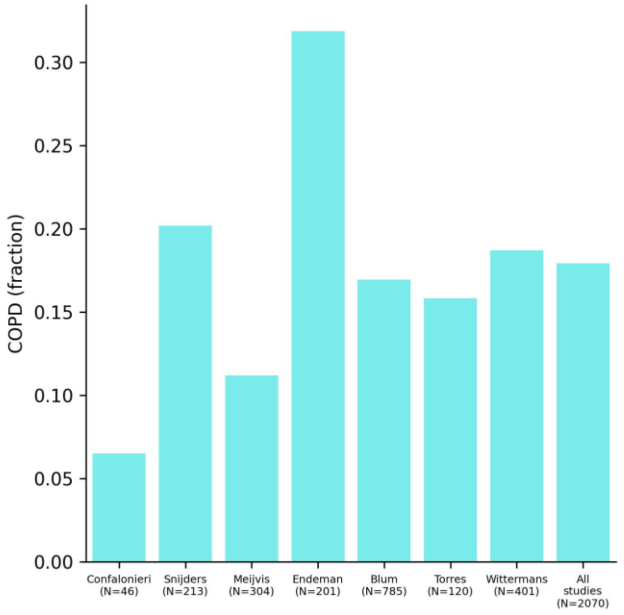

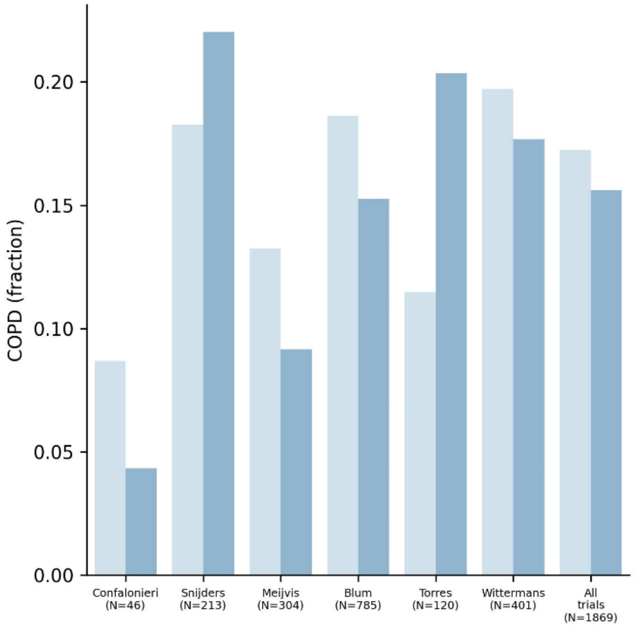

(u) PSI

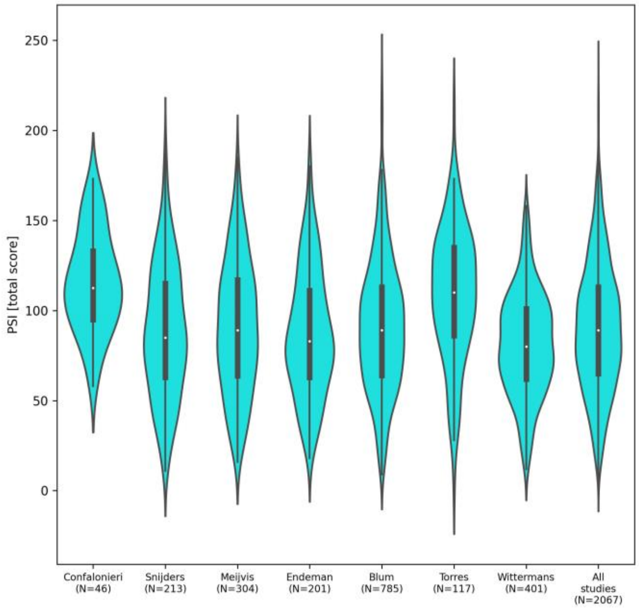

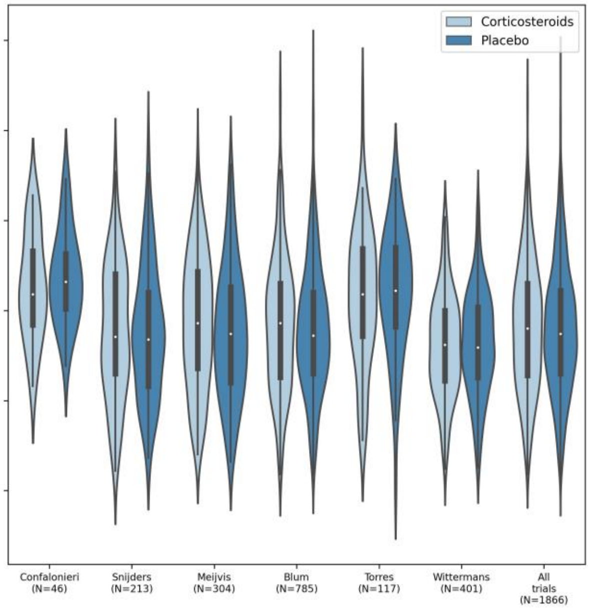

Figure F5: Results for each fold of the leave-one-trial-out cross-validation (LOTO-CV) separately, in procedure 1. Mortality rates (left), odds ratios (middle), and absolute mortality differences (right) are shown with 95% CIs. Number of events and total number of patients per subgroup are shown on the x-axis. In case the odds ratio was not defined (ie, when the mortality rate in the control arm is 0), or the point estimate was zero (ie, when the mortality rate in the treatment arm is 0), this part of the figure is omitted. In case the lower bound of the CI for the odds ratio was zero, this was annotated with an arrow.

(a) LOTO-CV fold in which Confalonieri et al[9] formed the test cohort

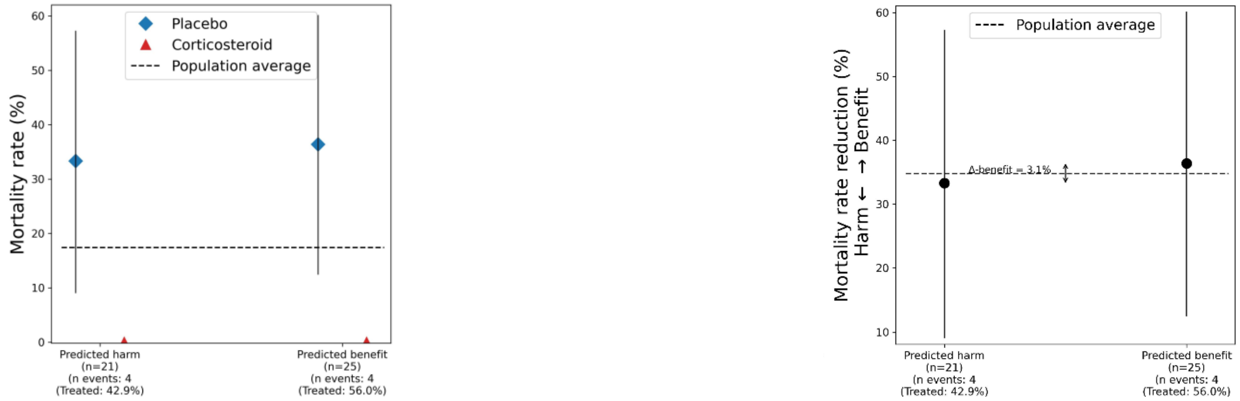

(b) LOTO-CV fold in which Snijders et al[10] formed the test cohort

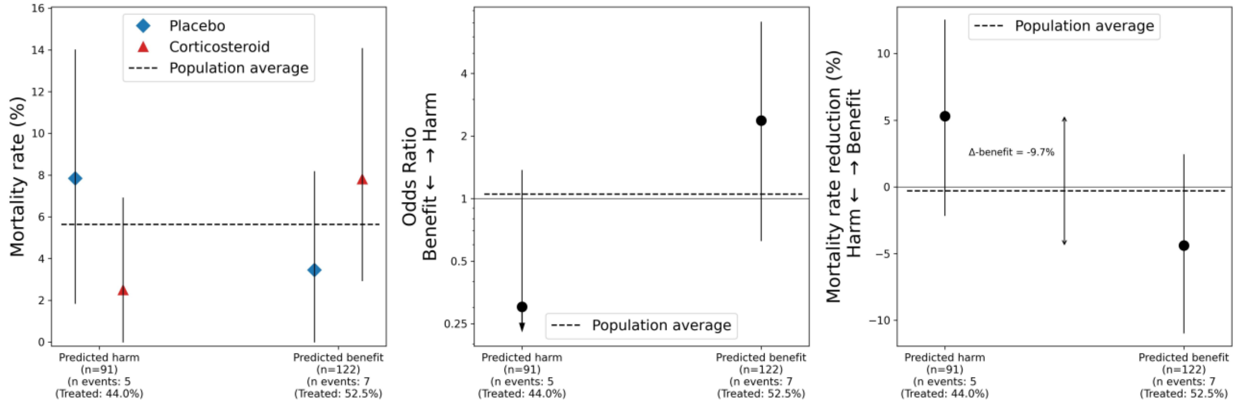

(c) LOTO-CV fold in which Meijvis et al.[11] formed the test cohort

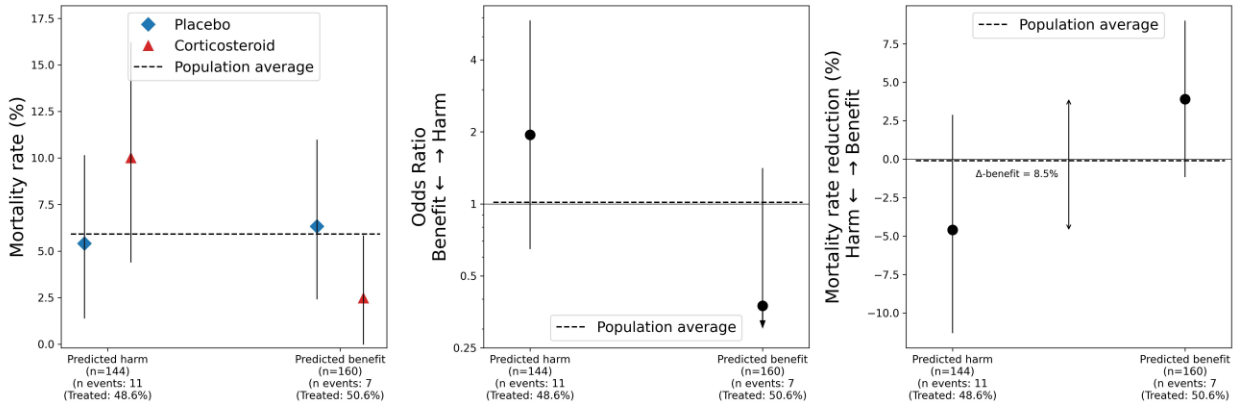

(d) LOTO-CV fold in which Blum et al.[7] formed the test cohort

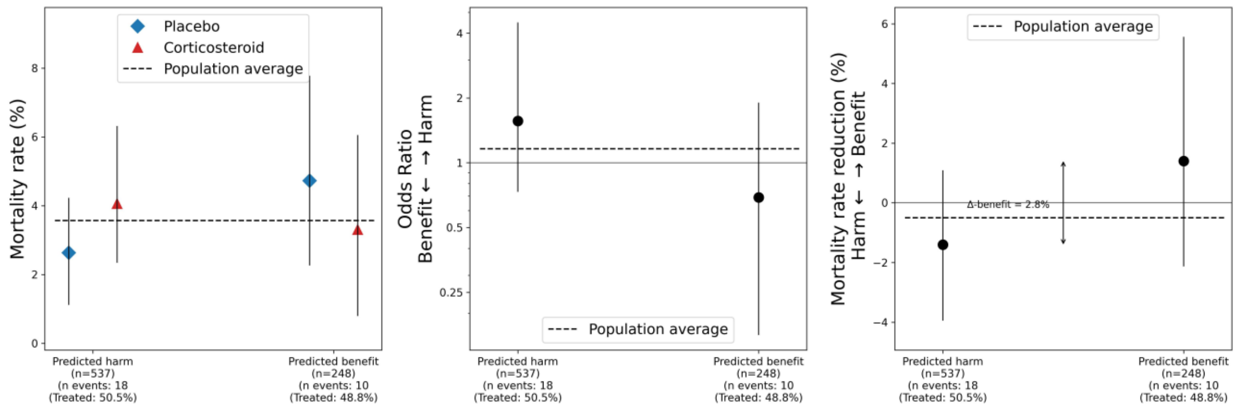

(e) LOTO-CV fold in which Torres et al.[12] formed the test cohort

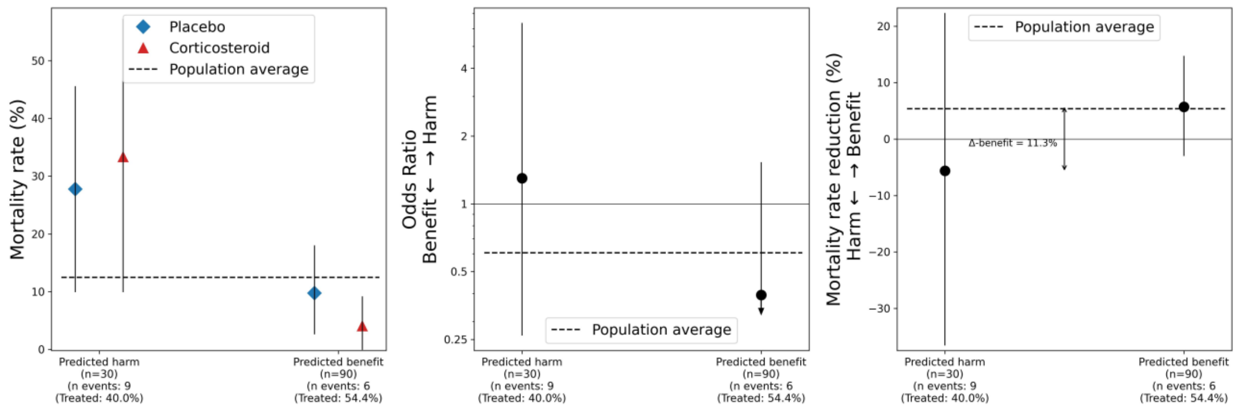

(f) LOTO-CV fold in which Wittermans et al. [13] formed the test cohort

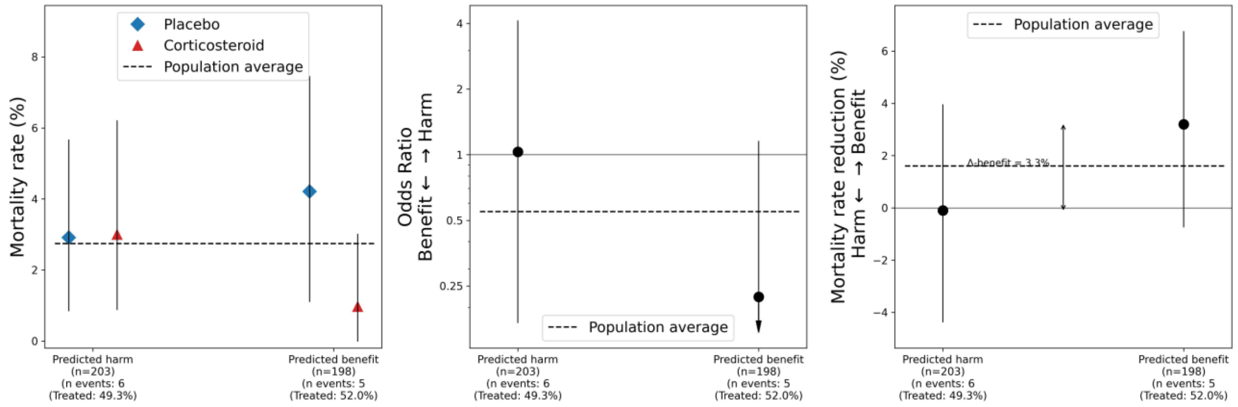

Figure F6: Results for each fold of the leave-one-trial-out cross-validation (LOTO-CV) separately, in procedure 2. Mortality rates (left), odds ratios (middle), and absolute mortality differences (right) are shown with 95% CIs. Number of events and total number of patients per subgroup are shown on the x-axis. In case the odds ratio was not defined (ie, when the mortality rate in the control arm is 0), or the point estimate was zero (ie, when the mortality rate in the treatment arm is 0), this part of the figure is omitted. In case the lower bound of the CI of the odds ratio was zero, this was annotated with an arrow.

(a) LOTO-CV fold in which Confalonieri et al[9] formed the test cohort

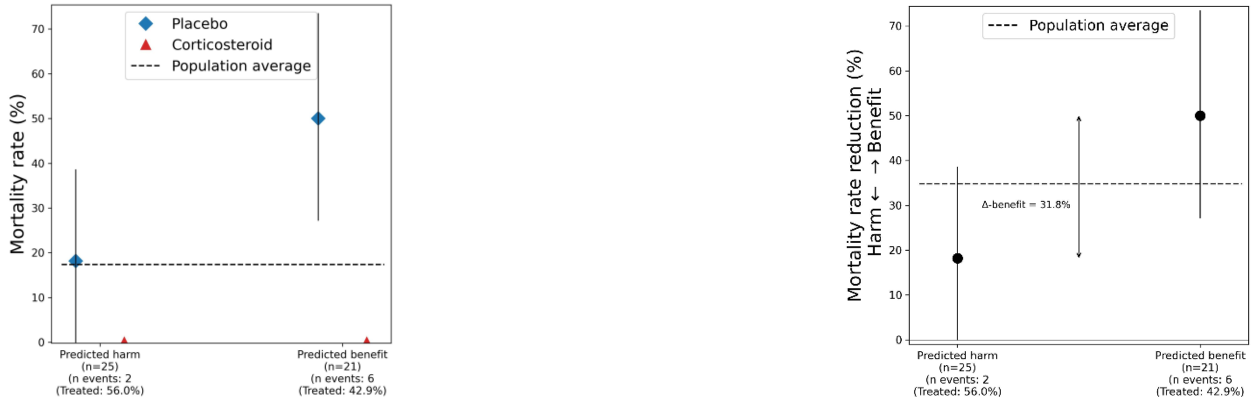

(b) LOTO-CV fold in which Snijders et al[10] formed the test cohort

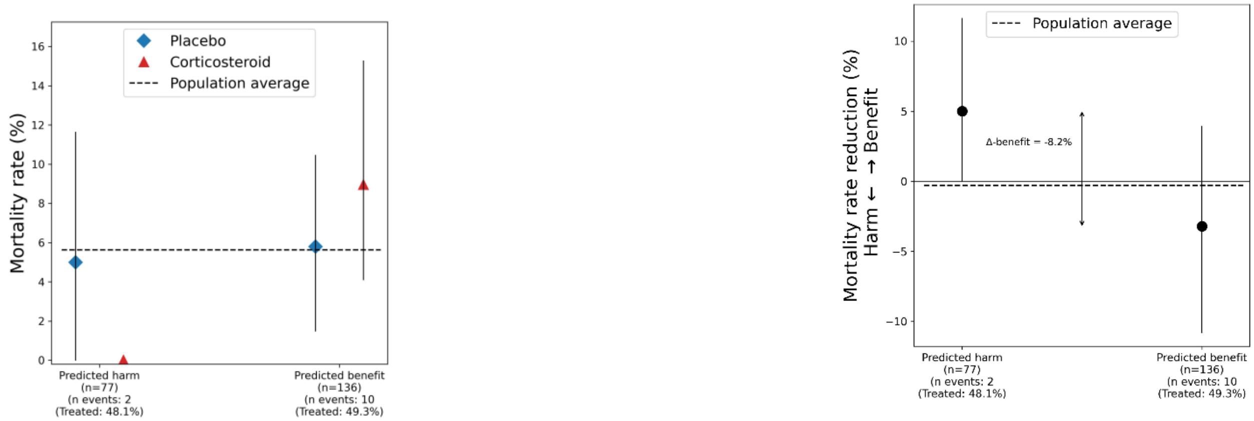

(c) LOTO-CV fold in which Meijvis et al.[11] formed the test cohort

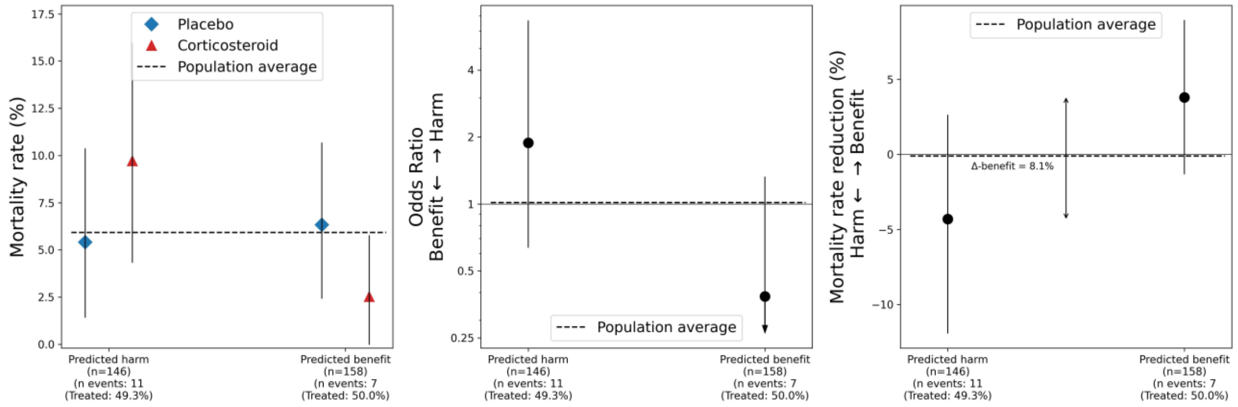

(d) LOTO-CV fold in which Blum et al.[7] formed the test cohort

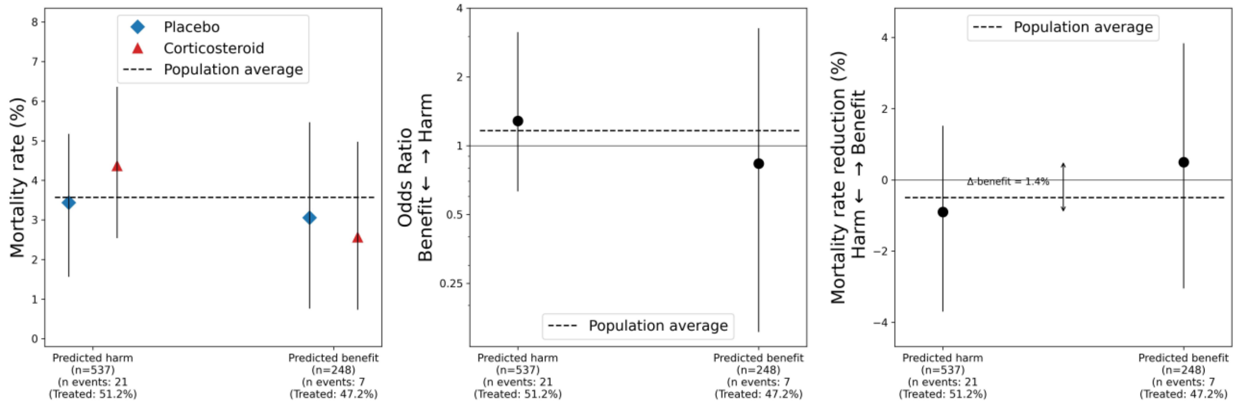

(e) LOTO-CV fold in which Torres et al.[12] formed the test cohort

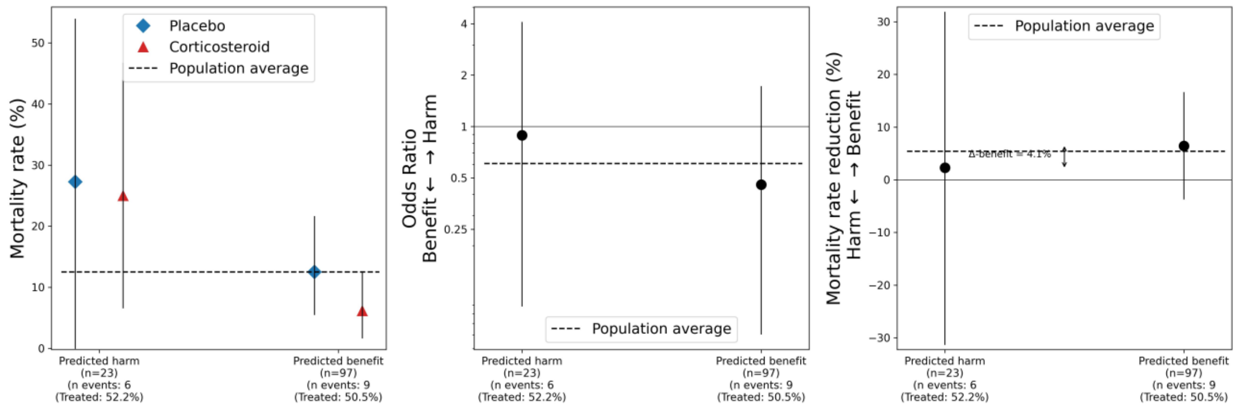

(f) LOTO-CV fold in which Wittermans et al. [13] formed the test cohort

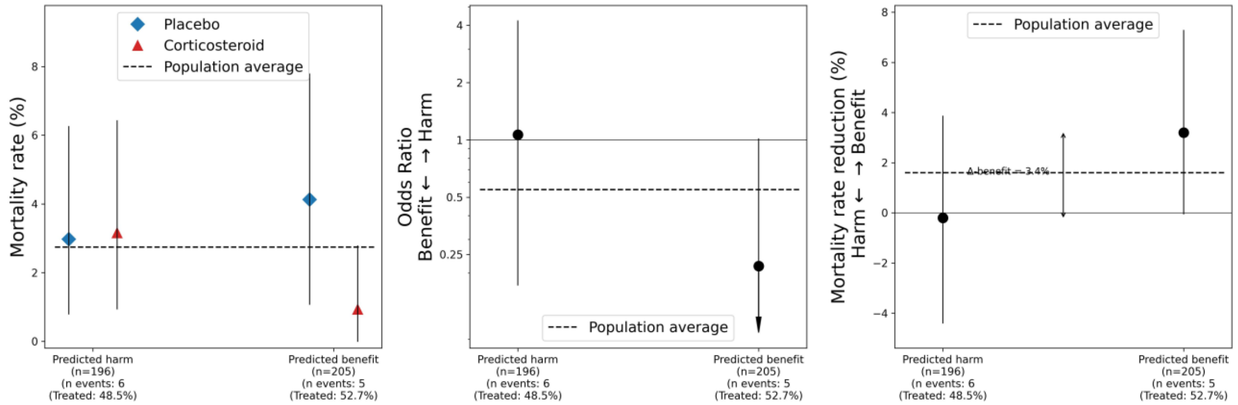

Figure F7: Extra subgroup analysis for low and high baseline glucose levels and the presence of diabetes mellitus (DM).

(a) Relative treatment effects in odds-ratios (with 95% confidence intervals) compared between DM and non-DM patients. On the x-axis, for each group, the number of 30-day mortality events and total number of patients are shown (ie, ‘n events / n patients’). DM was missing for 1 patient (0.05%), and this patient was excluded for this figure.

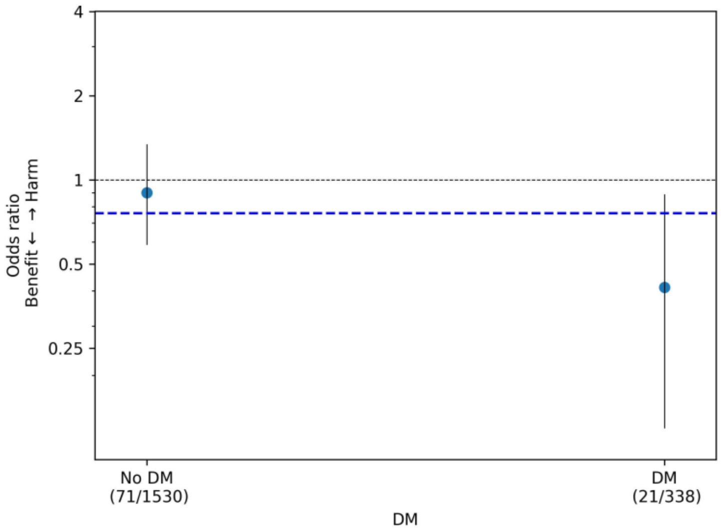

(b) Relative treatment effects in odds-ratios (with 95% confidence intervals) compared between CAP patients with low (ie, ≤ 7 mmol/L) and high (ie, > 7 mmol/L) baseline glucose. On the x-axis, for each group, the number of 30-day mortality events and total number of patients are shown (ie, ‘n events / n patients’). Glucose was missing for 219 patients (11.7%), and these were excluded for this figure.

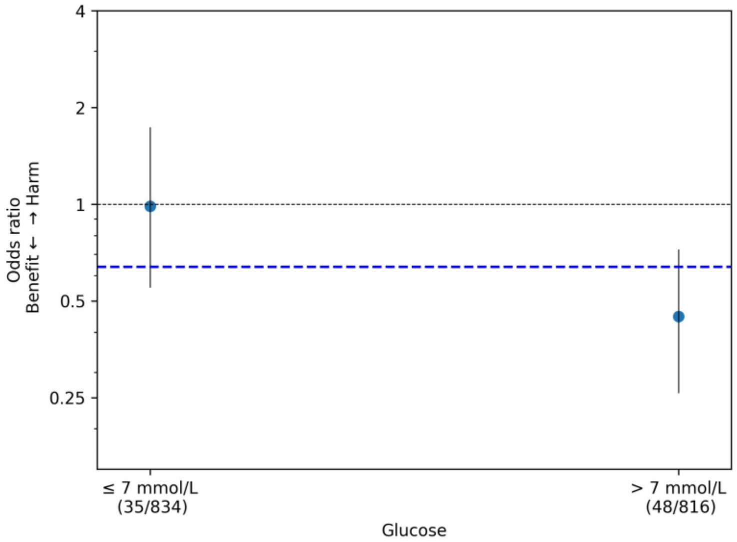

(c) Relative treatment effects in odds-ratios (with 95% confidence intervals) compared between CAP patients with low (ie, ≤ 7 mmol/L) and high (ie, > 7 mmol/L) baseline glucose, stratified for DM and non-DM patients. On the x-axis, for each group, the number of 30-day mortality events and total number of patients are shown (ie, ‘n events / n patients’). Either glucose or DM was missing for 20 patients (11.8%), and these were excluded for this figure. The lower bound of the CI for odds ratio for the DM group with low glucose was zero, denoted with arrow.

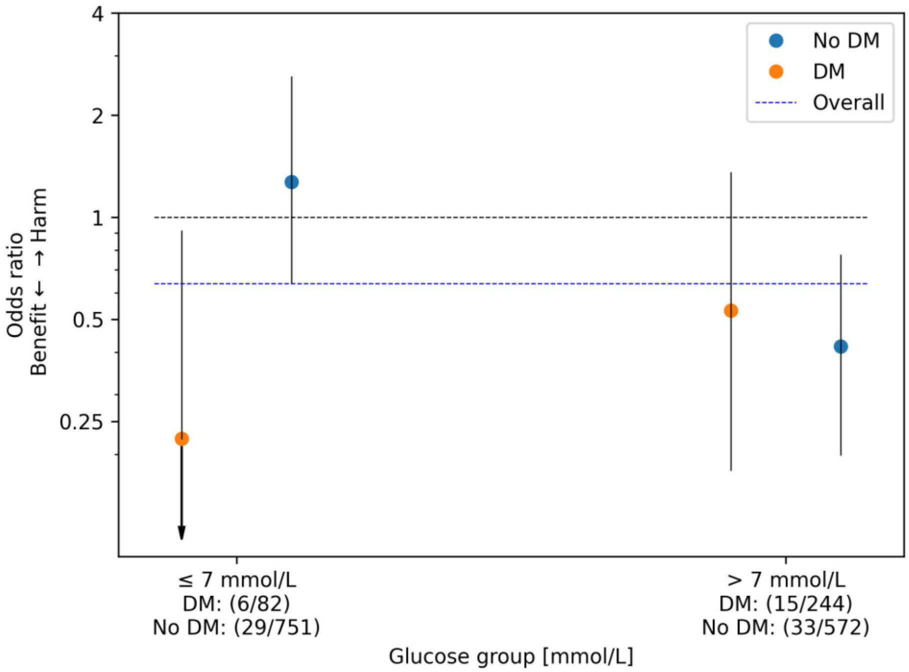

(d) Glucose distributions for DM and non-DM patients within the low (ie, ≤ 7 mmol/L) glucose group.

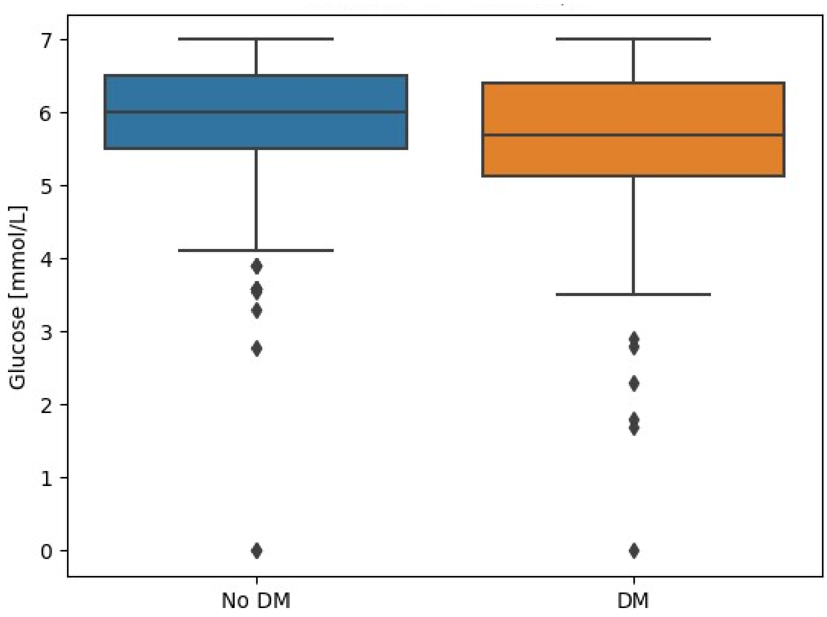

(e) Glucose distributions for DM and non-DM patients within the high (ie, >7 mmol/L) glucose group.

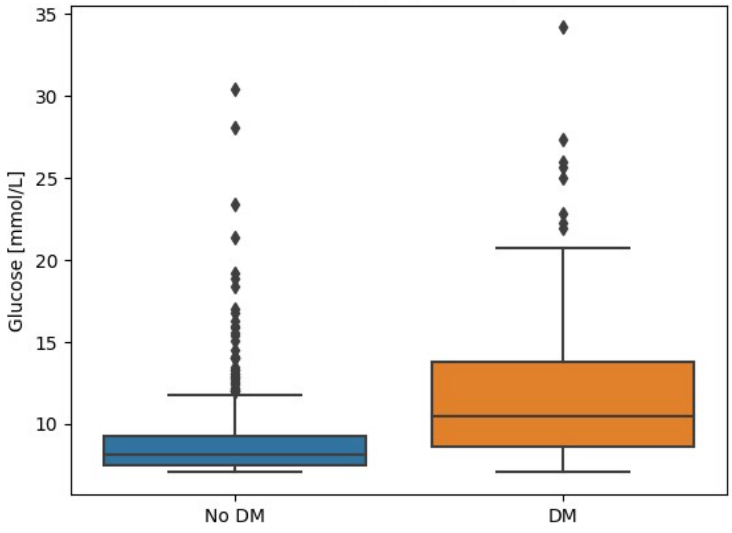

Figure F8: Discriminative performance of procedure 2 (referred to as ‘Effect-8 LASSO, including dichotomized variabels in appendix D), with and without constraint. With constraint, the optimal λ is selected among the λs which resulted in a model with two non-zero weights in each fold of the LOTO-CV procedure. Without constraint, the (overall) optimal λ is selected. The discriminative performance is in terms of AUC-benefits (boxplots were created with 2000 bootstrap samples).

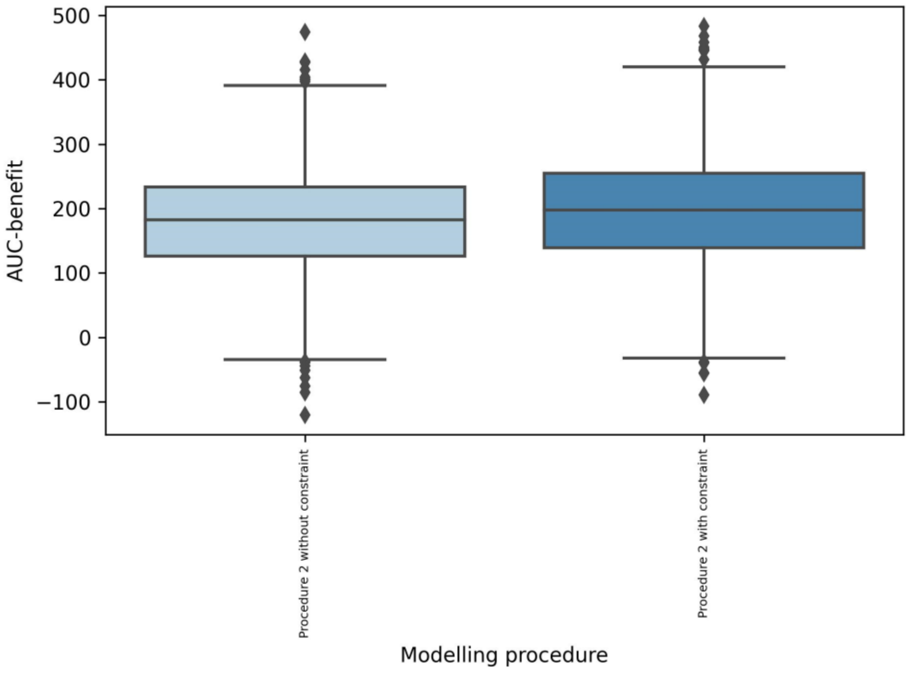

Table F1: The (default) wide and fine grid spaces used in the grid searches. All grids were created evenly spaced on a logarithmic scale. The variable 'center' is defined as $\log^{10}(\lambda_{opt})$, where $\lambda_{opt}$ is the optimal $\lambda$ found in the first, wide grid search.

| Grid search | Searched grid | N steps |
|---|---|---|
| *Wide (default)* | $10^{-2}$ to $10^{2}$ | 50 |
| *Fine* | $10^{(center - 0.3)}$ to $10^{(center + 0.3)}$ | 100 |

Table F2: Results of step 1 of the modelling procedures for each fold in the LOTO-CV procedure: variables were in- or excluded based on availability in both train and test cohorts.

(a) LOTO-CV fold in which Confalonieri et al[9] formed the test cohort

| | Missingness in train cohort (%) | Missingness in test cohort (%) | Excluded |
|---|---|---|---|
| ***Sex*** | 0 | 0 | |
| ***Age*** | 0 | 0 | |
| ***Neoplastic disease*** | 13.4 | 100 | ✓ |
| ***Liver disease*** | 18.7 | 0 | |
| ***CHF*** | 2.7 | 100 | ✓ |
| ***Renal disease*** | 27 | 0 | |
| ***Diabetes Mellitus*** | 0.3 | 0 | |
| ***COPD*** | 0.2 | 0 | |
| ***Resp. rate*** | 14.3 | 0 | |
| ***Dias. blood pressure*** | 6.7 | 0 | |
| ***Syst. blood pressure*** | 6.7 | 0 | |
| ***Temperature*** | 2.6 | 2.2 | |
| ***Heart rate*** | 2.5 | 0 | |
| ***$SaO_2$*** | 13.9 | 0 | |
| ***Creatinine*** | 1.2 | 0 | |
| ***Sodium*** | 1 | 4.3 | |
| ***Urea*** | 7.2 | 0 | |
| ***CRP*** | 2.1 | 0 | |
| ***Glucose*** | 11.6 | 0 | |
| ***WBC count*** | 0.9 | 4.3 | |

(b) LOTO-CV fold in which Snijders et al[10] formed the test cohort

| | Missingness in train cohort (%) | Missingness in test cohort (%) | Filtered |
|---|---|---|---|
| ***Sex*** | 0 | 0 | |
| ***Age*** | 0 | 0 | |
| ***Neoplastic disease*** | 5.8 | 100 | ✓ |
| ***Liver disease*** | 9.2 | 100 | ✓ |
| ***CHF*** | 5.3 | 0.5 | |
| ***Renal disease*** | 18.2 | 100 | ✓ |
| ***Diabetes Mellitus*** | 0.3 | 0 | |
| ***COPD*** | 0.2 | 0 | |
| ***Resp. rate*** | 15.5 | 0.5 | |
| ***Dias. blood pressure*** | 7.3 | 0.5 | |
| ***Syst. blood pressure*** | 7.3 | 0.5 | |
| ***Temperature*** | 2.9 | 0 | |
| ***Heart rate*** | 2.7 | 0.5 | |
| ***$SaO_2$*** | 13.4 | 16 | |
| ***Creatinine*** | 1.3 | 0.5 | |
| ***Sodium*** | 1.2 | 0 | |
| ***Urea*** | 7.7 | 0.9 | |
| ***CRP*** | 2.3 | 0 | |
| ***Glucose*** | 12.2 | 4.2 | |
| ***WBC count*** | 1.1 | 0 | |

(c) LOTO-CV fold in which Meijvis et al.[11] formed the test cohort

| | Missingness in train cohort (%) | Missingness in test cohort (%) | Filtered |
|---|---|---|---|
| ***Sex*** | 0 | 0 | |
| ***Age*** | 0 | 0 | |
| ***Neoplastic disease*** | 17.5 | 1.6 | |
| ***Liver disease*** | 21.4 | 0 | |
| ***CHF*** | 5.7 | 0 | |
| ***Renal disease*** | 30.8 | 0 | |
| ***Diabetes Mellitus*** | 0.3 | 0 | |
| ***COPD*** | 0.2 | 0 | |
| ***Resp. rate*** | 15.3 | 5.9 | |
| ***Dias. blood pressure*** | 7.4 | 2 | |
| ***Syst. blood pressure*** | 7.4 | 2 | |
| ***Temperature*** | 2.8 | 1.6 | |
| ***Heart rate*** | 2.6 | 1.6 | |
| ***$SaO_2$*** | 11.1 | 28.6 | |
| ***Creatinine*** | 1.1 | 2 | |
| ***Sodium*** | 0.9 | 1.6 | |
| ***Urea*** | 7.6 | 3.6 | |
| ***CRP*** | 2.1 | 1.6 | |
| ***Glucose*** | 12.2 | 6.2 | |
| ***WBC count*** | 0.9 | 1.6 | |

### (d) LOTO-CV fold in which Blum et al.[7] formed the test cohort

| | Missingness in train cohort (%) | Missingness in test cohort (%) | Filtered |
|---|---|---|---|
| ***Sex*** | 0 | 0 | |
| ***Age*** | 0 | 0 | |
| ***Neoplastic disease*** | 24.2 | 0 | |
| ***Liver disease*** | 29 | 0 | |
| ***CHF*** | 7.7 | 0 | |
| ***Renal disease*** | 41.8 | 0.1 | ✓ |
| ***Diabetes Mellitus*** | 0.4 | 0.1 | |
| ***COPD*** | 0.3 | 0 | |
| ***Resp. rate*** | 10.7 | 19.5 | |
| ***Dias. blood pressure*** | 10 | 0.8 | |
| ***Syst. blood pressure*** | 10 | 0.8 | |
| ***Temperature*** | 3.6 | 1 | |
| ***Heart rate*** | 3.5 | 0.8 | |
| ***$SaO_2$*** | 19.6 | 3.4 | |
| ***Creatinine*** | 1.5 | 0.8 | |
| ***Sodium*** | 1.4 | 0.4 | |
| ***Urea*** | 8.1 | 5.4 | |
| ***CRP*** | 2.8 | 0.8 | |
| ***Glucose*** | 4 | 24.1 | |
| ***WBC count*** | 1.3 | 0.4 | |

### (e) LOTO-CV fold in which Torres et al.[12] formed the test cohort

| | Missingness in train cohort (%) | Missingness in test cohort (%) | Filtered |
|---|---|---|---|
| ***Sex*** | 0 | 0 | |
| ***Age*** | 0 | 0 | |
| ***Neoplastic disease*** | 16.2 | 0 | |
| ***Liver disease*** | 13.4 | 100 | ✓ |
| ***CHF*** | 5.1 | 0 | |
| ***Renal disease*** | 22 | 100 | ✓ |
| ***Diabetes Mellitus*** | 0.3 | 0 | |
| ***COPD*** | 0.2 | 0 | |
| ***Resp. rate*** | 13.1 | 29.2 | |
| ***Dias. blood pressure*** | 1 | 100 | ✓ |
| ***Syst. blood pressure*** | 1 | 100 | ✓ |
| ***Temperature*** | 1 | 30 | |
| ***Heart rate*** | 0.9 | 28.3 | |
| ***$SaO_2$*** | 8.5 | 100 | ✓ |
| ***Creatinine*** | 1.2 | 1.7 | |
| ***Sodium*** | 0.9 | 3.3 | |
| ***Urea*** | 3.3 | 70 | ✓ |
| ***CRP*** | 1.7 | 8.3 | |
| ***Glucose*** | 12 | 1.7 | |
| ***WBC count*** | 0.8 | 3.3 | |

(f) LOTO-CV fold in which Wittermans et al. [13] formed the test cohort

| | Missingness in train cohort (%) | Missingness in test cohort (%) | Filtered |
|---|---|---|---|
| ***Sex*** | 0 | 0 | |
| ***Age*** | 0 | 0 | |
| ***Neoplastic disease*** | 18.8 | 0 | |
| ***Liver disease*** | 22.6 | 0 | |
| ***CHF*** | 6 | 0 | |
| ***Renal disease*** | 22.6 | 42.6 | ✓ |
| ***Diabetes Mellitus*** | 0.3 | 0 | |
| ***COPD*** | 0.2 | 0 | |
| ***Resp. rate*** | 17.2 | 0 | |
| ***Dias. blood pressure*** | 8.1 | 0 | |
| ***Syst. blood pressure*** | 8.1 | 0 | |
| ***Temperature*** | 3.2 | 0 | |
| ***Heart rate*** | 3.1 | 0 | |
| ***$SaO_2$*** | 16.8 | 0 | |
| ***Creatinine*** | 1.4 | 0.5 | |
| ***Sodium*** | 1.3 | 0 | |
| ***Urea*** | 8.7 | 0 | |
| ***CRP*** | 2.5 | 0.2 | |
| ***Glucose*** | 14 | 0 | |
| ***WBC count*** | 1.2 | 0 | |

Table F3: Baseline characteristics of the patients in the predicted harm and the predicted benefit group (identified in procedure 2). Data are n (%) or median (IQR). *PSI values are missing for three (0.2%) of the patients, therefore the total numbers in the severity classes do not add up to total number of patients in the two groups.

| | **Predicted harm group (N=1,004)** | **Predicted benefit group (N=865)** |
|---|---|---|
| ***Demographics*** | | |
| *Female sex* | 363 (36.2) | 382 (44.2) |
| *Age, (years)* | 73.0 (56.0-82.0) | 67.0 (56.0-77.0) |
| ***Clinical parameters*** | | |
| *Resp. rate, (breaths/min)* | 21.0 (18.0-25.5) | 24.0 (18.0-30.0) |
| *Dias. blood pressure, (mmHg)* | 71.0 (61.0-80.0) | 70.0 (60.0-80.0) |
| *Syst. blood pressure, (mmHg)* | 129.0 (113.0-143.0) | 125.0 (110.0-140.0) |
| *Temperature, (°C)* | 37.8 (37.1-38.6) | 38.1 (37.2-38.9) |
| *Heart rate, (bpm)* | 88.0 (75.8-101.0) | 98.0 (84.0-110.0) |
| *$SaO_2$, (%)* | 95.0 (92.0-97.0) | 94.0 (91.0-96.0) |
| ***Laboratory values*** | | |
| *Creatinine, (µmol/L)* | 86.0 (69.0-111.0) | 94.0 (75.0-125.0) |
| *Sodium, (mmol/L)* | 137.0 (134.0-139.0) | 134.0 (132.0-137.0) |
| *Urea, (mmol/L)* | 6.4 (4.5-9.8) | 7.1 (5.1-10.8) |
| *CRP, (mg/L)* | 111.0 (54.0-172.0) | 299.4 (241.0-365.0) |
| *Glucose, (mmol/L)* | 6.5 (5.7-7.7) | 7.6 (6.4-9.2) |
| *WBC count, ($10^9$ cells/L)* | 11.6 (8.5-15.2) | 14.1 (10.4-19.4) |
| ***Comorbidities*** | | |
| *Neoplastic disease* | 82 (8.2) | 38 (4.4) |
| *Liver disease* | 19 (1.9) | 20 (2.3) |
| *Congestive heart failure* | 190 (18.9) | 121 (14.0) |
| *Renal disease* | 200 (19.9) | 96 (11.1) |
| *Diabetes mellitus* | 167 (16.6) | 171 (19.8) |
| *Chronic obstructive pulmonary disease* | 159 (15.8) | 148 (17.1) |
| ***PSI**** | | |
| *Total score* | 89.0 (65.0-113.0) | 89.0 (65.0-114.0) |
| *Class I* | 140 (13.9) | 98 (11.4) |
| *Class II* | 158 (15.7) | 168 (19.5) |
| *Class III* | 217 (21.6) | 184 (21.3) |
| *Class IV* | 365 (36.4) | 289 (33.5) |
| *Class V* | 124 (12.4) | 123 (14.3) |

### References

1 Sterne JAC, Savović J, Page MJ, *et al.* RoB 2: A revised tool for assessing risk of bias in randomised trials. *BMJ* 2019; **366**: 1–8.

2 Dequin P-F, Meziani F, Quenot J-P, *et al.* Hydrocortisone in Severe Community-Acquired Pneumonia. *N Engl J Med* 2023; **388**: 1931–41.

3 Nafae RM, Ragab MI, Amany FM, Rashed SB. Adjuvant role of corticosteroids in the treatment of community-acquired pneumonia. *Egypt J Chest Dis Tuberc* 2013; **62**: 439–45.

4 Sabry NA, Omar EE-D. Corticosteroids and ICU Course of Community Acquired Pneumonia in Egyptian Settings. *Pharmacol & Pharm* 2011; **02**: 73–81.

5 Meduri GU, Shih M-C, Bridges L, *et al.* Low-dose methylprednisolone treatment in critically ill patients with severe  community-acquired pneumonia. *Intensive Care Med* 2022; **48**: 1009–23.

6 Seabold S, Perktold J. Statsmodels: Econometric and statistical modeling with python. In: 9th Python in Science Conference. 2010.

7 Blum CA, Nigro N, Briel M, *et al.* Adjunct prednisone therapy for patients with community-acquired pneumonia: A multicentre, double-blind, randomised, placebo-controlled trial. *Lancet* 2015; **385**: 1511–8.

8 Kent DM, Paulus JK, Van Klaveren D, *et al.* The Predictive Approaches to Treatment effect Heterogeneity (PATH) statement. *Ann Intern Med* 2020; **172**: 35–45.

9 Confalonieri M, Urbino R, Potena A, *et al.* Hydrocortisone infusion for severe community-acquired pneumonia: A preliminary randomized study. *Am J Respir Crit Care Med* 2005; **171**: 242–8.

10 Snijders D, Daniels JMA, De Graaff CS, Van Der Werf TS, Boersma WG. Efficacy of corticosteroids in community-acquired pneumonia: A randomized double-blinded clinical trial. *Am J Respir Crit Care Med* 2010; **181**: 975–82.

11 Meijvis SCA, Hardeman H, Remmelts HHF, *et al.* Dexamethasone and length of hospital stay in patients with community-acquired pneumonia: A randomised, double-blind, placebo-controlled trial. *Lancet* 2011; **377**: 2023–30.

12 Torres A, Sibila O, Ferrer M, *et al.* Effect of corticosteroids on treatment failure among hospitalized patients with severe community-acquired pneumonia and high inflammatory response: A randomized clinical trial. *JAMA* 2015; **313**: 677–86.

13 Wittermans E, Vestjens SMT, Spoorenberg SMC, *et al.* Adjunctive treatment with oral dexamethasone in non-ICU patients hospitalised with community-acquired pneumonia: A randomised clinical trial. *Eur Respir J* 2021; **58**: 1–10.

14 Smit J. Predicting individualized treatment effects of corticosteroids in community-acquired-pneumonia. https://github.com/jimmsmit/HTE_CAP.

15 Kane S. Corticosteroid Conversion Calculator. ClinCalc. https://clincalc.com/corticosteroids (accessed Jan 25, 2023).

16 Average sizes of men and women. https://www.worlddata.info/average-bodyheight.php.

17 Endeman H, Meijvis SCA, Rijkers GT, *et al.* Systemic cytokine response in patients with community-acquired pneumonia. *Eur Respir J* 2011; **37**: 1431–8.
